# Phylo-Plex: A phylogenetically informed, low-cost amplicon sequencing platform for deployable high-resolution genomic epidemiology

**DOI:** 10.1101/2025.02.19.25322526

**Authors:** Mathew A Beale, Vignesh Shetty, Kirsty E Ambridge, George Lacey, Sam Dougan, William Roberts-Sengier, Beth Sampher, Florent Lassalle, Matthew J Dorman, Mahlape P Mahlangu, Johanna ME Venter, Bianca Da Costa Dias, Martha Chipinduro, Tendai M. Washaya, Luanne Rodgers, Beauty Makamure, Ethel Dauya, Michael Marks, Etienne E. Müller, Rashida A Ferrand, Nicholas R Thomson

**Affiliations:** Parasites and Microbes Programme, Wellcome Sanger Institute, Cambridgeshire, UK; School of Mathematical and Statistical Sciences, College of Science and Engineering, University of Galway, University Road, Galway, Ireland; Department of Microbiology, Moyne Institute of Preventive Medicine, School of Genetics and Microbiology, Trinity College Dublin, Dublin, Ireland; Centre for HIV C STIs, National Institute for Communicable Diseases, Johannesburg, South Africa; Biomedical Research and Training Institute, Harare, Zimbabwe; Faculty of Medicine and Health Sciences, Midlands State University, Gweru, Zimbabwe; Faculty of Infectious and Tropical Diseases, London School of Hygiene and Tropical Medicine, London, UK; Hospital for Tropical Diseases, University College London Hospital, London, UK; Division of Infection and Immunity, University College London, London, UK

## Abstract

Genomic pathogen surveillance is a powerful tool for public health and research, but is costly and unachievable in low-resource settings. Most sub-genomic typing methods sacrifice resolution whilst remaining costly. We developed “Phylo-Plex”, a novel approach that identifies information-rich genomic regions to maximise phylogenetic information whilst minimising the number of regions. Applied to *Treponema pallidum* and *Neisseria gonorrhoeae*, we designed a high-resolution multiplex PCR sequencing scheme for lineage tracking pathogens with different extremes of genome variation. For *Treponema pallidum*, we also designed and evaluated the Phylo-Plex scheme in the laboratory and field settings by sequencing 72 clinical samples using MinION Flongle cells. Our *T. pallidum* scheme comprising 59 multiplex amplicons achieved high discrimination of fine-scale sublineages comparable to those defined using whole genomes, and demonstrating a qPCR detection limit ≤Ct 32. Variant calls from MinION amplicon sequencing were highly correlated with Illumina whole genome sequencing. We successfully deployed the method in a low-resource laboratory in Zimbabwe, costed at <£300/24 samples (£12.47/sample). Phylo-Plex enables low-cost tracking of priority pathogenic lineages in low resource settings and at scale.

## Introduction

Genomic pathogen surveillance has proven itself a powerful tool for both public health epidemiology and research^1^. Discriminating closely related isolates and deconvoluting complex samples and communities can enable finescale global tracking of pathogen populations, provide insights into epidemiology, transmission and the impact of clinical interventions such as antimicrobial usage or vaccines^2^. However, whole genome sequencing (WGS) pathogens at scale has substantial barriers to implementation including cost, complexity of data analysis, and time to results. This is particularly the case for low-resource settings or where pathogens cannot be readily cultured, necessitating direct or enriched metagenomic sequencing from clinical swabs and complex analytics^3,4^. Currently, the only molecular alternatives to WGS are typing approaches such as single gene or multi-locus sequence typing (MLST). Although designed for very different purposes, compared to WGS, typing is by definition comparatively low resolution and is not responsive to the evolutionary dynamics of the target pathogen^5–8^. A solution to this is multiplex PCR^9–12^ approaches such as the ARTIC network protocol (https://artic.network/)^11^, which amplifies the whole genome in a series of tiled amplicons, and was used during the SARS-CoV-2 pandemic to generate over 18 million complete viral genome sequences. This approach allowed near real-time tracking and, because of the small size of the genome (30Kb), for comprehensive identification of variants of interest and concern globally. However, this approach is not cost effective for generating larger complete bacterial genomes which range between 1-8 Mb in length, necessitating a more targeted approach.

Syphilis, caused by the bacterium *Treponema pallidum subspecies pallidum*, is an important sexually transmitted infection. Globally, *T. pallidum* causes 8 million new cases per annum and 700,000 cases of congenital syphilis leading to 220,000 early foetal deaths, stillbirths or neonatal deaths^13^. In most countries diagnosis of syphilis is restricted to either serological testing (in the context of antenatal care) or syndromic management (in the context of symptomatic individuals). *T. pallidum* is extremely difficult to culture^14^ and so whole genome-based surveillance of syphilis is not widely used, even in high resource settings. Until now the majority of available genomes have been sequenced with costly sequence capture enriched metagenomic approaches which require detailed bioinformatic analysis and significant computational infrastructure^8,15–17^. *T. pallidum* is subdivided into three phylogenetically distinct subspecies (subspecies *pallidum*, *pertenue* and *endemicum*), and subspecies *pallidum* comprises two main lineages (Nichols and SS14), as well as a number of sublineages^15^. Molecular typing schemes include a variable tandem repeat method^18^, a three-gene MLST scheme^7^, and a recently described seven-gene MLST^19^.

In this study, we developed and employed a novel whole genome informed approach for selecting optimal genome regions which can be targeted for multiplex PCR, dubbed “Phylo-Plex”. Given a representative collection of pathogen genomes, Phylo-Plex clusters single nucleotide polymorphisms (SNPs) along lineage-defining phylogenetic branches into information-rich amplicons which maximise resolution whilst minimising the number of genomic regions required for sequencing. Whilst Phylo-Plex is not intended to replace formal typing schemes, such as MLST, this approach yields high-resolution phylogenies which recapitulate population structures obtained from WGS data, and can be performed quickly and easily at a fraction of the cost. We applied the Phylo-Plex approach to identify optimal regions for PCR sequencing for both a low diversity pathogen (*T. pallidum*) and a high diversity pathogen (*Neisseria gonorrhoeae*). For *T. pallidum*, we then developed a high resolution Phylo-Plex scheme comprising 59 multiplex amplicons. We deployed our method in Harare, Zimbabwe and show how, when paired with low-cost nanopore sequencing, this approach provides a fast and flexible framework for SNP-based molecular sequence typing, that can be performed in a low resource setting at low cost. The value of Phylo-Plex is in its universal applicability to any pathogen where we have existing genomic data and key lineages need to be found or priority pathogens need to be tracked in any setting.

## Results

The Phylo-Plex approach we developed here takes WGS data from a representative global population framework, and identifies all discriminating SNPs linked to the major topological features (lineages, sublineages, etc) within the phylogeny. Then, regardless of the sublineage supported, all discriminatory SNPs are positionally clustered along the genome and used in a hierarchical selection algorithm to find the optimal SNP set which maximises the discriminatory power for each lineage, whilst minimising the number of amplicons required. Candidate regions are then used for multiplex PCR primer design, which can be validated *in silico* and *in vitro* before deployment (Figure 1).

**Figure 1.**
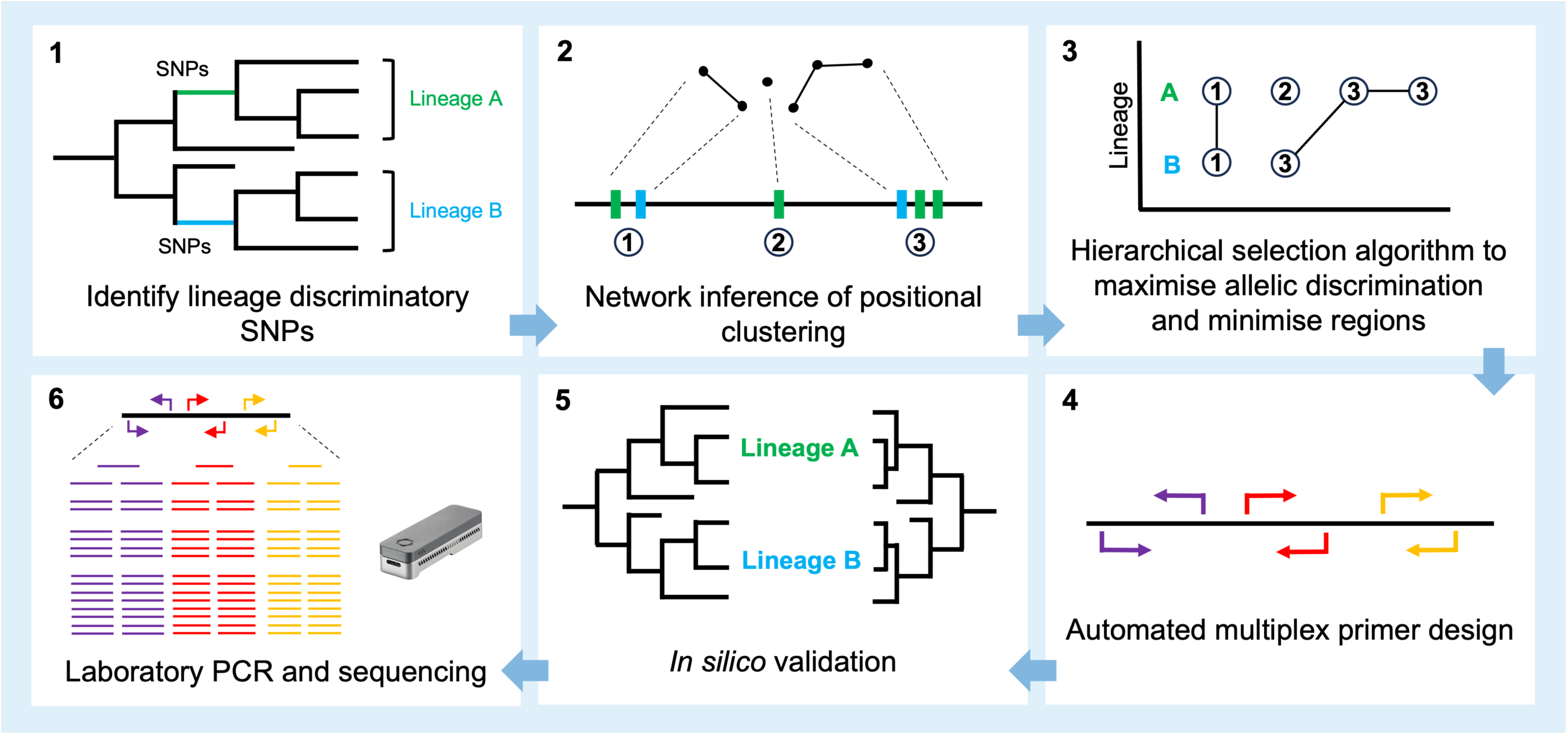
Overview of Phylo-Plex workflow. 1 – Pathogen genome populations are analysed to determine SNPs that delineate individual lineages along ancestral branches. 2 – Discriminatory SNPs are clustered based on their genome position to identify information-rich candidate regions. 3 – Hierarchical selection of candidate regions maximises discrimination for each lineage whilst minimising the number of regions chosen. 4 – Selected regions are used for automated multiplex primer design. 5 – Primer designs are validated *in silico* to ensure the final scheme will amplify efficiently and without contamination, and that genome population structures are accurately reconstructed. 6 – Primers are then optimized and validated in the laboratory using sequencing before deployment.

### Identifying population structure defining SNPs

We constructed a globally representative dataset of 607 publicly available *T. pallidum* genomes (as of 2022), including high-quality genomes with ≥95% of reference positions at ≥5X coverage, and spanning *T. pallidum* subspecies *pallidum* (TPA, n=530), *T. pallidum* subspecies *pertenue* (TPE, n=75) and *T. pallidum* subspecies *endemicum* (TEN, n=2) (Supplementary Table 1). Notably, the dataset was enriched for genomes from North America and Europe, and underrepresented for genomes from lower- and middle-income countries, particularly in Africa, reflecting biases in publicly available data^15^. Sequencing reads were mapped to a common reference genome (NC_021508.1) and hypervariable and recombining regions were masked using an initial list of known recombinant genes, followed by *de novo* inference of recombination using Gubbins^20^. We inferred a whole genome phylogeny using IQ-tree^21,22^, and designated phylogenetic sublineages using rPinecone^23^ v0.1.0 with a threshold of up to 10 SNPs from the common ancestral node. This resulted in 40 sublineages and 33 singletons, including the 17 sublineages previously defined within TPA^15^, and 23 newly-defined sublineages within TPE (Supplementary Figure 1).

Next, within this population structure we identified vertically inherited discriminatory SNPs by applying population genomic statistical tests of allelic segregation to a recombination-masked alignment, using Fixation Index^24^ (F_ST_) to define a list of SNPs that discriminate sublineages. Given that most *T. pallidum* genomic data are metagenomically derived and frequently contain uncertain genomic positions due to low coverage, this approach enabled delineation of informative SNPs whilst accommodating missing data. Retaining SNPs with F_ST_ ≥0.9, a combination of multiple SNPs together enables robust identification of each lineage or sublineage (Supplementary Figure 2). We applied this approach to each of the 40 sublineages defined in our population, as well as to delineate subspecies (TPA and TPE) and TPA major lineages (Nichols and SS14), resulting in a single list comprising 1549 discriminatory SNPs, of which 855 occurred on phylogenetic branches leading to individual sublineages, and thus discriminatory for sublineages, whilst the remaining 694 discriminatory SNPs fell on deeper branches and distinguished *T. pallidum* subspecies and major lineages (Supplementary Figure 3).

Examining the distribution of the 855 SNPs discriminating sublineages, we found that the number of SNPs along discriminatory branches of the phylogeny differed between sublineages (median 26 SNPs/sublineage, range 1-153; Supplementary Figure 4). Consistent with previous findings^15^, the power to distinguish sublineages within the highly clonal SS14-lineage of TPA was limited (median 4 SNPs/sublineage, range 1-6). In contrast, sublineages within Nichols-lineage TPA (median 20.5 SNPs/sublineage, range 7-60) and in TPE had much higher discrimination (median 39 SNPs/sublineage, range 16-153), reflecting more deeply branching sublineages in these parts of the phylogeny.

### SNP refinement and hierarchical selection of candidate regions

To design an efficient amplicon scheme, we needed to optimise the choice of SNPs to maximise useful information whilst minimising the overall number of amplicons. To ensure multiplex PCR was efficient, we also needed to ensure amplicons were kept small (<1000 bp), and had similar lengths and melting temperatures. Examining the genomic positions of discriminatory SNPs, we found that although these SNPs were widely distributed across the genome for each sublineage (median 13,164 bp between SNPs, Supplementary Figure 5), they could be positionally clustered when analysed as a population (median 694 bp between SNPs). We evaluated the effect of clustering SNPs based on their genome position, forming network edges between SNPs based on different distance thresholds, and selected a distance of 300 bp since this clustered a high number of SNPs whilst keeping the majority of clusters below 1000 bp in length (Supplementary Figure 6). Of our initial 1549 discriminatory SNPs, 988 sites fell within 300 bp of at least one other discriminatory site, suggesting that this was an effective method for identifying localised phylogenetically information-rich regions of the genome (Figure 2A).

**Figure 2.**
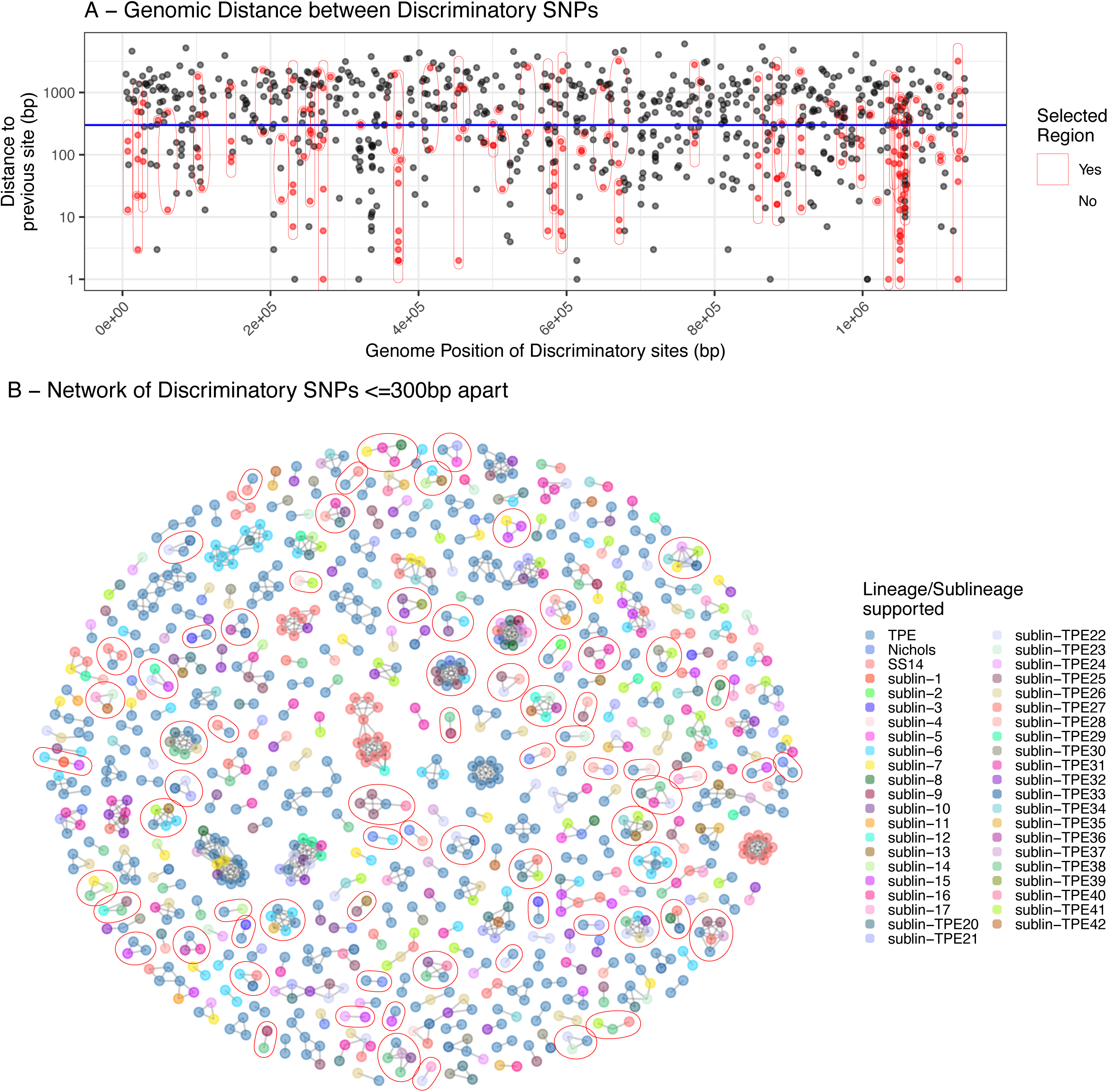
Positional clustering of discriminatory SNPs enables identification of information rich regions for PCR. A – Genomic distance between discriminatory SNPs for 40 sublineages. Blue line indicates 300 bp distance between sites. Black and Red points represent all discriminatory SNP-sites. Red points represent the clustered location-optimised SNPs represented in final amplicon panel design. B – Network showing the discriminatory SNPs coloured by the lineage they define and clustered by genome position. Nodes indicate individual SNPs and are coloured according to the sublineage supported. Edges indicate SNPs ≤ 300bp from each other, and form clusters of information-rich genomic regions. Red rings indicate clusters included in the final design.

We constructed a network of clustered discriminatory SNPs using pairwise positional genome distances, allowing edges where SNPs were separated by ≤300 bp, and identifying 311 distinct network components, each containing >1 discriminatory sites (median 2, range 2-15) - henceforth referred to as ‘candidate regions’ (Figure 2B). For the remaining 561 discriminatory SNPs not included within a network component (i.e. singleton sites not positionally linked to another), we identified these as additional singleton regions, leaving a total of 872 candidate regions comprising 1549 individual SNPs. Of these, we excluded 50/872 larger candidate regions with total length ≥385 bp to minimise the PCR amplicon length, leaving 822 candidate regions.

Given the varying levels of SNP support for the different sublineages of TPA and TPE, we needed to optimise the number of potential amplicons whilst maintaining support for each sublineage. We designed an algorithm to iteratively select candidate regions to maximise the number of discriminatory sites for each sublineage, whilst minimising the total number of regions included in the final scheme (Supplementary Figure 7). We considered the importance of maximising SNPs supporting each sublineage, whilst incorporating redundancy for PCR failures - aiming to avoid any single region being the sole supporter of a sublineage (which could lead to biases driven by hypervariable regions or residual recombination, as well as the risk of losing all support should PCR for that region fail).

Briefly, we constructed a ranked list of candidate regions based on the number of distinct sublineages supported by the SNPs within each region. Using an iterative loop, where initial support for each sublineage was considered to be zero regions, each candidate region was evaluated for its SNP support of specific sublineages. If a region contained SNPs that supported a sublineage, the counts for each supported sublineage were updated and the region was added to the scheme, unless the sublineage had already reached a minimum threshold of regions. For the *T. pallidum* scheme, we selected a minimum threshold of three regions, representing a compromise to provide robust discrimination whilst keeping the total number of amplicons small. Once a sublineage reached this threshold, new regions were not evaluated against it unless they contributed to another sublineage. To minimise potential biases driven by hypervariable regions or residual recombination, counts were increased by one for each sublineage supported by a candidate region even if that region contained multiple discriminatory sites to that sublineage. This process was repeated iteratively for all sublineages and regions. Consequently, our algorithm ensures all sublineages contain at least the minimum threshold discriminatory sites, unless there were fewer than this threshold initially available, whilst some sublineages (typically those representing more deeply branching lineages) contain more than the minimum (as subsequent sublineage picks may enrich discriminatory sites for other sublineages).

Our Phylo-Plex algorithm prioritised 76/822 candidate genomic regions that define the 40 sublineages within the dual global phylogenetic framework of TPA and TPE (Supplementary Figure 4). We re-examined the discriminatory power for each sublineage using this revised list and found that support for each sublineage was maintained, whilst the variability in terms of sites per sublineage was greatly reduced (mean 5.5 SNPs, range 1-48) (Supplementary Figure 4). To ensure similar PCR amplification dynamics and to make room for flanking primer design, we extended the length of each Phylo-Plex amplicon (we termed “Phy-cons”) to 700 bp by adding equal length flanking sequences to each end. We used these candidate regions for automated design of 20- 24 bp primers with 59–63°C melting temperatures with PrimalScheme^10,11^ (Supplementary Table 2), which produced 72 primer pairs with an amplicon size distribution of 511-628 bp (4/76 candidate regions were unsuited to multiplex primer design and were excluded). We also designed and integrated a primer set for recovering the ribosomal 23S region (important for inferring antimicrobial resistance to macrolides in *T. pallidum* – in different pathogens, gene-specific markers for other resistance or virulence determinants could be used) to complete the final amplicon scheme of 73 Phy-cons. All 73 primer pair designs were initially evaluated for specificity using primer BLAST and *in silico* PCR (see Methods).

### Evaluation of laboratory sequencing and bioinformatics

Next, we evaluated the robustness of the laboratory and bioinformatic methods. We performed initial validation using ten-fold dilution series of genomic DNA extracted from four rabbit-passaged reference strains. We then selected 72 clinical swab samples from PCR-confirmed syphilis patients with a *Treponema* qPCR Ct between 23.9-32.2 from South Africa, and generated multiplex PCR amplicons in batches of 24 using the *Treponema* Phylo-Plex scheme (TP-Phylo-Plex). Because all Phy-cons were of a similar size, we used agarose gels to confirm overall pooled PCR performance, and sequencing coverage metrics to evaluate individual amplicon performance.

To ensure our scheme was easily and affordably deployable in low resource settings, we opted for MinION sequencing (Oxford Nanopore). We ligated individual sample barcodes to multiplex-PCR products once PCR success had been confirmed by gel electrophoresis, before pooling the barcoded PCR products and generating sequencing libraries. We used multiplex PCR products pooled from 24 individual samples as input for sequencing on MinION (Oxford Nanopore Technologies) Flongle flow cells, yielding 172,494-303,104 reads per run (after filtering of low quality <Q9, unmapped or reads outside of the accepted size range of 450-800bp) mapping to the targeted regions of the reference genome (NC_021508.1). This amounted to a median of 8,858 reads per sample (range 1,878-20,414), distributed across 73 individual amplicons, with median coverage per amplicon of 104 (range 0-612) across the three sequencing runs and 72 samples.

Using read mapping coverage for each Phy-con, we found that 4/72 samples performed poorly with a mean coverage of <50X across all amplicons, and these were excluded from further analysis (Supplementary Figure 8). Notably, these four samples were not outliers in terms of Treponema qPCR - indeed the majority of South African samples performed well, suggesting the upper sensitivity limit for successful sequencing of the TP-Phylo-Plex assay was at least Ct 32 (Supplementary Figure 8).

Analysing coverage for individual Phy-cons, as expected amplicon recovery was improved for samples with increased pathogen load (determined by qPCR Ct; Supplementary Figure 9). However, 15/74 Phy-cons in the scheme consistently yielded reduced coverage in multiplex PCR (but performed well as single-plex PCR assays). For the purposes of the scheme development, we removed these amplicons and retrospectively evaluated the impact on discriminatory power for sublineages (Supplementary Figure 4), henceforth focussing on the 59 remaining Phy-cons that worked consistently in multiplex. Note that this stringent approach was primarily for the purposes of scheme evaluation in the field, and further validation and primer balancing might have enabled retention of some Phy-cons. This would not have changed the way TP-Phylo-Plex was seen to work under a range of different conditions.

We next evaluated the sequencing accuracy of the Phy-cons generated for TP-Phylo-Plex on MinION. For this, we performed whole genome sequencing of the same 72 South African samples using the pooled sequence capture assay^3,15^ on Illumina NovaSeq to provide a gold standard comparator (Supplementary Table 3). We obtained high quality (≥75% reference sites at ≥5X coverage) Illumina consensus genomes for 59 of the 68/72 samples which also had high Phy-con sequence coverage using MinION. Focusing on the regions of the whole genome represented by the Phy-cons, we directly compared the variant calls generated from these 59 samples by the two orthogonal sequencing methods, examining the 181 variable sites contained within the 59 Phy-cons for discordant results. Of these, we found perfect concordance for 179/181 variable sites across all samples. Of the two discrepant sites, the first (NC_021508.1 position 7002) affected 1/59 samples and was due to a deletion detected in the Illumina sequence (7003delG) compared to the reference and MinION sequences (7003G). On closer inspection, this was found to be a novel deletion present in both sets of sequencing reads, but was missed in the MinION sequencing because our pipeline does not call INDELS. The second discrepancy (NC_021508.1 position 916096) occurred in 2/59 samples, and this site was found to be variable across the dataset (reference C allele, samples were either C or T). The two discrepant samples contained the ‘T’ variant allele in the Illumina data, but both had very low sequencing coverage in the relevant MinION sequencing amplicon and SNP loci (one mixed SNP with 2/3 reads supporting ‘T’; one clear ‘T’ allele supported by 6 reads). In both samples, this discrepancy with the Illumina data was caused by insufficient read coverage from the MinION sequencing, leading to the variant caller failing to correctly call the SNP. Given that we would not want a SNP to be inferred without sufficient coverage, this is intended behaviour. To improve reproducibility and accessibility, we reimplemented the bioinformatics pipeline using NextFlow^25^, and this included introducing additional quality steps around coverage that were designed to account for these issues in future, including ensuring reference sites with insufficient coverage in MinION data were called as ‘N’ in the final inferred sequence.

### Recapitulation of *T. pallidum* population structures by TP-Phylo-Plex

To assess the accuracy of phylogenetic delineation, we extracted the SNPs present within the final 59-amplicon scheme from a whole genome alignment and compared the resulting phylogeny to the one derived from the whole genomes (Figure 3). The distance matrices were correlated (Mantel statistic r=0.363, p<0.0001, 10,000 permutations), and we used Treespace^26^ to demonstrate the trees were highly concordant with sublineage (concordance=0.900) compared to whole genome tree bootstraps (median concordance=0.337, 100 bootstraps) and whole genome tip-randomised trees (median concordance=0.188, 100 permutations) (Supplementary Figure 10).

**Figure 3.**
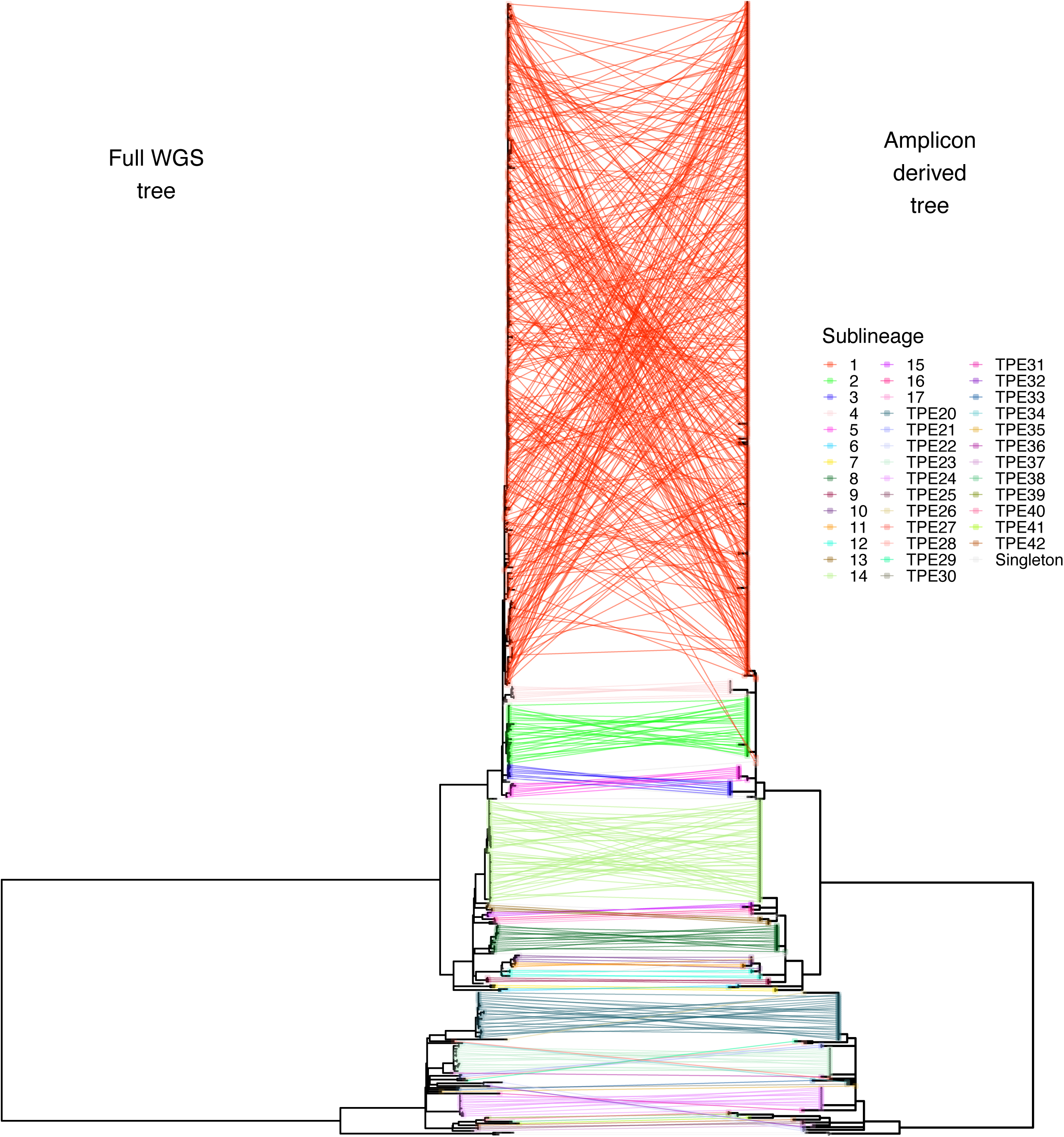
Recapitulation of whole genome population structure using Phylo-Plex. Tanglegram comparing whole genome phylogeny with phylogeny calculated from the *in silico* predicted amplicons. Broad sublineage clustering is replicated in the vast majority of cases. Note, tree scales are not identical, since the branch lengths in the amplicon-derived tree were extended to illustrate differences; the underlying topologies were not changed.

Next, we evaluated TP-Phylo-Plex’s ability to discriminate the 40 sublineages used in the original design. *In silico* evaluation of the original 73 amplicon design successfully delineated all 40 sublineages. However, the removal of 15 poorly performing amplicons resulted in a 59- Phy-con scheme which did not include highly discriminatory SNPs for 8 sublineages. Since Phy-cons can contain additional SNPs which have F_ST_ <0.95 that may still discriminate profiles, we examined the unique amplicon profiles predicted for each sublineage from the original WGS dataset. We found two instances where two sublineages (sublineages 10 and 11; sublineages 39 and 40) could not be discriminated from each other and thus TP-Phylo-Plex could not distinguish within these two groups using the 59- Phy-con scheme (whilst still being clearly delineated from other sublineages) (Supplementary Figure 11). Notably, those sublineages occurred in clonal lineages such as TPA SS14-Lineage, which had the smallest number of available discriminatory sites even at the WGS level, and were thus most affected by removal of amplicons. Because the level of discrimination can be ‘tuned’ in Phylo-Plex, users can make decisions about the epidemiological value of discriminating between those sublineages in their analyses.

We further evaluated the ability of TP-Phylo-Plex to discriminate previously described population structures, using a previously published whole genome phylogeny of 237 TPA genomes from the UK^27^ (Supplementary Figure 12). The dataset used for designing the scheme incorporated many of the same genomes and targeted the same sublineages, and TP-Phylo-Plex effectively recapitulated the population structure of UK syphilis.

### Application to more complex pathogen populations

For initial development, we used *T. pallidum* genomes which are highly genetically conserved, with sparsely distributed SNPs across the core genome, limiting the available genomic sites which can be used for discrimination. In populations of pathogens with more diverse genomes, there would be more available SNPs and these may be more densely distributed, enabling simpler design with shorter clustering distances. To evaluate the method with a more diverse population structure we selected the highly diverse sexually transmitted bacteria, *Neisseria gonorrohoeae*, using a large and recent genomic epidemiology study^28^ comprising 5578 genomes collected in Victoria, Australia. Taouk and colleagues used core gene MLST to identify 1262 transmission pairs or clusters^28^, and we selected all clusters comprising 10 or more genomes (n=79) for tracking. We reanalysed the sequencing data, inferring discriminatory SNPs for the 79 transmission clusters (8,692/88,6771 SNPs had F_ST_ ≥0.95). We examined the optimal positional distance between SNPs for clustering, and found that this was lower than for *T. pallidum* (Supplementary Figure 13). However, for the purposes of this evaluation we used the same settings as previously, clustering SNPs within 300 bp to form a positional network, resulting in 849 multi-SNP Phy-cons and 965 singleton SNPs. We excluded Phy-cons smaller than 700 bp, and performed hierarchical Phy-con selection to find the optimal combination of Phy-cons as previously, resulting in a final scheme comprising 169 Phy-cons and 785 discriminatory SNPs (Supplementary Figure 14, Supplementary Table 6). Notably, the computational resources required to repeat the analysis with the more complex dataset were substantially increased (requiring up to 128 Gb memory for some processes). This *in silico* scheme showed discriminatory support for 76/79 transmission clusters (Supplementary Figure 14) with clear phylogenetic discrimination (Supplementary Figure 15).

### Application in a low-resource setting

To determine the practical and operational feasibility of setting-up and using Phylo-Plex in a low resource setting, we set-up TP-Phylo-Plex at the Biomedical Research and Training Institute (BRTI; Harare, Zimbabwe), a small, ISO 15189 accredited service laboratory based in a converted townhouse. Equipment and molecular biology and sequencing reagents were shipped to Zimbabwe from the UK; key learning points here were around delays due to obtaining import permits. We implemented DNA extraction, qPCR diagnostics, multiplex PCR, library prep and sequencing on the MinION Flongle device (Oxford Nanopore Technologies) (Figure 4). We also performed full bioinformatic analysis using a Nextflow pipeline developed for processing Phy-con data, and real-time interpretation using an easy-to-use custom web app developed in Shiny (see Methods). We established realistic timelines for performing the assay in this setting, and were able to generate amplicons, perform sequencing and bioinformatics in under two days (Figure 4, Supplementary Table 4). As part of this process, two local PhD students were trained in all laboratory processes. Notably, power cuts occurred daily across Harare, but the BRTI laboratory had backup solar power cells. The backup system did malfunction, preventing PCR for two days. This highlights the need for robust power supply systems or qPCR machines that can run from batteries when using the method in a low resource setting.

**Figure 4.**
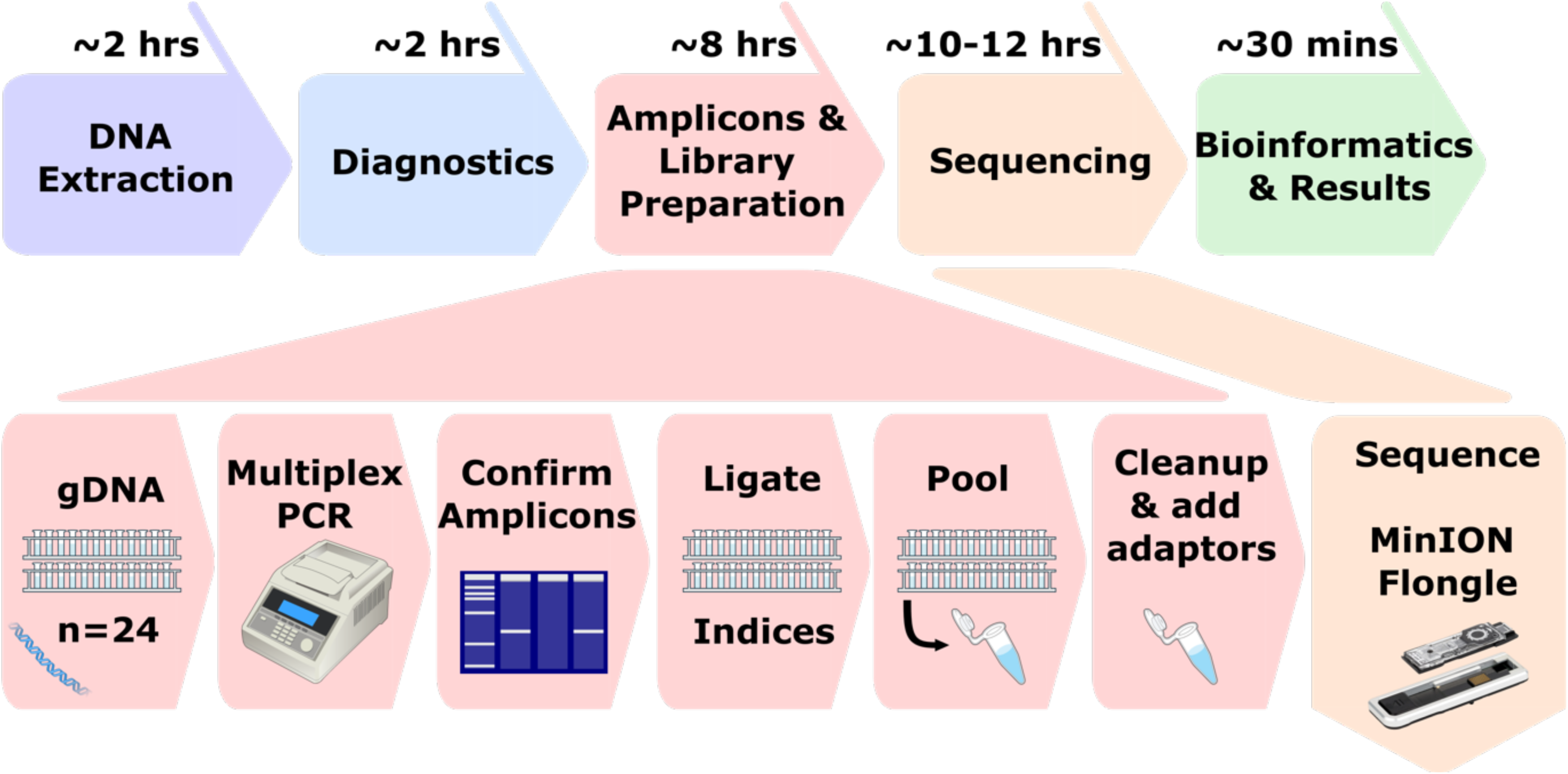
Workflow and timelines for Phylo-Plex protocol. DNA is extracted from clinical swabs, before molecular diagnostics (multiplex qPCR). Confirmed PCR positive samples proceed to Phylo-Plex multiplex PCR and library preparation, before sequencing on MinION Flongle flowcells. Bioinformatic analysis is automated using a single command Nextflow pipeline and interpretation is performed using a web-based Shiny App.

For this field evaluation, we prospectively recruited 100 individuals with genital ulcers in Harare, Zimbabwe, as part of an ongoing multi-country research study investigating the aetiology of genital ulcers. Of 100 genital ulcer swabs collected, 14 were qPCR positive for *T. pallidum* (qPCR Cts range 21.9 – 35.3). We used the 59 Phy-con TP-Phylo-Plex assay to sequence the 14 samples (including different types of technical replicates: independent DNA extractions of swabs taken from the same patient lesions, samples 454a and 454b, samples mz446a/mz446c and mz446b; independent PCRs performed on the same DNA extraction, samples mz446a and mz446c, samples mz476a and mz476b; Total 18), and recovered 12 complete TP-Phylo-Plex profiles. The technical replicates for sample mz476 had identical SNP calls, whilst although both of the replicates for sample mz454 successfully sequenced, one did not pass analytical quality checks. Three samples (5 replicates) with very high qPCR Ct >34 all failed to amplify at sufficient depth) (Supplementary Figure 16). This is consistent with our earlier observation that samples from South Africa performed well up to a qPCR Ct of 32, and suggests an approximate limit for successful sequencing between Ct 32 and 34. These samples yielded novel SNP profiles, reflecting that *T. pallidum* sequencing from Zimbabwe (as with the rest of Africa) was sparse prior to this work.

### Phylo-Plex offers a low-cost alternative for genomic surveillance

We evaluated the cost of implementing the 59-amplicon TP-Phylo-Plex, including all reagents (purchased in the UK using publicly displayed list prices) and consumables required for sequencing of PCR-positive DNA extracts. Excluding one-time setup costs (such as purchase of a sequencer and high-performance laptop), and assuming samples were sequenced in batches of 24, we estimated the cost to be £12.47/sample (Table 1), in addition to existing costs for DNA extraction and quality control. Of this cost, the most expensive components were the sequencing (£64.32/Flongle flow cell, £2.68/sample) and the native sample barcoding (£108.00/24 samples, £4.50/sample) reagents.

**Table 1.** Cost estimate for Phylo-Plex assay. Costs estimated using UK list pricing (institutional discounting may reduce this), and based on reagent volumes and usage. Initial setup costs may require larger purchases including MinION sequencer, High-performance laptop, Qubit Fluorometer, Thermocycler. Microcentrifuge, although many laboratories equipped for molecular biology, including those in Lower- and Middle Income Countries, may already have much of the required infrastructure (sometimes as a legacy of SARS-C0V-2 sequencing).

| Stage | Reagents | per 24 | per sample |
| --- | --- | --- | --- |
| PCR Primers | PCR Primers | £4.32 | £0.18 |
| PCR | Q5 Hot Start High-Fidelity 2X Master Mix (100) | £11.76 | £0.49 |
| PCR | Agarose / loading dye / EtBr / ladder | £1.56 | £0.07 |
| Library | NEBNext Ultra II End Repair/dA-tailing module (96) | £35.28 | £1.47 |
| Library | NEB Blunt/TA Ligase Master Mix (250) | £44.40 | £1.85 |
| Library | NEBNext Quick Ligation Module (20) | £20.40 | £0.85 |
| Library | ONT Native Barcoding Kit 24 V14 | £108.00 | £4.50 |
| Library | Qubit dsDNA HS Assay Kit (500 rxns) | £1.48 | £0.06 |
| Sequencing | ONT Flongle Flow Cell | £64.32 | £2.68 |
| **Reagent Total (Assuming List Prices)** |  | **£291.52** | **£12.15** |
| **Plasticware** |  |  |  |
| PCR | PCR Tubes | £0.52 | £0.02 |
| Library | Qubit Assay Tubes | £1.96 | £0.08 |
| Library | Microfuge Tubes | £0.30 | £0.01 |
| All | Filter Tips | £5.00 | £0.21 |
| **Plasticware Total (Assuming List Prices)** |  | **£7.78** | **£0.32** |
| **Total Assay Cost** |  | **£299.30** | **£12.47** |

Future replacement of native barcoding by ligation with custom primers introduced during PCR would enable further cost savings, whilst sequencing in larger batches on higher capacity MinION flowcells (e.g. 96-384 plex) could reduce sequencing costs. Moreover, whilst the scheme used here implemented amplicons of approximately 700 bp, designing schemes with smaller amplicons (e.g. 300-400 bp) would allow for higher multiplexing of hundreds to thousands of samples on Illumina short-read sequencing platforms.

We compared the costs of our Phylo-Plex assay to that of traditional MLST, which typically uses 7 amplicons and requires 14 Sanger sequencing reads per sample, costing commercially £28–£112/sample (£2-£8/reaction). MLST can also be adapted for high-throughput sequencing, such as nanopore. One approach is to adapt the Phylo-Plex methods by using existing MLST primers in discrete PCRs for each gene which are pooled before library preparation and sequencing (Supplementary Table 5), resulting in an increased per-sample cost of PCR (7 PCR reactions at £3.89 compared to a single PCR reaction at £0.56). MLST would also require less sequencing depth (7 vs. 59 regions), allowing higher multiplexing per run. Since the MinION Flongle run cost is fixed at £64.32, sequencing cost per sample decreases with higher throughput. However, because library costs remain fixed and additional PCRs must be performed, applying an unaltered 7-gene MLST schemes to 24-plex samples would be more expensive (£15.32/sample) than TP-Phylo-Plex (£12.15/sample), and 7-gene MLST remains more costly even at 192-plex. Alternatively, if MLST PCR primers are redesigned to enable multiplex PCR, overall costs of a 7-gene MLST can be reduced to £11.99/sample at 24-plex and £9.64 at 192-plex.

### Future proofing the Phylo-Plex and the Phy-con design

Although Phylo-Plex was specifically optimised to recover known population structures and sublineages, our approach of retaining and using SNP information means that detection of novel SNPs and sublineages is theoretically possible, provided the genomic variants occur within the targeted amplicon regions. We originally used rPinecone to cluster *T. pallidum* sublineages on the basis of being within 10-SNPs from a common ancestral node^23^, meaning that individual genomes within the same sublineage can be separated by up to 20 SNPs. Within the TP-Phylo-Plex amplicons, representing approximately 3.6% of the genome, most samples from the same sublineage were identical or differed by a single SNP (Supplementary Figure 11). Detection of a novel sublineage using TP-Phylo-Plex would therefore require at least two novel SNPs occurring within the available Phy-cons. Notably, *T. pallidum* accumulates approximately one substitution per genome every 7 years^15^, and novel sublineages with sufficient SNPs divergence for detection using TP-Phylo-Plex would therefore also be epidemiologically distinct.

Next, to quantify the sensitivity of TP-Phylo-Plex to novel variation, we simulated vertically inherited (non-recombining) SNPs accumulating in the genome by introducing random *in silico* mutations to the 1.139 Mb *T. pallidum* genome at different frequencies (0.1%, 0.05%, 0.01%, 0.005%, 0.001%, 0.0005%), corresponding to defined SNPs/genome (1140, 570, 114, 57, 11, 6). We counted how many WGS mutations occurred within TP-Phylo-Plex amplicons, evaluating each mutational frequency over 250 simulations, and found that at 0.001% genomic sites mutated (11 SNPs/genome – similar to the discriminatory level within our sublineages), only 5.2% of simulations had ≥2 SNPs occurring within the 59-amplicon TP-Phylo-Plex scheme. However, this increased to 45.6% of simulations at 0.005% genome sites mutated (57 SNPs) and to 87.6% of simulations at 0.01% genomic sites (114 SNPs) (Supplementary Figure 17). Therefore, the probability of distinguishing a novel sublineage depends on the level of genetic divergence – substantially divergent novel sublineages separated by >100 SNPs have a high probability of being identified, whilst the probability is reduced with less divergent sublineages. This is similar to how a standard MLST scheme would detect a new sequence type, except Phylo-Plex captures a larger proportion of the genome, and therefore has greater opportunity to detect novelty.

Should novel diversity be identified (either through Phylo-Plex, or through more conventional WGS surveillance), a further consideration is how straightforward it would be to enhance detection of novel diversity through adding additional amplicons. Notably, PCR primers represent a very small fraction of total assay costs – including a small number of additional primers would have minimal cost implications. However, since the primers are designed to work together in a multiplex, new primers would need to be validated *in silico* to ensure they were compatible with existing primers, as well as *in vitro* to ensure the multiplex PCR still functioned correctly. A simpler and more flexible solution would be to add a second more locally or regionally targeted multiplex PCR including new Phy-cons. This same approach could also be used for tracking non-chromosomal elements such as plasmids or AMR genes. The PCR products of both multiplexes could be combined before library preparation, at an estimated additional cost of £0.67/sample (Table 1), largely driven by the reagent costs of performing a second PCR. Alternatively, these secondary primer sets could simply be deployed on samples showing variation in the more general Phylo-Plex amplicon set.

## Discussion

As genomic surveillance becomes routine in high income countries, there is a need to maximise the utility of global microbial whole genome data we generate as a community so it can be used for epidemiological tracking in all settings. As a consequence of the international SARS-CoV-2 response, multiplex amplicon sequencing is now a proven technology widely used by laboratories around the world^11^. We sought to build on this capacity by developing a novel approach which enables low-cost and adaptable sequence-based pathogen surveillance in all settings, including those that are resource poor^29^.

Genome sequencing of ‘difficult to cultivate pathogens’ such as *Treponema pallidum* and some other STIs represents an ongoing technical challenge. Approaches for direct sequencing from clinical specimens include metagenomics^30^, and this can be supplemented with methods for host depletion through differential lysis or differential methylation patterns^31^, enrichment of on-target reads through adaptive sequencing^32^, selective multiple displacement amplification^33^, and sequence capture using baits^34,35^. Each of these offers varying degrees of enrichment, with sequence capture often being preferred for enriching low abundance target organisms^35^.

In the UK, Illumina sequencing of bacterial genomes is offered commercially in the region of £65/sample, and typically makes use of high throughput sequencers to reduce per-sample costs, requiring large batches of samples to make them affordable for pathogen sequencing. This can impact the timeliness of results whilst awaiting sufficient samples to complete a batch. Typing methods such as MLST are sometimes perceived as a cheaper alternative, but a typical MLST scheme uses 7 amplicons and requires 14 Sanger sequencing reads/sample, available at £2-£8/reaction commercially. Therefore, traditional MLST costs £28-£112/sample. By comparison, our TP-Phylo-Plex protocol costs £12.47/sample, and still recapitulates the current global population structure derived from whole genomes with a high degree of precision. Phylo-Plex achieves laboratory cost savings through both multiplexing and by using high-throughput sequencing, and existing MLST schemes (whilst designed for different use cases) could similarly reduce costs in this manner, particularly if redesigned to work in multi-plex PCR. Moreover, Phylo-Plex can be run in small batches (<£300/24 sample run) and give results in 24-48 hours, but because it can use multiple sequencing technologies, it can be scaled to enable very high throughput (hundreds of samples/run) sequencing. This makes it ideal for deployment to investigate emerging outbreaks *in situ* as well as more long-term surveillance studies.

We chose *T. pallidum* as a model pathogen because it is extremely expensive and complicated to culture, syphilis is a major global health concern and there is still limited data on its global diversity and spread. Moreover, Phylo-Plex is particularly attractive for *T. pallidum* because of the need to use costly metagenomic methods to recover genomes, as well as low genetic diversity and minimal recombination which made design against a reference genome straightforward. To show that this approach was suitable for a range of pathogens, we also designed an *in silico* scheme for a large and genetically diverse collection of *N. gonorrhoeae*, enabling discrimination of fine scale transmission clusters. Phylo-Plex is also appropriate for low-cost genomic surveillance of viral, bacterial and fungal pathogens, and because of the robust and portable nature of the method, as well as the relative ease of laboratory training, is ideally suited and proven for use in low resource settings, enabling tracking of diseases of public health importance such as mpox, chlamydia, typhoid, and cholera.

Unlike traditional molecular typing methods, Phylo-Plex uses population genomics-informed, phylogenetically robust grouping of taxa within specie(s). Inspired by computational hierarchical SNP typing methods such as GenoTyphi^36^, we developed an approach that focuses on key ancestral SNPs for clustering genomes, but refined the concept using network approaches to reduce operational complexity and design a low-cost laboratory method suitable for all settings. This is a similar concept to that of Tau-typing^37^, which also identifies phylogenetically discriminatory marker regions, but here we focus on maximising resolution with the minimum amount of regions. Despite recovering only 3.6% of the total genome length, our TP-Phylo-Plex scheme showed high concordance with the whole genome phylogeny, and indeed showed higher concordance than bootstrapped datasets derived from the whole genome itself. Moreover, our laboratory validation showed high accuracy in SNP detection, supporting our use of nanopore sequencing for field deployment.

Phylo-Plex can be used both as a standalone surveillance tool, and also to supplement and extend whole genome sequencing approaches; for a given population, a subset of samples could undergo WGS to identify key lineages, followed by design of a Phylo-Plex scheme to enable scaleup, and we describe important steps for how new schemes can be developed and evaluated before deployment. Existing Phylo-Plex schemes could also be rapidly adapted to a new outbreak simply by adding a separate pool of amplicons. Importantly, the sensitivity of a Phylo-Plex scheme can be adjusted – used a very stringent threshold to remove 15/74 amplicons from the TP-Phylo-Plex for operational reasons, yet this still enabled full delineation of 31/40 known sublineages. As with any multi-plex PCR, careful laboratory optimisation (including balancing primer concentrations, cycling conditions and modifying primers for compatibility) is required to achieve optimal results. Further optimisation might have allowed inclusion of these additional amplicons, thus enabling delineation of the remaining sublineages for the TPA scheme.

Phylo-Plex is capable of detecting novel lineages, albeit this is largely dependent on the level of nucleotide divergence between lineages. Adding more amplicons offers greater resolution and can enable antimicrobial resistance or virulence genotyping, at minimal additional cost, but this must be balanced against the technical complexity and time to optimise. The sensitivity required can be judged according to whether a particular lineage or sublineage, detectable by WGS and circulating in relevant populations, is of sufficient clinical relevance to require discrimination from other lineages. A recent example would be the current global health emergency associated with mpox lineage 1b which may have distinct disease outcomes^38^, where being able to rapidly and cheaply discriminate the causal lineage in an outbreak can inform both public health management and patient treatment. Indeed, we argue this is the real value of Phylo-Plex – a method for phylogenetically delineating strains which can be tuned to any pathogen, population, or epidemiological question.

## Methods

### *In silico* methods and design

### Dataset preparation

We constructed a globally representative dataset of 607 publicly available *Treponema pallidum* genomes, including high quality genomes with ≥95% of reference positions at ≥5X coverage. The vast majority of these had been sequenced using the pooled sequence capture method on an Illumina sequencer^3^ (Supplementary Table 1). Sequencing reads were mapped to a common reference genome (SS14, NC_021508.1), and masked for previously described hypervariable, repetitive and recombining genes, as well as using Gubbins^20^ v2.4.1 to further identify and filter recombining regions as previously described^15^. From the resulting multiple sequence alignment, we then constructed a maximum likelihood phylogeny using IQ-Tree^21,22^ v1.6.12 with a GTR+R model and 1000 ultrafast bootstraps^39^. We also used IQ-Tree to generate 100 standard non-parametric bootstrap trees for evaluating tree concordance. We inferred phylogenetic sublineages using rPinecone^23^ v0.1.0 with a threshold of up to 10 SNPs from the common ancestral node.

### Identifying and refining discriminatory sites

We converted whole genome sequence alignments into Variant Call Format (VCF) using snp-sites^40^ v2.5.1, before importing the data into R using vcfR^41^ v1.15.0. We created binary matrices for each genome indicating membership of each sublineage. We also included comparisons for *T. pallidum* subspecies (*pallidum* and *pertenue*), as well as the subspecies *pallidum* major lineages Nichols and SS14. For each sublineage, we then tested each variable SNP for population segregation using Weir and Cockerham’s F_ST_, using the ‘wc’ command in hierfstat^42^ v0.5.11. We considered sites with F_ST_ ≥0.90 sufficient for downstream analysis. Since our analysis included sites masked to ‘N’, we identified and excluded sites where the ‘N’ could potentially be misrepresented as a discriminatory SNP.

To identify information-rich regions of the genome, we calculated pairwise positional distances between discriminatory SNPs. We then subset data to include distances below individual thresholds, testing 50 bp intervals from 50-1000 bp. For each set of distances, we formed edge networks using the network package v1.18.2 in R, and extracted network components using iGraph v2.0.3, counting the number of network components (genomic regions), as well as the number of discriminatory SNPs per region and size range of the regions. Note that this approach of linking individual SNPs by distance can result in chains of connected SNPs that span many thousands of bases. Increasing the size of the linkage interval results in more SNPs within a network component, but increasingly leads to longer amplicons. We selected 300 bp for linkage as optimal for *T. pallidum*, since this balanced network components and SNP count against region size.

### Primer design

For multiplex primer design, we used the command line version of PrimalScheme^10,11^ v1.3.2, using the option to supply multiple distinct alignments for combined primer design. Using the SS14 reference genome (NC_021508.1), we generated a representative pseudomolecule, where genomic positions in our genome collection that were variable in ≥0.5% of genomes were masked to ‘N’. We then extracted target regions into individual fasta files, with the target region itself masked to ‘N’, and flanked by 150bp unmasked sequence. We specified a target amplicon size of 684bp (selected due to the typical target region size and allowing for flanking regions) in PrimalScheme to ensure multiplex primer design was forced to occur around the masked target region. We used the standard GC content and specified predicted melting temperatures of 59– 63°C ^10^. All primers were evaluated *in silico* using primer BLAST^43^ (implemented in primerTree^44^ v1.0.6), as well as using *in silico* PCR (available at https://github.com/sanger-pathogens/sh16_scripts/blob/master/legacy/in_silico_pcr.py) against SPAdes^45^ assemblies of the genome data.

### Macrolide resistance primer design

Macrolide resistance in *T. pallidum* is mediated by point mutations (A2058G, A2059G) in the ribosomal 23S sequences. Although published primers for *Treponema* 23S exist^46,47^, these were not optimised for compatibility with the multiplex scheme, necessitating design of new primers. To identify highly conserved *Treponema* specific regions in the 23S gene, which occurs universally across bacteria, we performed Compact Bit-Sliced Signature Index^48^ (COBS) search against a collection of ∼661,000 uniformly assembled bacterial genomes^49^. We used the *T. pallidum* ribosomal operon as a template, and generated 71 bp sliding windows along the *Treponema* operon as queries for COBS, examining identical (100%) search hits from the full bacterial dataset. Identical search hits included to representatives of other *Treponema* species (the dataset contained 174 assemblies classified as *Treponema* of which were 125 classified as *T. pallidum*) as well as a broad range of bacterial genera, with one window resulting in 229,791 identical hits (of which 51,460 were to *Streptococcus pneumoniae* and 48,124 were to *Staphylococcus aureus*). The median genome count for each window was 129 (reflecting hits to species in the Treponema genus), and regions of the *T. pallidum* 23S operon identified as matching ≥5% above the median (135.5) were initially flagged as potentially cross-genus matches, and this was confirmed by examining the species identities of the hits. Windows flagged for likely contamination were masked in the fasta file used as input to PrimalScheme for primer design using the conditions described for the full discriminatory scheme, resulting in two alternative primer designs which were screened using primer BLAST. After laboratory validation, the best performing of these two primer pairs was selected for use within the scheme.

### *Application to Neisseria gonorrhoeae* dataset

To evaluate the performance of the Phylo-Plex approach using a more complex dataset, we selected a large *Neisseria gonorrhoeae* dataset comprising 5880 genomes from Victoria, Australia^28^. In the original analysis, the authors defined 1177 putative transmission clusters based on core gene MLST alleles. For this evaluation, we aimed to design a Phylo-Plex scheme that could recover any transmission cluster comprising ≥10 genomes (n=79). We downloaded fastq files for 5878 publicly available sequences from the European Nucleotide Archive (ENA), and all reads were mapped to the *Neisseria gonorrhoeae* reference genome (FA1090, NC_002946.2) using snippy v4.6.0 (available at https://github.com/tseemann/snippy) with default settings to construct a reference mapped alignment. We extracted all SNPs using snp-sites and followed the workflow described above for identifying discriminatory SNPs for the 79 transmission clusters. Due to the complexity of the dataset, it was not possible to complete the analysis on a laptop due to excessive memory usage, and individual functions were reformatted to run as independent processes on a high-performance computer cluster. It was also not possible to run gubbins for exclusion of recombining sites on this dataset due to computational complexity. However, recombining regions that are exclusively discriminatory for a sublineage are potentially still useful within the Phylo-Plex framework. Moreover, recombination blocks are typically represented in our network as large components where nearly all SNPs support the same sublineage (Supplementary Figure 14), and the Phylo-Plex algorithm is designed to prioritise regions that support multiple sublineages. For phylogenetic analysis of such a large and complex dataset, we used IQ-Tree as described above with a GTR-Gamma model, but we excluded bootstrapping as this took over 30 days of CPU wall time.

### *In silico* validation

To evaluate the schemes, we initially simulated amplicons from Illumina-derived whole genomes. Based on the target regions, we extracted SNPs and constructed multiple sequence alignments using the vcfR^41^ v1.15.0, ape^50^ v5.8 and seqinr^51^ v4.2.36 packages, from which we inferred phylogenies using IQ-Tree^21^ v1.6.12. We compared the whole genome phylogenies to the inferred amplicon phylogenies for phylogenetic consistency using tanglegrams produced using ggtree^52^ v3.12.0, distance matrix comparison using Mantel tests in the vegan^53^ v2.6.6.1, as well as correlating whole genome derived sublineages with amplicon derived phylogenies using the treeConcordance function in Treespace^26^ v1.1.4.3. We examined distinct sublineage clustering *in silico* by constructing networks of identical amplicon profiles, and correlating these with WGS-derived sublineage assignments in R. Since *T. pallidum* genomes often contain uncertain sites, for this aspect we performed ancestral reconstruction of SNPs using pyjar v0.1 (available at https://github.com/simonrharris/pyjar) and extracted the pairwise cophenetic SNP distances from the ancestrally reconstructed phylogeny using ape. We subsequently performed direct comparisons on Illumina whole genome and MinION amplicon variant calls and phylogenetic placement derived from the same samples by integrating the variant calls from both methods (described below) using bcftools merge^54^ v1.19.

To evaluate the ability of Phylo-Plex to detect novel variation, we used a custom python script (randomly_mutate_genome.py, available at https://github.com/matbeale/TP-Phylo-Plex_paper_2025) to introduce random mutations at defined frequencies (0.1%, 0.05%, 0.01%, 0.005%, 0.001%, 0.0005%) into the SS14 reference genome (NC_021508.1). We simulated 250 different mutated genomes at each frequency, and converted the resulting fasta files to multi-sample variant call files (VCF) using snp-sites. We imported VCFs into R using vcfR and partitioned the SNPs for each simulated genome. For each set of variants, we calculated how many intersected with the amplicon positions using iRanges^55^ v2.38.0 and created summary statistics using tidyverse^56^ v2.0.0.

### Laboratory workflow

#### Validation samples

For initial validation of PCRs, we used ten-fold dilution series of high copy number rabbit passaged strains (SEA-83-1, BAL-2, BAL-6, Haiti-B). We also used a clinical dataset of 72 syphilis swab DNA samples derived from patients with genital ulcers in South Africa, which were sequenced using both Illumina (pooled sequence capture^3^) and the Phylo-Plex method. We also prospectively recruited 100 patients with genital ulcers in Zimbabwe as part of an ongoing study (“Multi-country Aetiology of Genital Ulcer Survey” – MAGUS); 14 *T. pallidum* PCR-positive swab DNA samples were sequenced using the protocol in a low-resource laboratory at the Biomedical Research and Training Institute in Harare, Zimbabwe. This included a combination of replicates for three samples; one sample (mz454) was independently extracted from two separate aliquots of swab lysate as technical replicates; one samples with high total DNA and Treponema qPCR load (mz476) was sequenced in duplicate from the same extract as technical replicates; one sample with low total DNA and Treponema qPCR load (mz446) was sequenced as both types of technical replicates..

### Sample Preparation

For the validation in Zimbabwe, ulcers were swabbed using flocked swabs in transport medium (Corning), with 1 ml lysate realiquoted into cryovials for storage at -80°C ahead of processing. DNA extraction was performed on 200µl swab lysate using the QIAamp DNA Mini Kit (Qiagen) according to the manufacturer’s instructions.

### PCR amplification

We modified the ARTIC network (https://artic.network/) SARS-CoV-2 sequencing protocol (available at https://www.protocols.io/view/artic-sars-cov-2-sequencing-protocol-v4-lsk114-bp2l6n26rgqe/v4) for multiplex PCR and library preparation, adjusting reaction volumes to reduce cost and increasing cycling times to improve PCR efficiency of 700 bp amplicons. Briefly, for each sample we prepared a PCR reaction mix containing 6.25µl Q5 Hot Start High-Fidelity 2X Master Mix (New England Biolabs), 2µl pooled multiplex primers (10 mM; Integrated DNA Technologies), 1.75µl nuclease-free water, and 2.5µl template DNA (total reaction volume of 12.5µl). Samples were initially denatured for 30 seconds at 98°C, followed by 35 cycles of 15 seconds at 98°C, 4 ½ minutes at 62°C and 30 seconds at 72°C. Performance of bulk PCR products was assessed using agarose gels, ensuring a band was present at 700-800 bp.

### Library preparation and sequencing

PCR products were used directly without cleanup, and diluted 1:5 (or less, if fewer than 24 samples/run) in nuclease-free water. We performed end repair using the NEBNext Ultra II End Prep module (New England Biolabs), combining 1.2 µl Ultra II End Prep buffer with 0.5µl End Prep Enzyme Mix, and adding 8.3µl of diluted PCR product, before incubation for 15 minutes at room temperature (∼20°C), then inactivation for 15 minutes at 65°C, then 1 minute on ice. PCR products were individually barcoded using 5µl Blunt/TA Ligase Master Mix (New England Biolabs), 1.25µl DNA barcode (SQK-NBD114.24, or SQK-NBD114.96; Oxford Nanopore Technologies), 3µl nuclease-free water, and 0.75µl end-repaired product from the previous step; ligation reactions were incubated for 20 minutes at room temperature, followed by 10 minutes at 65°C, then 1 minute on ice. All barcoded PCR products were then pooled into a single tube for onward preparation. We performed two rounds of SPRI bead cleanup (AmpureXP, Beckman-Coulter) at 0.4X, using the Short Fragment Buffer for washing (Oxford Nanopore Technologies). After cleanup and quantification, we ligated sequencing adaptors (Oxford Nanopore Technologies) by combining 10µl NEBNext Quick Ligation Reaction Buffer (5x), Quick T4 DNA Ligase (both New England Biolabs), 5µl Native Adaptor (Oxford Nanopore Technologies), and 30µl pooled library, followed by incubation for 20 minutes at room temperature. We then performed two further rounds of SPRI bead cleanup, before final quality control and library preparation.

Pooled multiplex amplicon libraries were sequenced in groups of 24 on a single run of the MinION Flongle device (Oxford Nanopore Technologies) using either R9.4.1 (initial validation and testing of South African samples) or R10.4.0 (field deployment and Zimbabwe samples) chemistry, with the change motivated by discontinuation of R9.4.1 reagents and flowcells. Where fewer than 24 samples were available (e.g. in Zimbabwe), we reduced the dilution factor of initial PCR products from 1:5 up to 1:3 to ensure sufficient total DNA was available for subsequent steps.

### Bioinformatic analyses

#### Basecalling, QC, Variant calling

Illumina data was processed and analysed as previously described^15^. Briefly, enriched metagenomic sequencing reads were screened using Kraken2^57^ v2.1.2 and *Treponema* reads were extracted using seqtk v1.3 (available at https://github.com/lh3/seqtk). We mapped sequencing reads to the SS14 reference (NC_021508.1) after masking previously described recombinogenic regions, called variants using bcftools^54^ v1.19 to construct a multiple sequence alignment, and used Gubbins^20^ v2.4.1 to identify and mask additional recombinant sites.

Initial validation of MinION amplicon data was conducted using a custom pipeline. Sequencing runs were basecalled and demultiplexed using the high accuracy model in Guppy v6.4.2. Sequencing reads with >Q9 were mapped to the SS14 reference (NC_021508.1) using minimap2^58^ v2.16, and quality metrics were produced using samtools^59^. In particular, we calculated coverage metrics for each individual amplicon by parsing the outputs of samtools depth v1.6. After mapping, we trimmed reads to remove primers using Cutadapt^60^ v1.15, before performing SNP calling using Clair3^61^ v1.0.1 (available at https://github.com/HKU-BAL/Clair3) using the “res_dna_r9.4.1_e8.1_hac_v033” model. We compared variants inferred from Nanopore data to Illumina data by merging filtered VCF files using bcftools^54^ v1.19.

After initial validation of methods, we subsequently reimplemented this workflow in a streamlined NextFlow^25^ pipeline (available at https://github.com/sanger-pathogens/long-read-ampliseq; workflow shown in Supplementary Figure 18). This replaced the basecaller with Dorado v0.5.1 (available at https://github.com/nanoporetech/dorado), incorporated additional quality steps, and refined variant calling and pseudosequence generation. Raw sequencing reads (fast5/pod5) were base-called using Dorado with the “dna_r10.4.1_e8.2_400bps_hac@v4.3.0” model and minimum quality score set to 9, and demultiplexed using the appropriate barcode kit name specified (e.g. ‘SQK-NBD114-24’). samtools^59^ v1.19.2 was used to convert the resulting bams to fastqs. Primers and low quality (below Q15 at each end) bases were trimmed from fastqs using Cutadapt^60^ v4.7, with lower read length cutoff set to 450 and upper read length cutoff set to 800 to ensure only reads of the expected length were included. Remaining bases with quality <15 were also masked using seqtk v1.4 (available at https://github.com/lh3/seqtk). Mapping of the reads against the masked SS14 reference was performed using Minimap2^58^ v2.26. Unmapped reads and alignments with mapping quality below 50 were filtered using samtools^59^. As each read should map to one of the target regions, alignments not overlapping with these regions or that are secondary or supplementary were also filtered. Variant calling was performed using Clair3^61^ v1.0.9 in haploid mode, using the “r1041_e82_400bps_hac_v430” model and specifying to only call SNPs with a minimum coverage of 5. Bcftools^54^ v1.20 was used to merge the genome variant call files (gvcfs) from all samples to a multi-sample vcf and also to convert to tab-delimited format for simple parsing. A custom script run in Python 3.10.12 (available in the pipeline) was used to create a consensus fasta file for each sample with individual target regions, as well as all the target regions concatenated, including variants and reference calls with minimum genotype quality of 1 and masking everything else. Individual target region fastas were also concatenated to make a multiple sequence alignment, from which SNPs were extracted with snp-sites^40^ v2.5.1. Throughout the pipeline, various tools are used for QC reporting at different stages, including PycoQC v2.5.2 (available at https://github.com/a-slide/pycoQC), FastQC^62^ v0.12.1 (available at https://github.com/s-andrews/FastQC), samtools, bedtools v2.31.1, as well as custom Python scripts. MultiQC^63^ v1.22.2 is used to create a summary report combining output from PycoQC, FastQC and samtools stats/flagstat for all samples. Typical run time for this pipeline on a high-performance laptop for a TP-Phylo-Plex MinION Flongle sequencing run was 20-40 minutes (depending on the number of reads generated by the sequencing). Fieldwork in Zimbabwe used https://github.com/sanger-pathogens/ONTAP v1.0.0 for analysis.

We also developed a Shiny web interface (“AmpliSeq_QC_Frontend.R”) in R which can be loaded using Rstudio, included with the pipeline code at https://github.com/sanger-pathogens/ONTAP, to enable straightforward guided interpretation of bioinformatics outputs for use by non-bioinformaticians in the field. This takes the directory path of a completed NextFlow pipeline run as input, and provides a point-and-click interface to assess quality and coverage metrics, allowing the user to select and deselect samples, and reanalysing results (including phylogenies and plots) in real-time.

### Evaluation of *in vitro* amplicon performance

To identify poorly performing amplicons amongst the South African validation set, we first identified and excluded samples which performed poorly across all amplicons based on an overall mean coverage of <50X. By doing this, we ensured the analysis was not substantially biased by sample. Amongst the samples which performed well overall, we then identified poorly performing amplicons by asking what proportion of amplicons met a minimum coverage threshold of 25X (using the quantile function in R). We then excluded amplicons where >10% of the samples had a mean coverage below 25X. For the TP-Phylo-Plex assay, this led to exclusion of 15/74 amplicons.

### Statistics and plotting

Phylogenies and tanglegrams were plotted using ggtree^52^ v3.12.0, and all other plots were produced using ggplot2^64^ v3.5.1 in R^65^ v4.4.2. Networks were plotted using ggnetwork^66^ v0.5.13 and iGraph^67^ v2.0.3, and all statistics were calculated in R. Code workflows were made in draw.io and Lucidchart. Additional diagrams were made in Inkscape. ChatGPT v3.5 was used for initial code scaffolding and to assist with troubleshooting during the development of the Shiny application for Nextflow data interpretation.

### Ethical approvals

Clinical samples from South Africa were collected as part of routine public health surveillance, and ethical approval for genome sequencing was granted by the University of the Witwatersrand Human Research Ethics Committee (Medical) (Ethics clearance certificate no. M230157). Clinical swabs from Zimbabwe were collected as part of the Multi-Country Aetiology of Genital Ulcer Study (MAGUS), and ethical approvals were granted for patient recruitment, diagnostics and genomics from the Medical Research Council of Zimbabwe (MRCZ/A/2878), the Biomedical Research and Training Institute Institutional Review Board (Ap/175/2022) and the London School of Hygiene and Tropical Medicine Ethics Committee (26731).

## Data Availability

European Nucleotide Archive (ENA) Accessions for Illumina sequencing reads from the initial dataset used for primer design are listed in Supplementary Table 1. Accessions for the Illumina reads from South African syphilis are listed in Supplementary Table 3 and are available at the ENA under project accession PRJEB60271. Oxford Nanopore sequencing reads are available from the ENA under project accession PRJEB85457, and are listed in Supplementary Table 3 (for South Africa) and Supplementary Table 4 (for Zimbabwe).
The code used in this paper (including for identifying discriminatory sites and the hierarchical selection algorithm) are available at https://github.com/matbeale/TP-Phylo-Plex_paper_2025. The Nextflow pipeline for automated data processing of Phylo-Plex data is available at https://github.com/sanger-pathogens/long-read-ampliseq.

https://github.com/matbeale/TP-Phylo-Plex_paper_2025

https://github.com/sanger-pathogens/ONTAP

## Data and Code Availability

European Nucleotide Archive (ENA) Accessions for Illumina sequencing reads from the initial dataset used for primer design are listed in Supplementary Table 1. Accessions for the Illumina reads from South African syphilis are listed in Supplementary Table 3 and are available at the ENA under project accession PRJEB60271. Oxford Nanopore sequencing reads are available from the ENA under project accession PRJEB85457, and are listed in Supplementary Table 3 (for South Africa) and Supplementary Table 4 (for Zimbabwe).

The code used in this paper (including for identifying discriminatory sites and the hierarchical selection algorithm) are available at https://github.com/matbeale/TP-Phylo-Plex_paper_2025 and at https://doi.org/10.5281/zenodo.14894111. The Nextflow pipeline for automated data processing of Phylo-Plex data is available at https://github.com/sanger-pathogens/ONTAP.

## Author Contributions

Conceptualisation: MAB; Methodology: MAB, VS, KEA, GL, SD, WRS, BS, FL, MJD; Formal Analysis: MAB; Investigation: MAB, VS, KEA, GL, MC, TMW; Resources: MAB, SD, WRS, BS, FL, MPM, JMEV, BDCD, EM, LR, BM, MM, NRT; Data Curation: MAB, VS; Writing – Original Draft: MAB; Visualisation: MAB; Supervision: LR, RAF, NRT; Project Administration: MAB, MM, BM, ED, EM, RAF, NRT.

## Acknowledgements

We thank the teams and administrators at the National Institute for Communicable Disease in South Africa and at the Zvitambo and Biomedical Research and Training Institute labs in Harare, Zimbabwe, as well as all partners involved in the MAGUS study. We acknowledge the Core Sequencing, Pathogen Informatics and Samples teams at the Wellcome Sanger Institute.

This work was part-funded by a Gates Foundation grant (INV-035896) to MAB, MM, RAF and NRT. MAB, VS, KEA, GL, SD, WRS, BS, FL, MJD and NRT were supported by Wellcome funding to the Sanger Institute (206545/Z/17/Z).

This work was supported, in whole or in part, by the Gates Foundation [INV-035896] and the Wellcome Trust [206545/Z/17/Z]. The conclusions and opinions expressed in this work are those of the authors alone and shall not be attributed to the Gates Foundation or the Wellcome Trust. Under the grant conditions of the Foundation, a Creative Commons Attribution 4.0 License has already been assigned to the Author Accepted Manuscript version that might arise from this submission. Please note works submitted as a preprint have not undergone a peer review process.

---

**Supplementary Figure 1.**
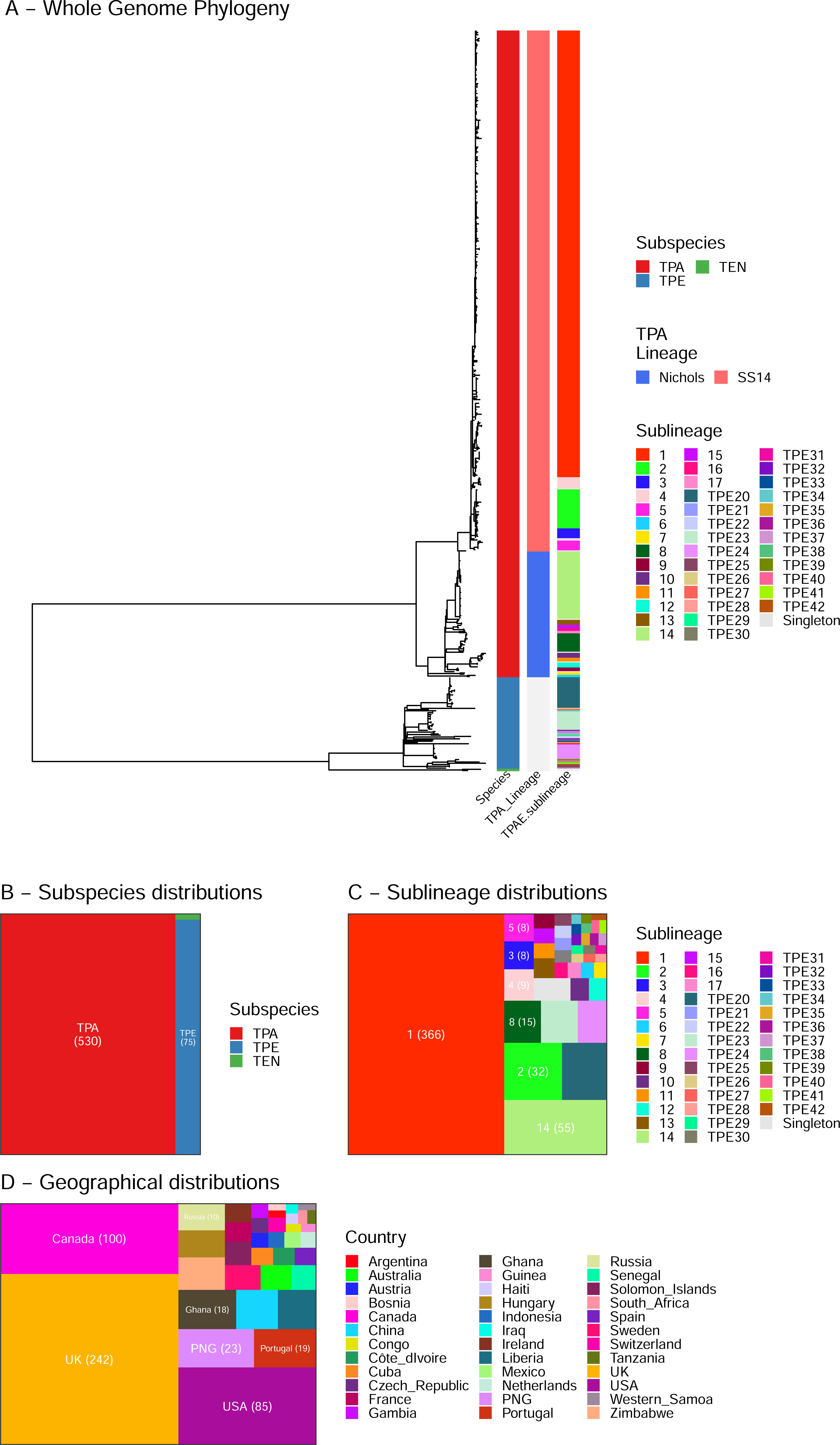
Population structure and characteristics of initial *Treponema* dataset used for Phylo-Plex design. A – Maximum Likelihood phylogeny of 607 *Treponema pallidum* genomes, with coloured tracks showing Subspecies, Lineage (within subspecies pallidum), and Sublineage (across all subspecies). B - Treemap plot showing distribution of genomes by *T. pallidum* subspecies: TPA – subspecies *pallidum*, TPE – subspecies *pertenue*, TEN – subspecies *endemicum*. C - Treemap plot showing distribution of genomes by T. pallidum subspecies (clustered as samples sharing a common ancestral node ≤10 SNPs. D - Treemap plot showing distribution of genomes by country.

**Supplementary Figure 2.**
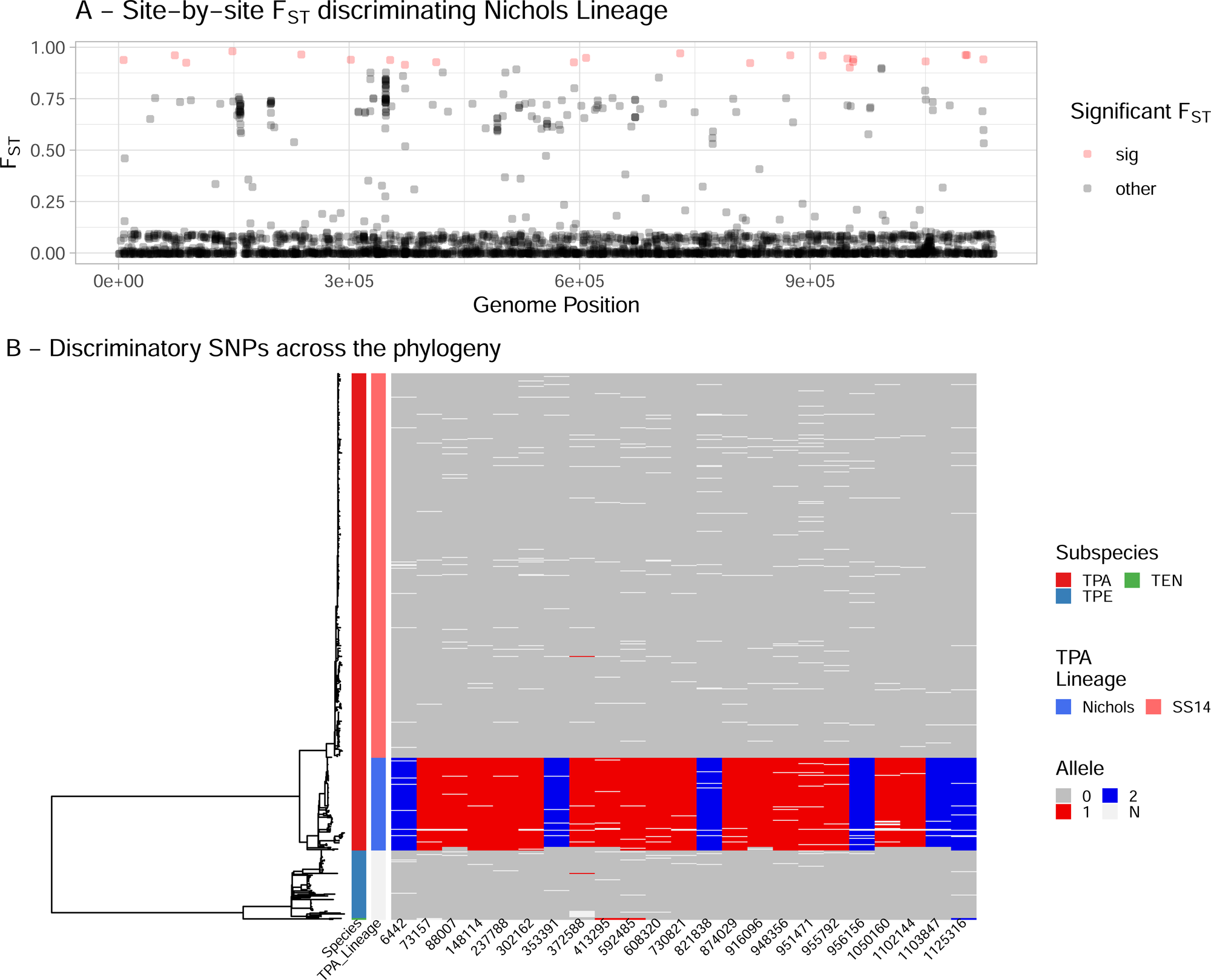
Identifying discriminatory SNPs for a single lineage. A – Population fixation analysis (F_ST_) of variable sites. Sites in red discriminate *T. pallidum* Nichols Lineage from all other lineages at F_ST_ ≥0.9. B – Maximum likelihood whole genome phylogeny showing allelic identity at each discriminatory site identified in A. Colours indicate allele detected (pale grey indicates ‘N’, where data was missing – common in metagenomic data).

**Supplementary Figure 3.**
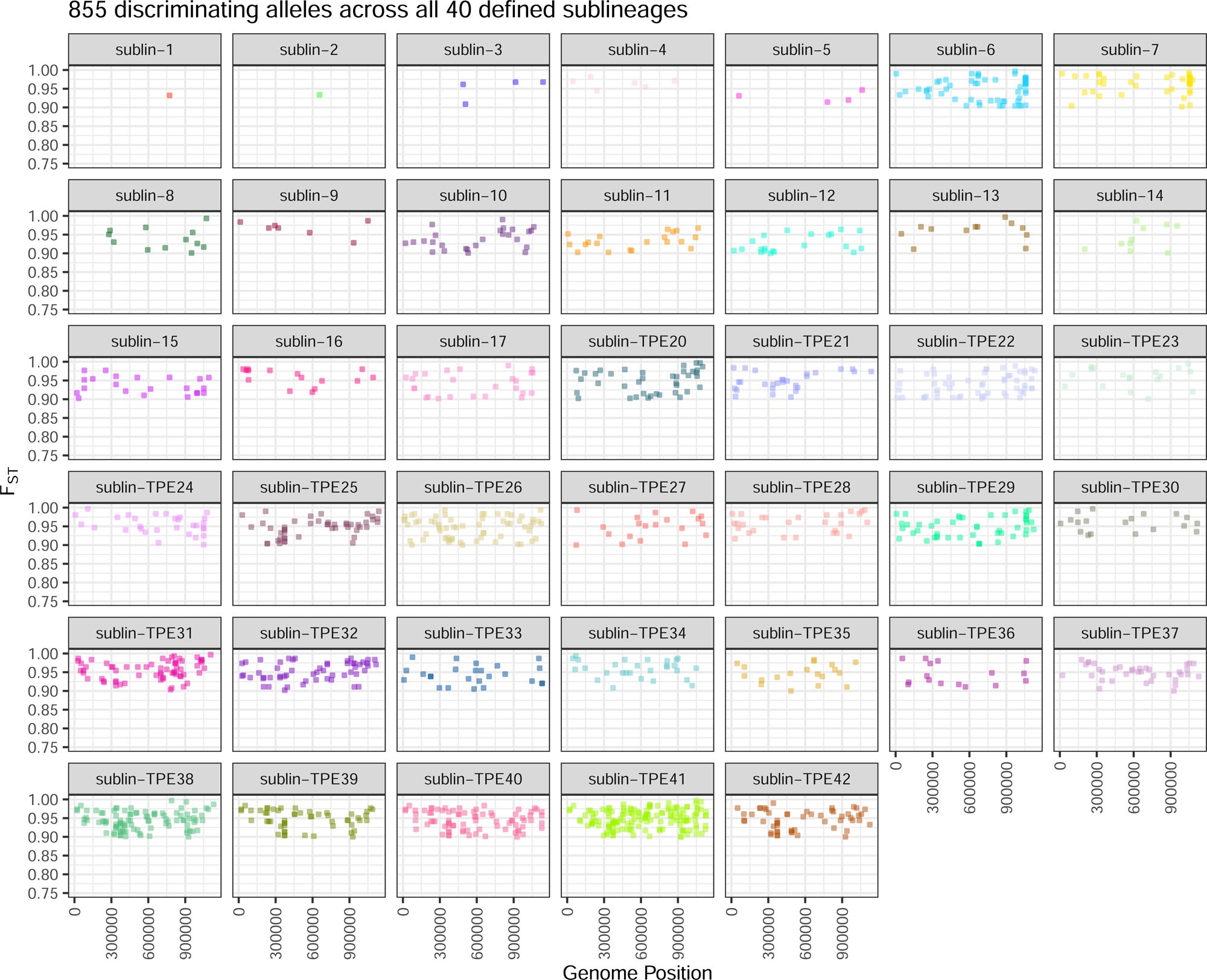
Identifying discriminatory SNPs for 40 sublineages. Population fixation analysis (F_ST_) of variable sites for each sublineage - plots show only sites with F_ST_ ≥0.9.

**Supplementary Figure 4.**
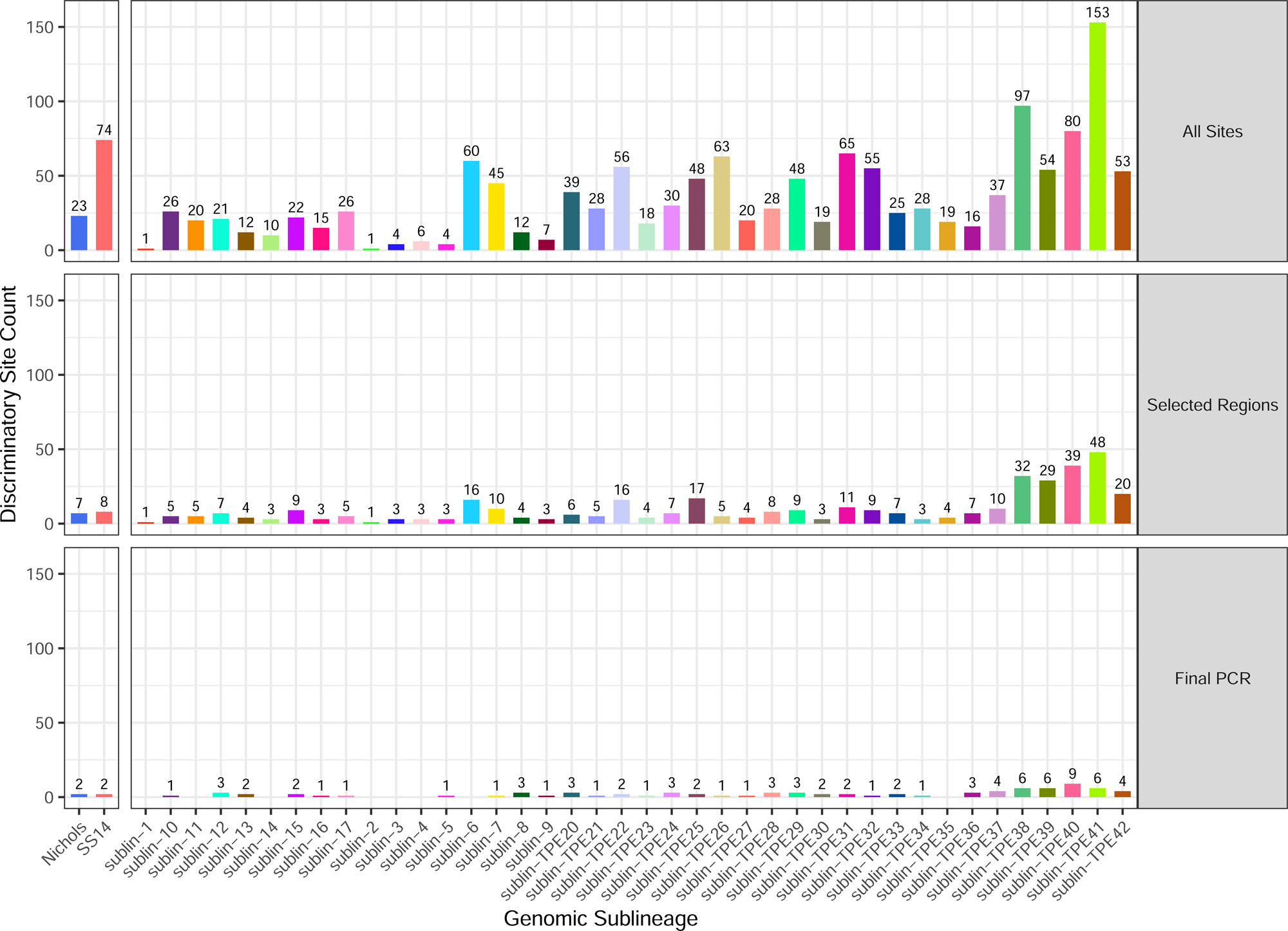
Discriminatory site support for each sublineage. Plot shows number of discriminatory sites specifically supporting each lineage and sublineage when considering (i) all discriminatory sites identified in the genome dataset, (ii) sites included in 74 regions selected by the Phylo-Plex selection algorithm, (iii) sites remaining after full optimization of the 59-amplicon multiplex PCR. The final PCR used for evaluation removed 15 amplicons and therefore removed direct support for 8 sublineages (‘*’), but future iterations would modify the primer designs and balance to reinclude these.

**Supplementary Figure 5.**
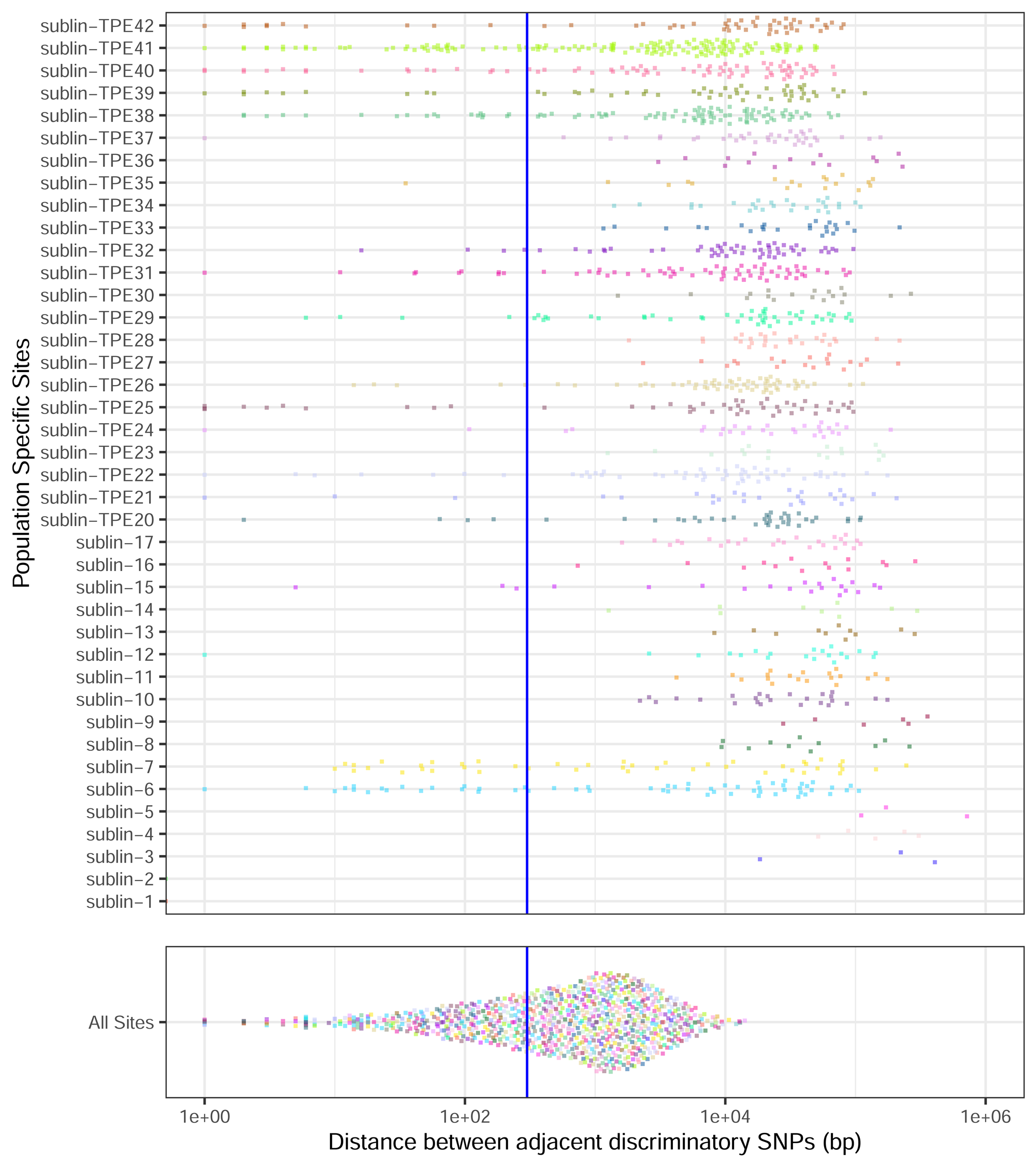
Genomic distance between individual sites is reduced when considered as a total population. Genetic distance between adjacent discriminatory SNPs according to sublineage and as a total population. Blue line highlights 300 bp.

**Supplementary Figure 6.**
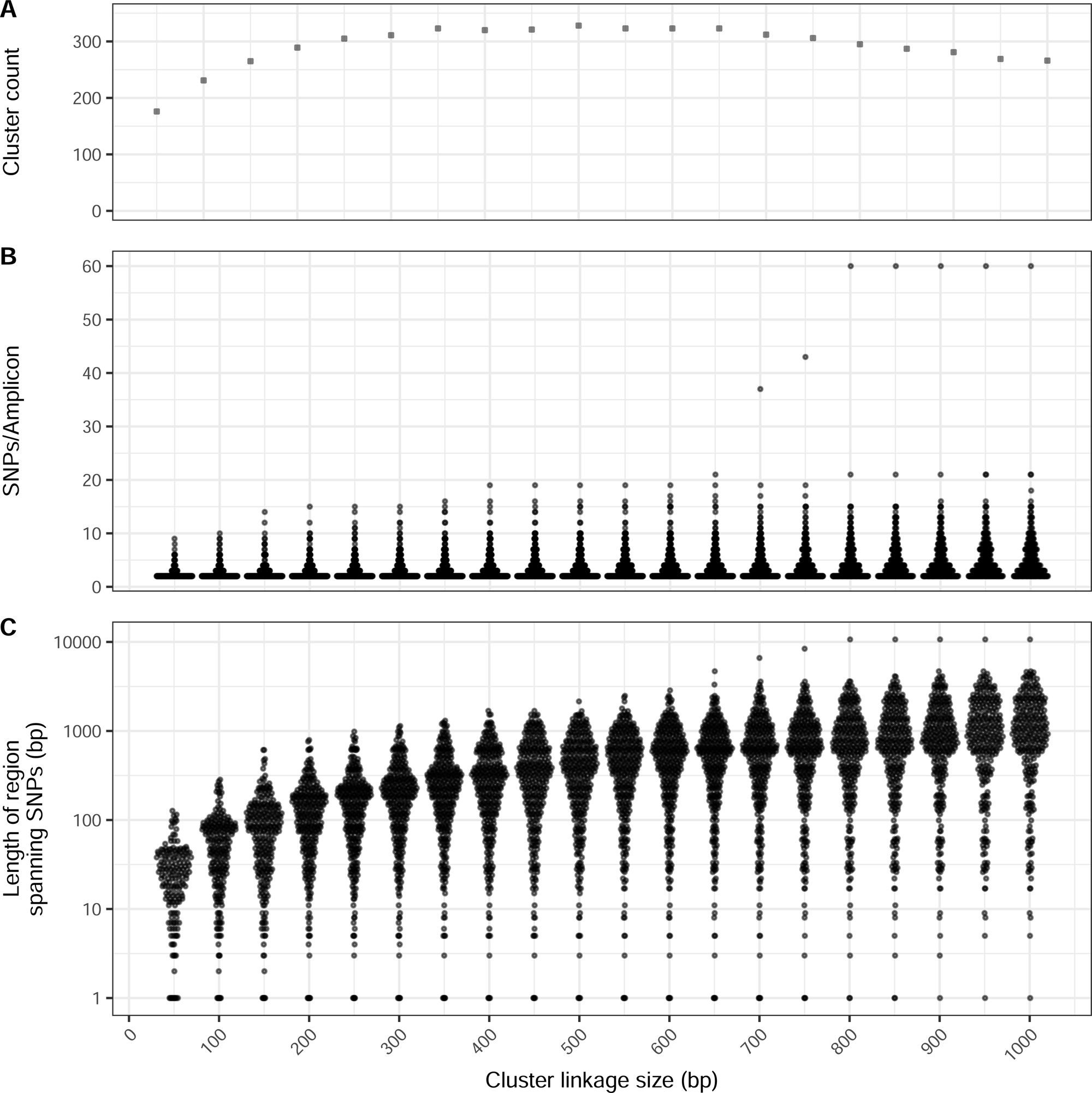
Changing distance used to link SNPs as clusters impacts cluster count, SNPs per amplicon and minimum amplicon size. The selection of an appropriate genomic distance for positional clustering can be tuned and impacts the number of SNPs that merge into clusters. However, increasing cluster distance between individual SNPs can also substantially impact the total length of SNP networks, resulting in candidate amplicons too long to reasonably amplify in multiplex PCR. A – Number of positional clusters produced with different genomic distance between sites. B – Number of SNPs present in each cluster (candidate amplicon) produced with different genomic distance between sites. C – Genomic distance between furthest SNPs in cluster (i.e. minimum length of candidate amplicon) produced with different genomic distance between sites.

**Supplementary Figure 7.**
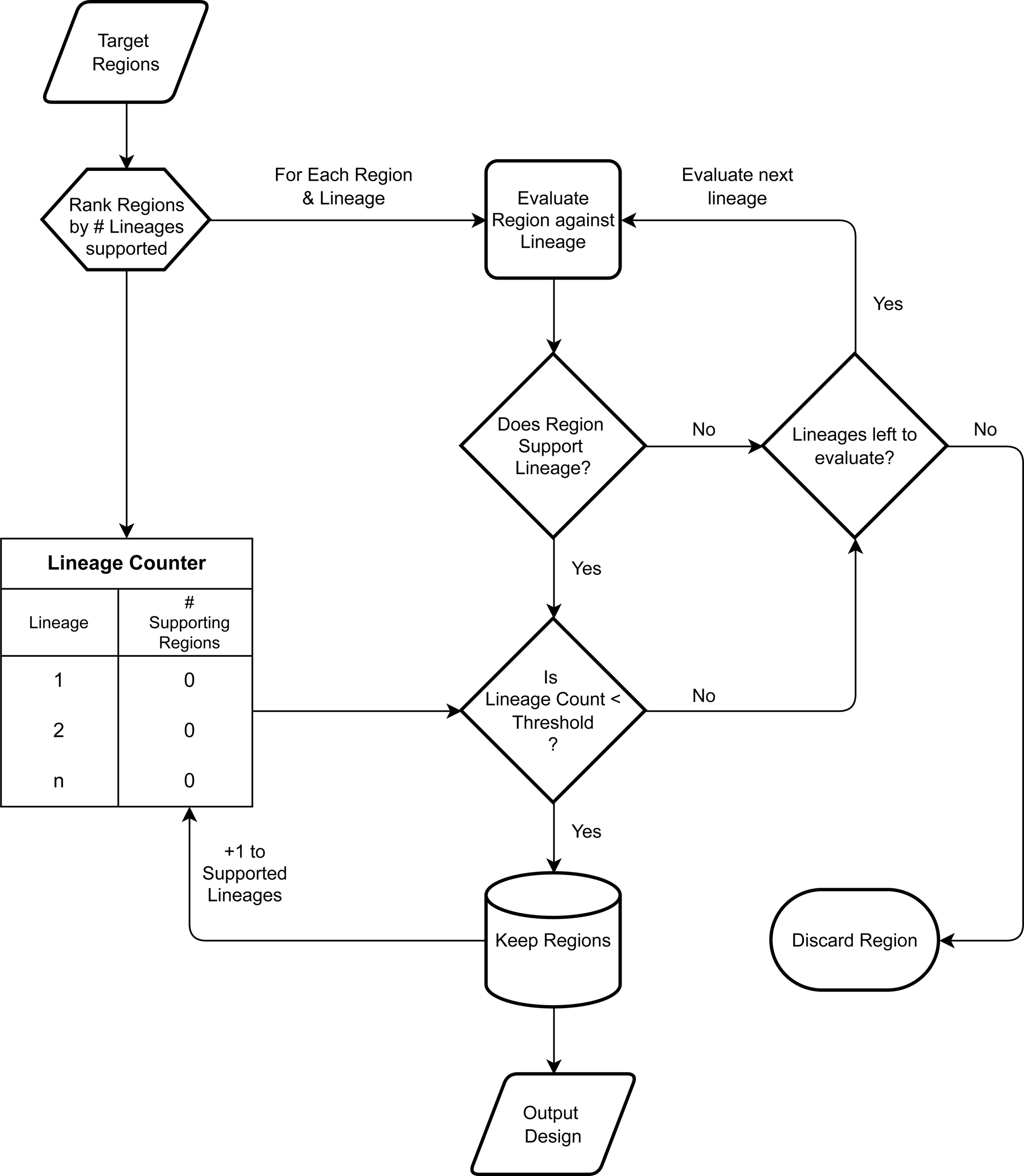
Hierarchical selection algorithm for maximising discriminatory power whilst minimising total number of amplicons. Each candidate region is evaluated based on the sublineages the SNPs within it support, and regions are added until support for a sublineage meets a minimum threshold (3 SNPs), after which, support for that sublineage is no longer considered.

**Supplementary Figure 8.**
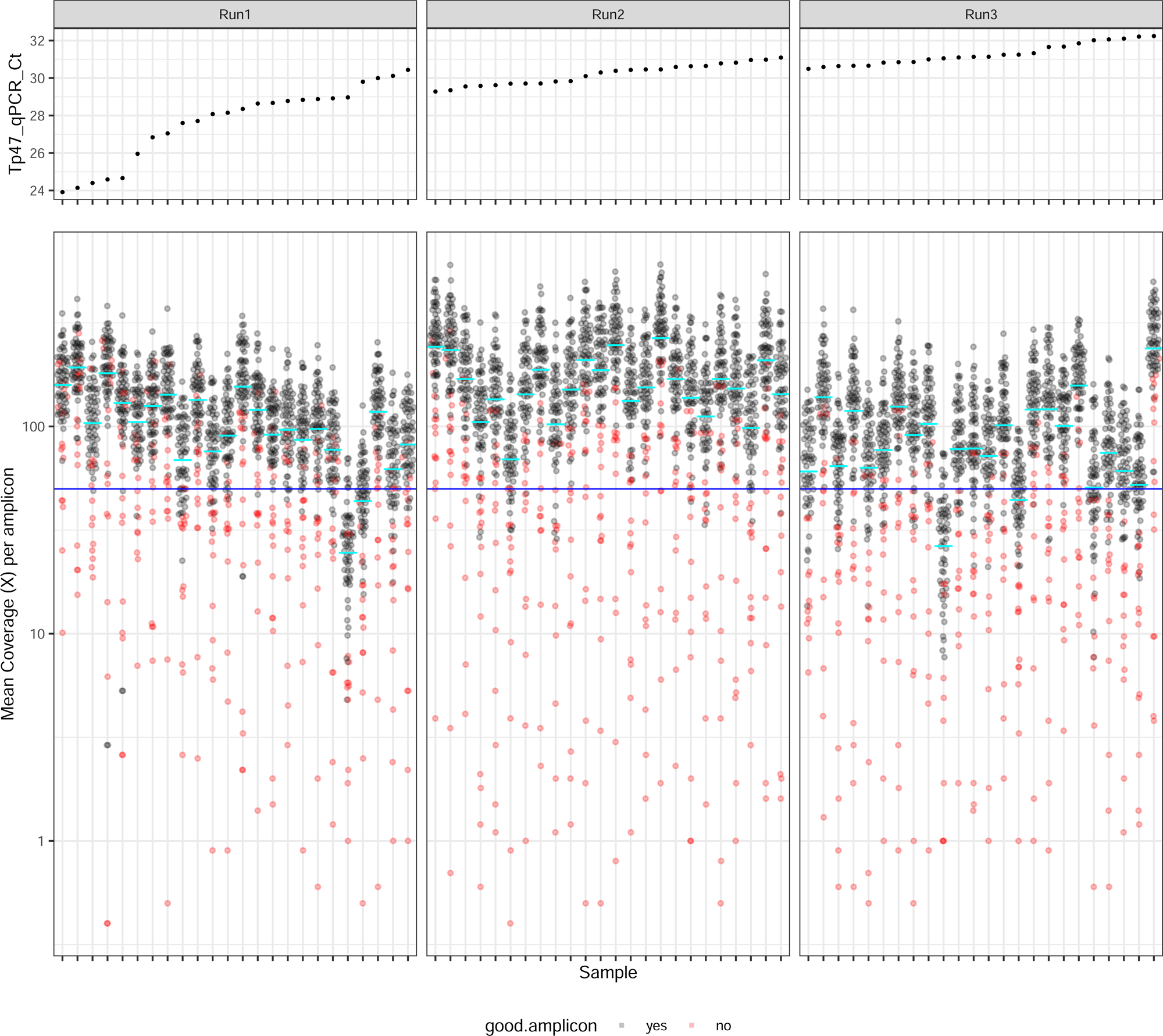
Sequencing performance of 72 South African clinical syphilis samples. Points show mean coverage for 74 amplicons from each of 72 samples, ordered by input Treponema qPCR Ct. Samples were sequenced on three separate runs of MinION Flongle cells. Blue line indicates 50X coverage. Red points indicate 15 amplicons which consistently performed poorly (see Supplementary Figure 9). Cyan lines indicate mean coverage of all amplicons in that sample.

**Supplementary Figure 9.**
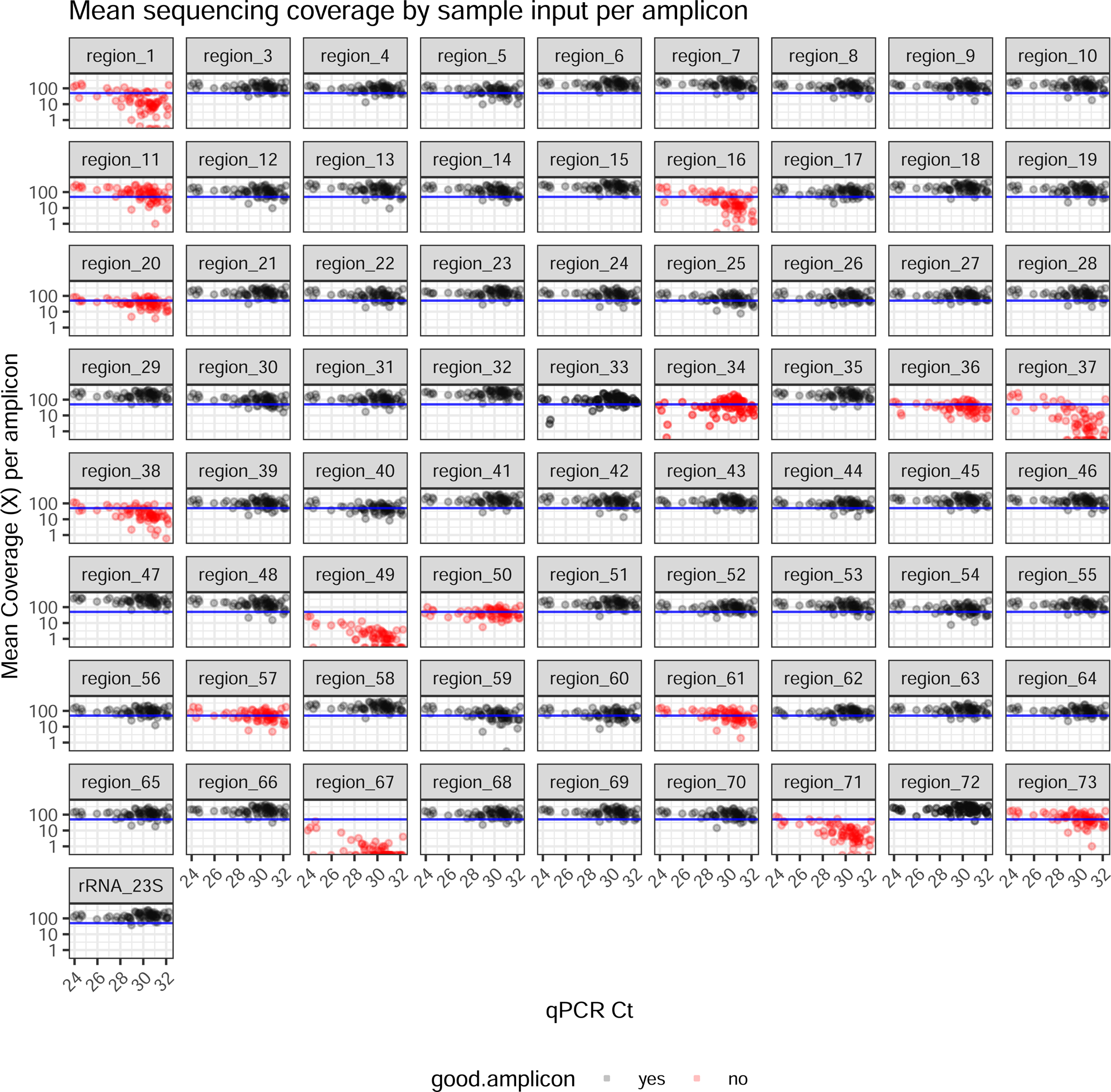
Sequencing performance of 74 multiplex amplicons in clinical syphilis samples. Points show mean coverage for 74 amplicons from each of 72 South African clinical syphilis samples compared to input Treponema qPCR Ct. Blue line indicates 50X coverage. Red points indicate 15 amplicons where ≥10% of samples had <25X coverage (after excluding samples which performed poorly overall).

**Supplementary Figure 10.**
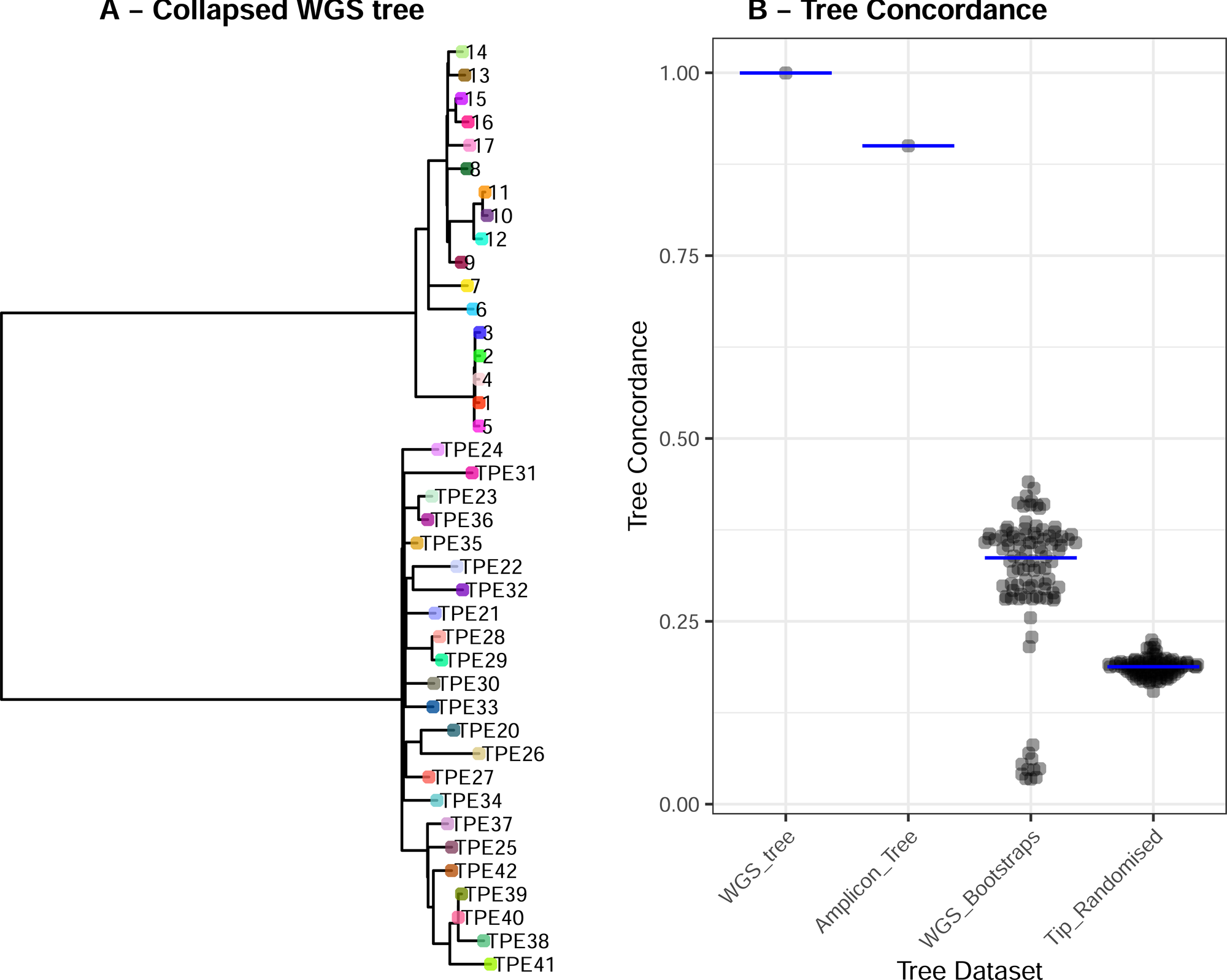
Quantitative comparison between WGS derived phylogeny and Amplicon derived phylogeny demonstrates high concordance. A – Collapsed WGS phylogeny (one tip per sublineage), B – Tree concordance between collapsed WGS tree and (i) full WGS tree, (ii) Amplicon derived tree (simulated), (iii) 100 bootstraps derived from full WGS tree, (iv) 100 tip-randomised full WGS trees.

**Supplementary Figure 11.**
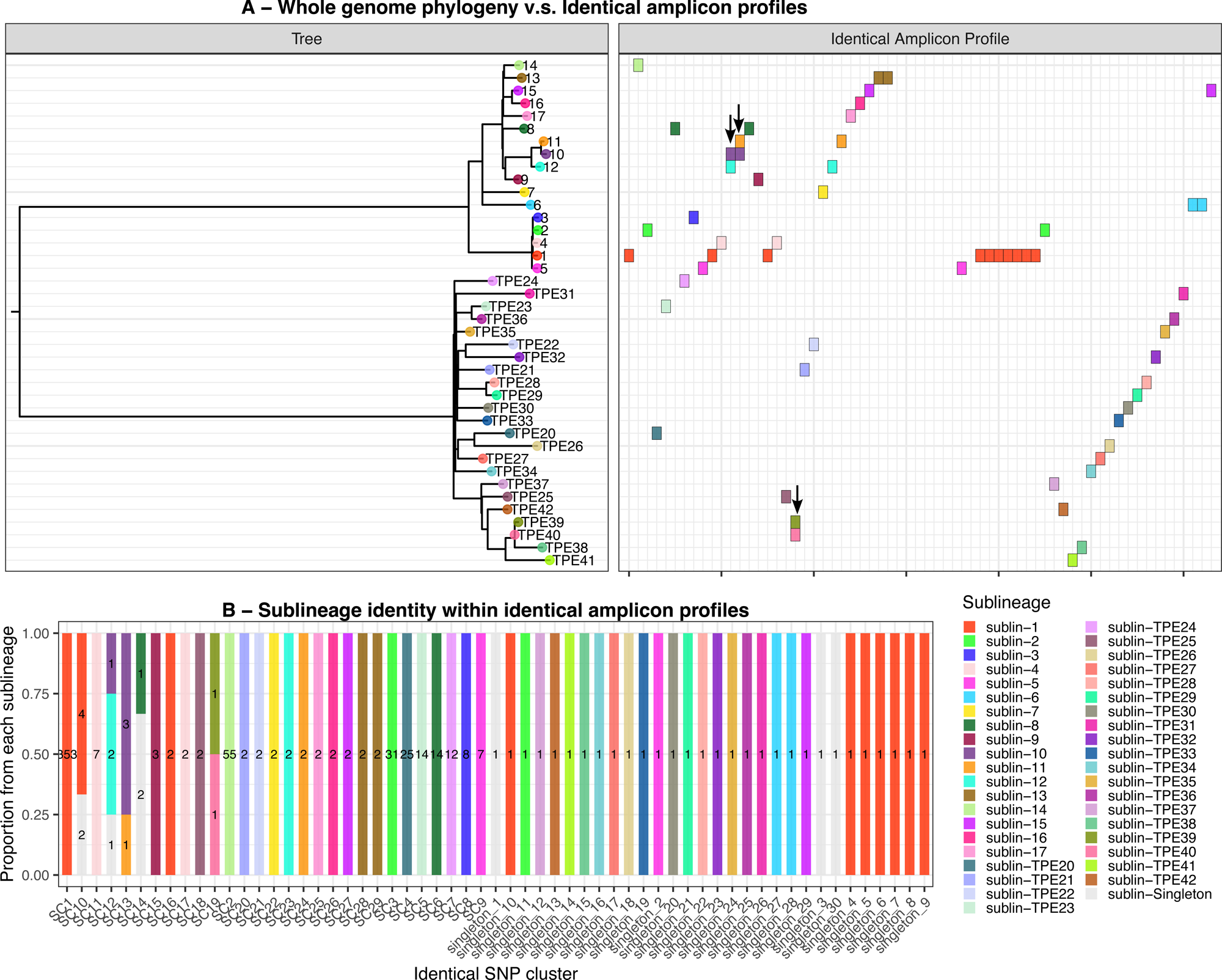
*In silico* sublineage concordance between WGS and amplicons. A – Collapsed WGS phylogeny (one tip per sublineage), against Amplicon profiles (identical sequences). Each column represents a cluster of identical amplicon profiles, coloured by the whole genome sublineage. Amplicon profiles which include multiple sublineages (indicated by arrows) are not fully resolved. Some sublineages consist of multiple amplicon profiles, reflecting expected SNP diversity in the amplicons. B – Distribution of samples within the amplicon profiles according to sublineage. Bars are coloured by sublineage proportion, and numbers indicate the genome count in the analysis.

**Supplementary Figure 12.**
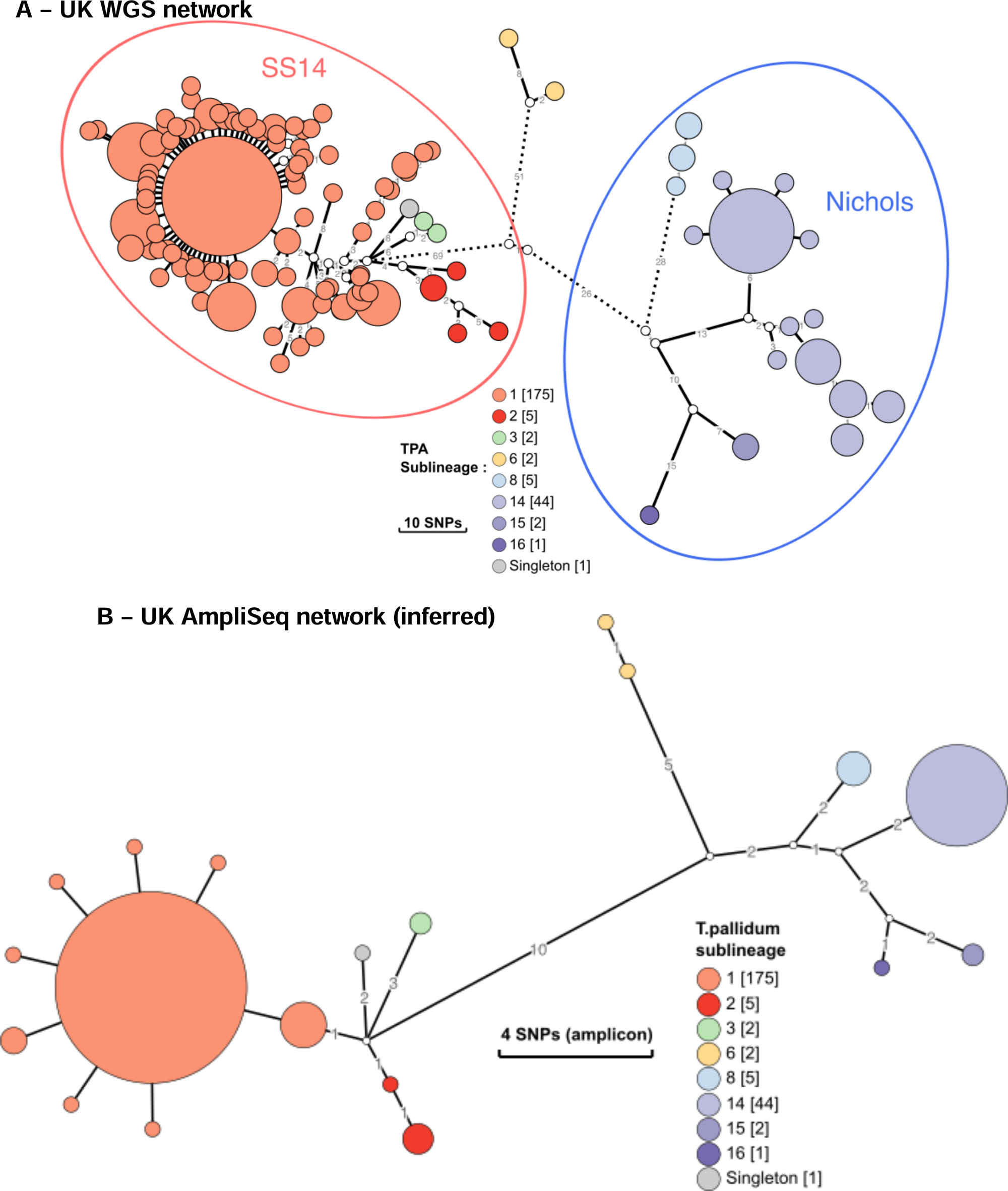
*In silico* reconstruction of UK syphilis population structure using genomes and amplicons. A – Minimum spanning tree of UK syphilis genomes coloured by sublineages (Beale 2023). Branch lengths indicate SNPs, and dotted lines indicate truncated branches ≥20 SNPs. B – Minimum spanning tree of UK syphilis dataset simulated using final 59-amplicon TP-Phylo-Plex scheme.

**Supplementary Figure 13.**
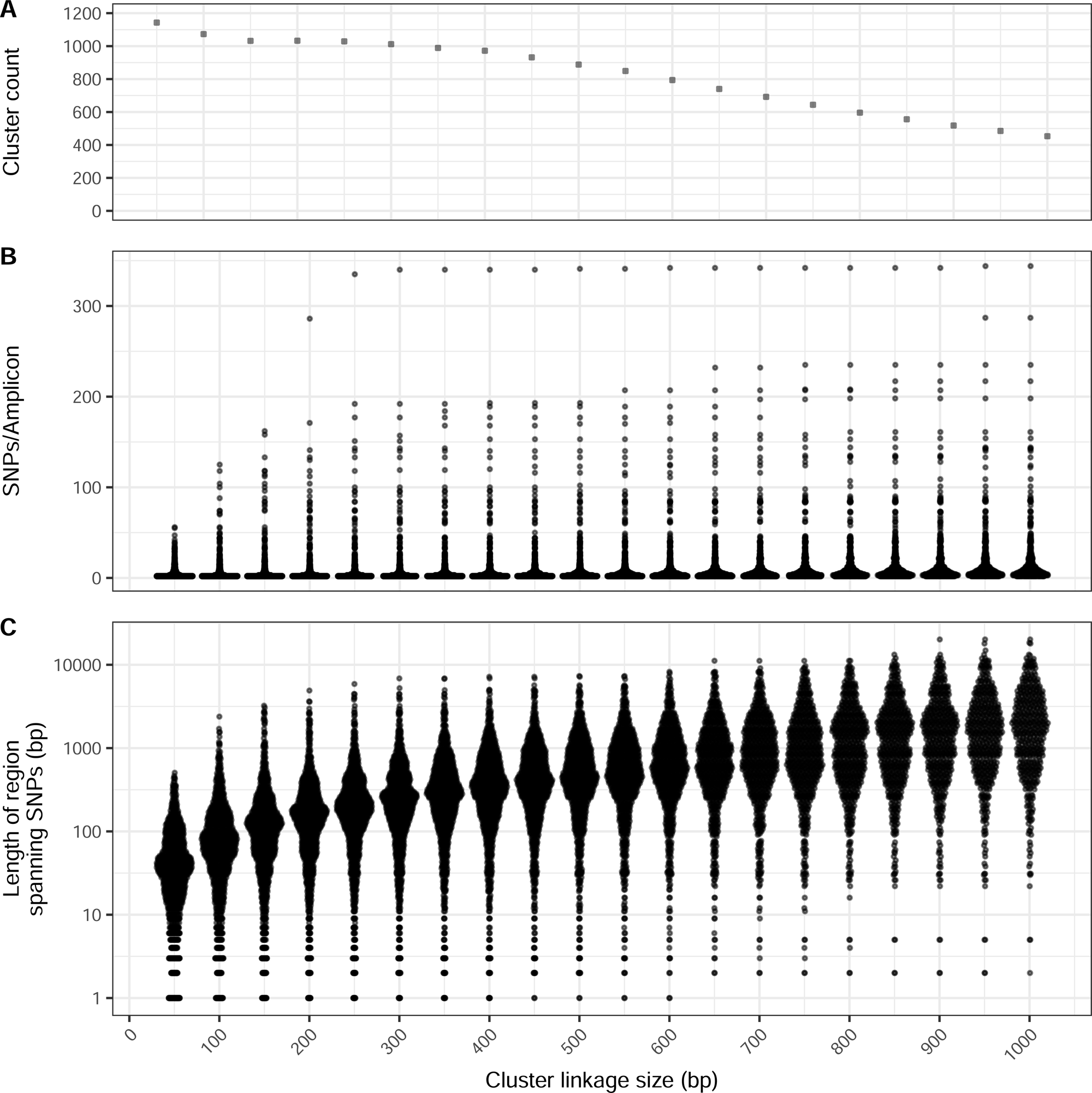
Effect of changing distance used to link SNPs for *Neisseria gonorrhoea*. Plots show cluster count, SNPs per amplicon and minimum amplicon size.

**Supplementary Figure 14.**
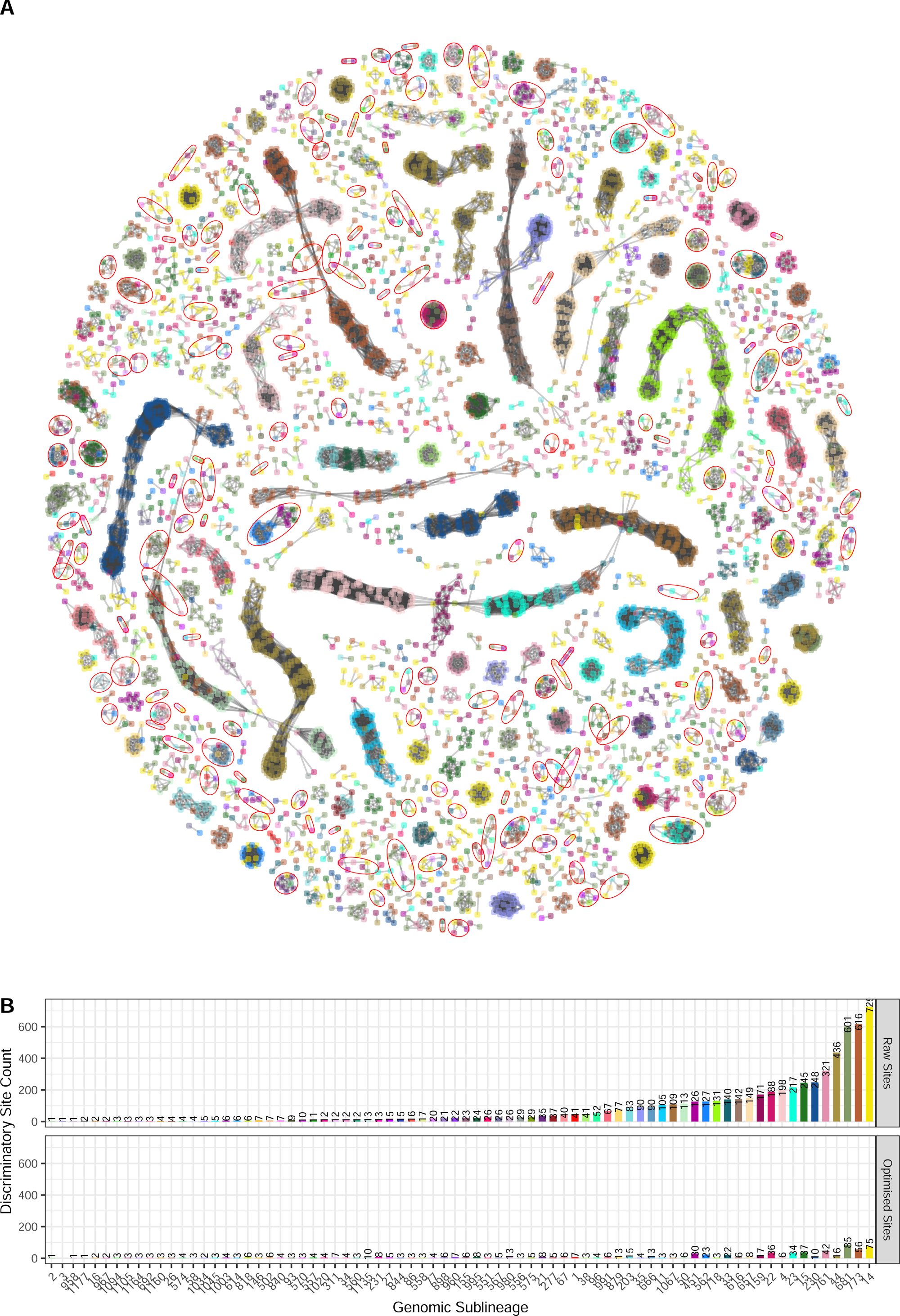
Selection of optimal amplicons (Phy-cons) enables design of an efficient scheme to recover transmission clusters in *Neisseria gonorrhoea*. A - Network showing the discriminatory SNPs coloured by the lineage they define and clustered by genome position. Nodes indicate individual SNPs and are coloured according to the sublineage supported. Edges indicate SNPs ≤ 300bp from each other, and form clusters of information-rich genomic regions. Red rings indicate clusters included in the final design. Large blocks of linked SNPs associated with the same sublineage likely indicate regions of recombination. B - Discriminatory site support for each sublineage, showing number of discriminatory sites specifically supporting each lineage and sublineage when considering (i) all discriminatory sites identified in the genome dataset, (ii) sites included in 169 regions selected by the Phylo-Plex selection algorithm.

**Supplementary Figure 15.**
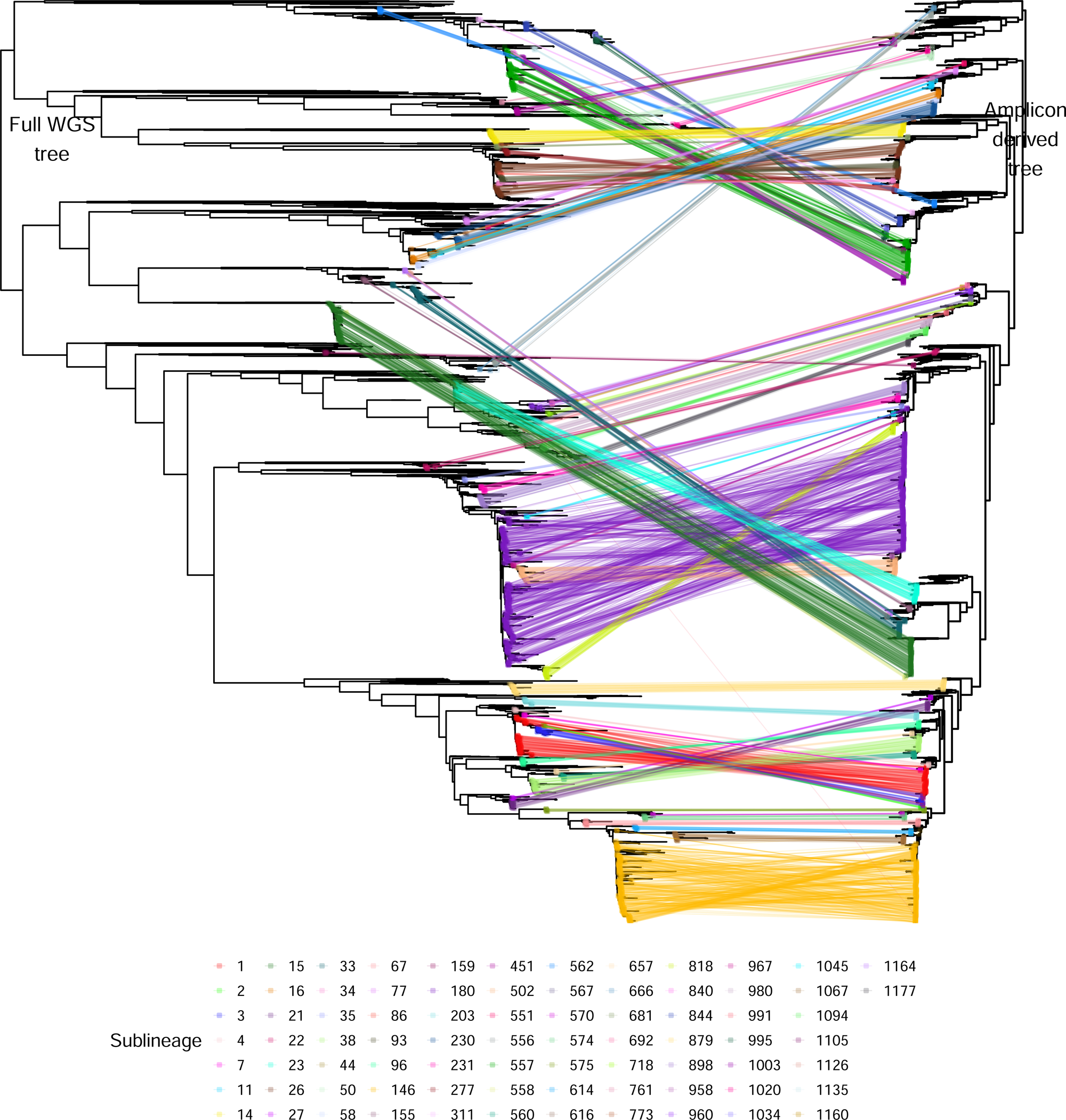
Phylogenetic recapitulation of *Neisseria gonorrhoeae* whole genome population structure and transmission clusters using Phylo-Plex. Tanglegram comparing whole genome phylogeny with phylogeny calculated from the *in silico* predicted amplicons. Broad sublineage clustering is replicated in the vast majority of cases. Note, tree scales are not identical, since the branch lengths in the amplicon-derived tree were extended to illustrate differences; the underlying topologies were not changed.

**Supplementary Figure 16.**
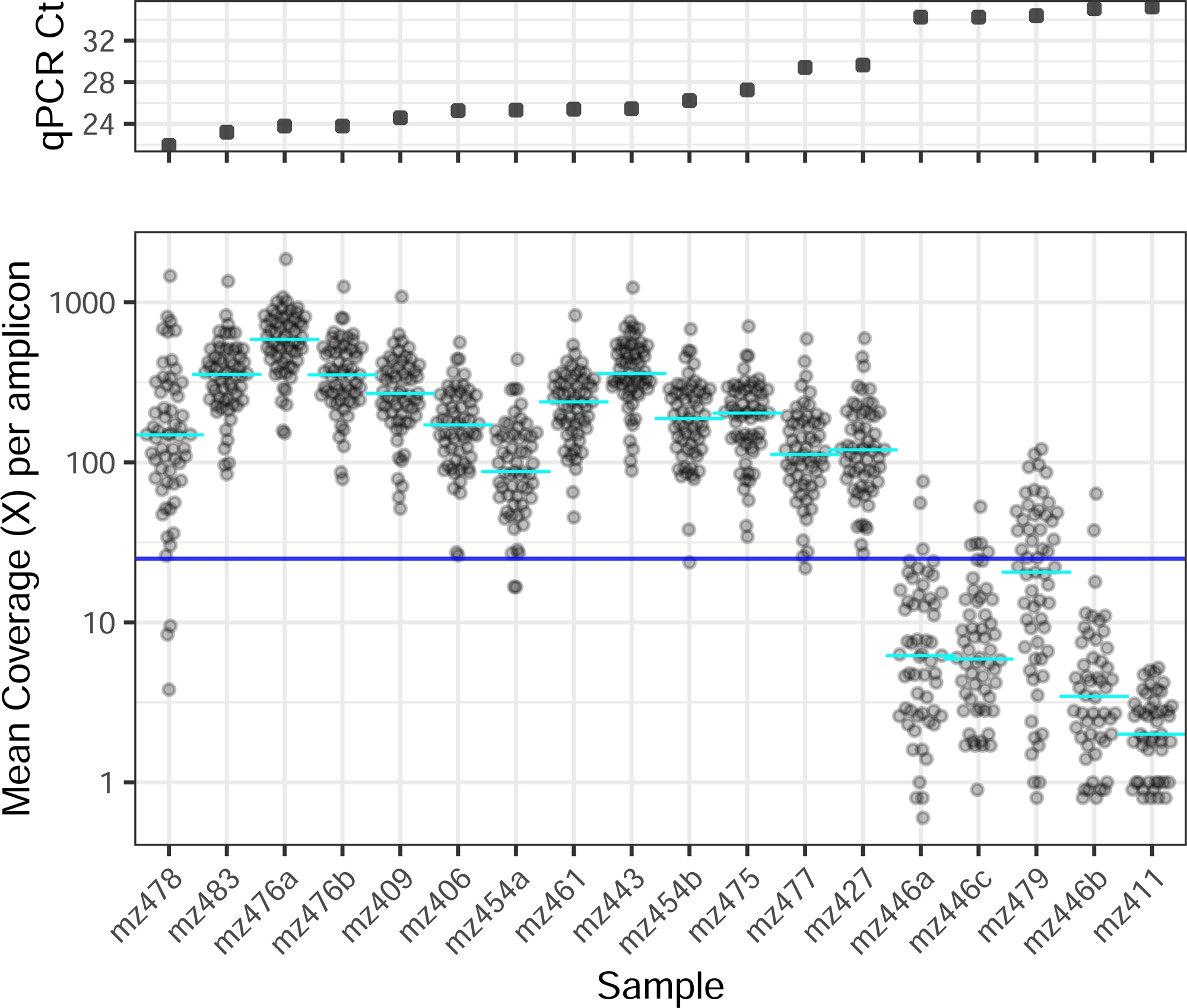
Amplicon recovery for 17 samples at Zimbabwe field site. Points show mean coverage for 59 amplicons from each of 14 samples collected, processed and sequenced in Zimbabwe, ordered by input Treponema qPCR Ct. Blue line indicates 25X coverage threshold. Includes technical replicates (mz476a/b, mz446a/b/c).

**Supplementary Figure 17.**
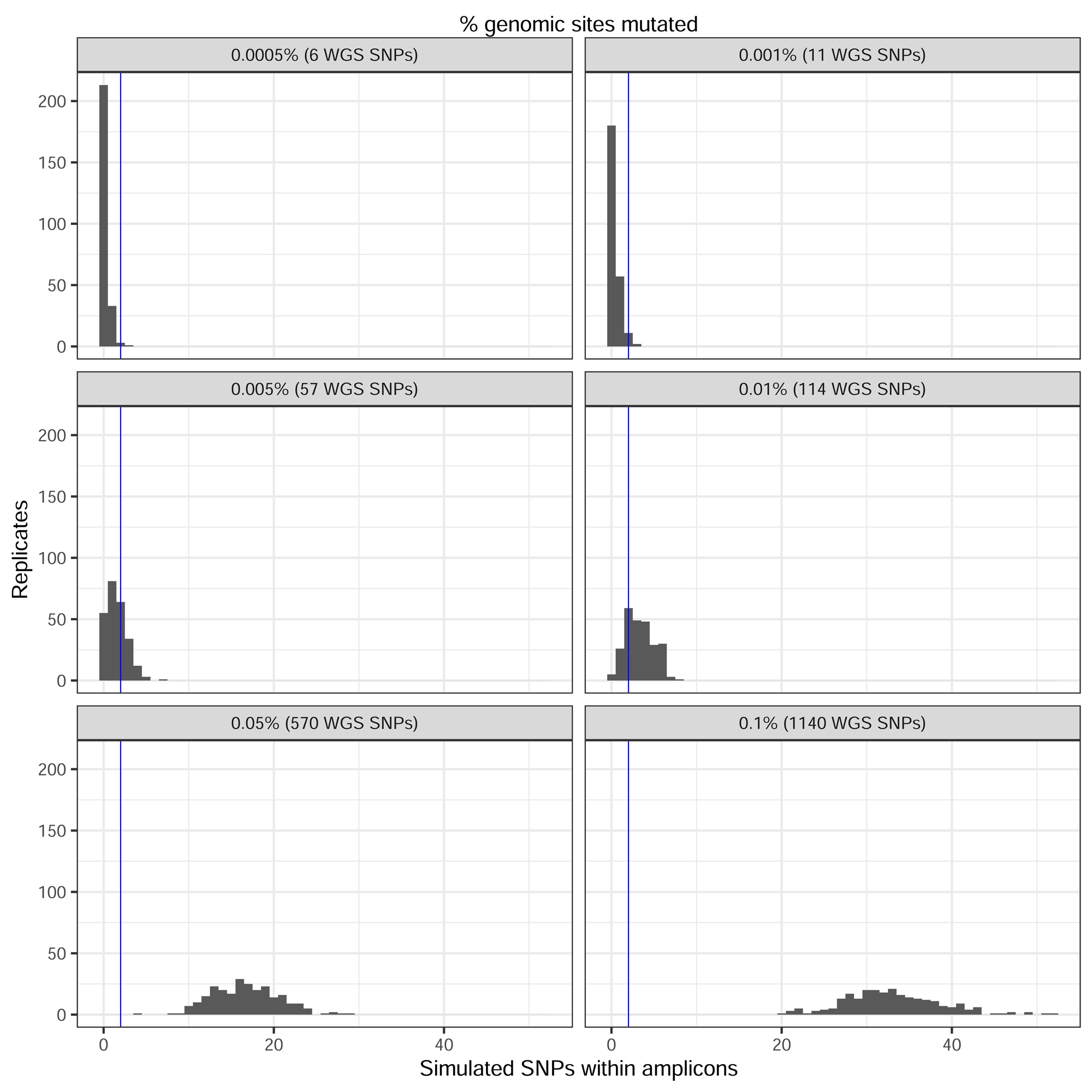
Phylo-Plex can detect *de novo* mutations and novel sublineages but is insensitive to minor changes. *In silico* mutations in whole *T. pallidum* genomes contained within TP-Phylo-Plex amplicons (250 simulations). With 0.001% genomic sites mutated (11 SNPs – similar to discriminatory level within most *T. pallidum* sub-lineages), only 5.2% of replicates had ≥2 SNPs occurring within Phy-cons. However, at 0.005% genome sites mutated (56 SNPs), this rose to 45.6% of replicates, and at 0.01% genomic sites (113 SNPs), it rose to 87.6% of replicates. Blue line indicates 2 SNPs (minimum SNPs needed to theoretically detect a sublineage as different in amplicon data).

**Supplementary Figure 18.**
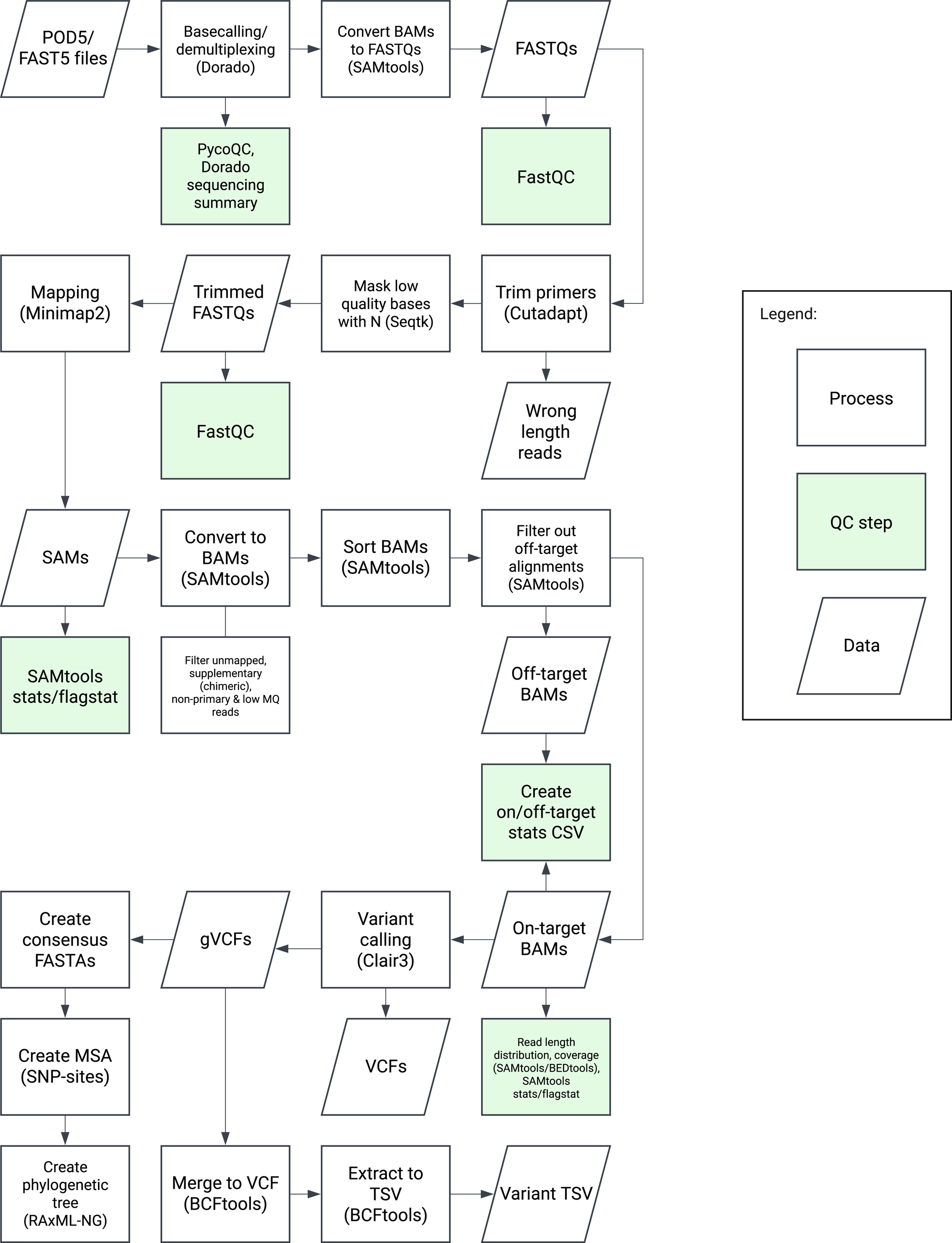
Workflow of NextFlow pipeline for automated processing of Phylo-Plex amplicon sequence data. Complete workflow for processing raw ONT sequence files (POD5/Fast5), including basecalling, quality assessment, trimming and filtering, mapping, coverage assessment, variant calling and pseudosequence generation.

**Supplementary Table 1. Metadata and list of genomes used for designing the TP-Phylo-Plex scheme.**

**Supplementary Table 2. Primers designed for TP-Phylo-Plex scheme.**

**Supplementary Table 3. Description of sequences and metadata for validation samples from South Africa.**

**Supplementary Table 4. Description of sequences and metadata for field work samples from Zimbabwe.**

**Supplementary Table 5. Cost breakdown of applying the Phylo-Plex method to MLST.**

**Supplementary Table 6. Genomic regions inferred for efficient reconstruction of *Neisseria gonorrhoeae* transmission clusters.**

