## Supplementary Figures for "Phylo-Plex: A phylogenetically informed, low-cost amplicon sequencing platform for deployable high-resolution genomic epidemiology"

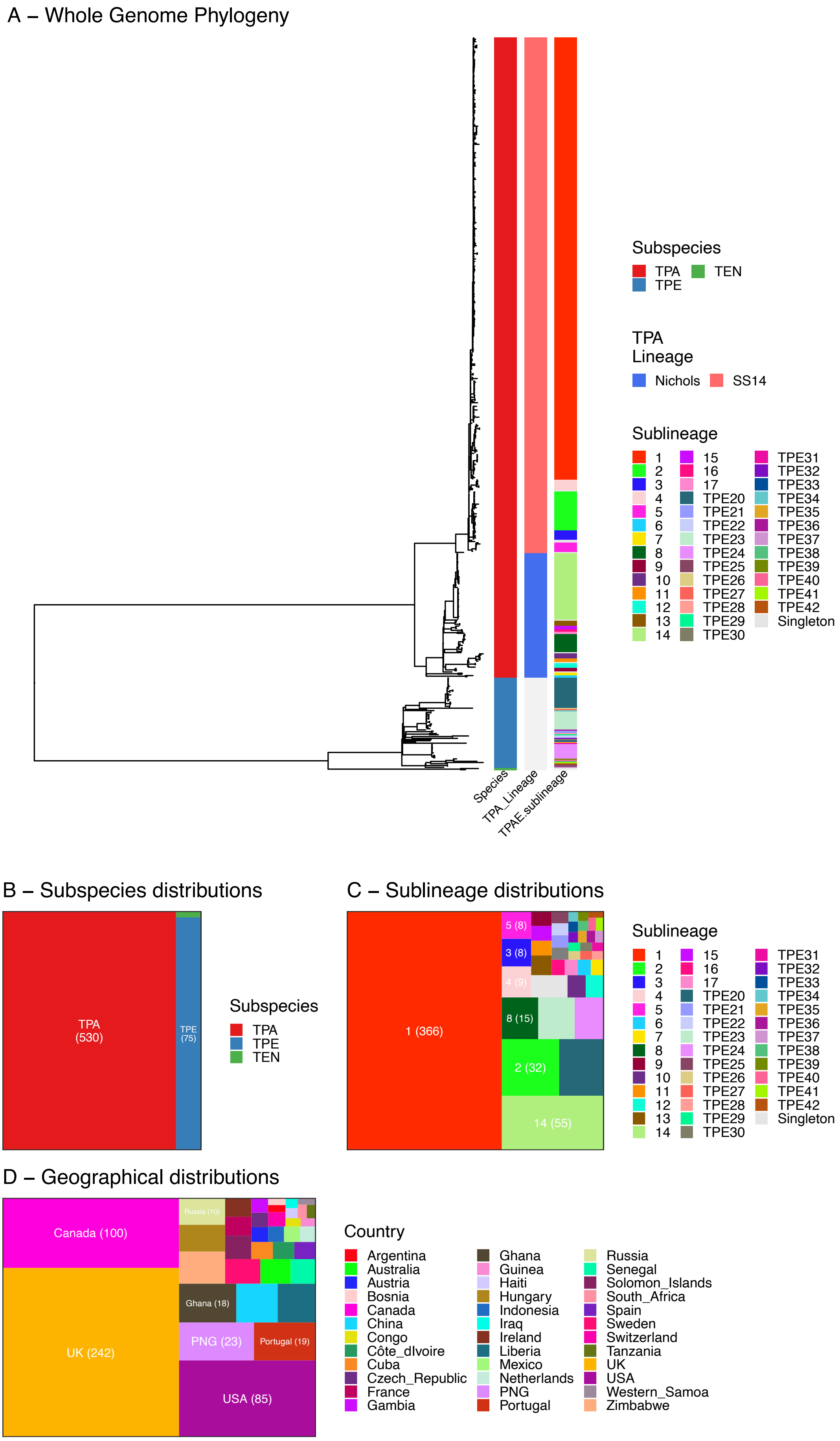
A – Whole Genome Phylogeny
Species
TPA_Lineage
TPAE sublineage
Subspecies
TPA
TEN
TPE
TPA
Lineage
Nichols
SS14
Sublineage
TPE20
TPE21
TPE22
TPE23
TPE24
TPE25
TPE26
TPE27
TPE28
TPE29
TPE30
TPE31
TPE32
TPE33
TPE34
TPE35
TPE36
TPE37
TPE38
TPE39
TPE40
TPE41
TPE42
Singleton
B – Subspecies distributions
TPA
(530)
TPE
(75)
Subspecies
TPA
TPE
TEN
C – Sublineage distributions
1 (366)
5 (8)
3 (8)
4 (9)
8 (15)
2 (32)
14 (55)
Sublineage
TPE20
TPE21
TPE22
TPE23
TPE24
TPE25
TPE26
TPE27
TPE28
TPE29
TPE30
TPE31
TPE32
TPE33
TPE34
TPE35
TPE36
TPE37
TPE38
TPE39
TPE40
TPE41
TPE42
Singleton
D – Geographical distributions
Canada (100)
Russia (10)
Ghana (18)
UK (242)
PNG (23)
Portugal (19)
USA (85)
Country
Argentina
Australia
Austria
Bosnia
Canada
China
Congo
Côte_dIvoire
Cuba
Czech_Republic
France
Gambia
Ghana
Guinea
Haiti
Hungary
Indonesia
Iraq
Ireland
Liberia
Mexico
Netherlands
PNG
Portugal
Russia
Senegal
Solomon_Islands
South_Africa
Spain
Sweden
Switzerland
Tanzania
UK
USA
Western_Samoa
Zimbabwe

**Supplementary Figure 1. Population structure and characteristics of initial *Treponema* dataset used for Phylo-Plex design.** A – Maximum Likelihood phylogeny of 607 *Treponema pallidum* genomes, with coloured tracks showing Subspecies, Lineage (within subspecies *pallidum*), and Sublineage (across all subspecies). B - Treemap plot showing distribution of genomes by *T. pallidum* subspecies: TPA – subspecies *pallidum*, TPE – subspecies *pertenue*, TEN – subspecies *endemicum*. C - Treemap plot showing distribution of genomes by *T. pallidum* subspecies (clustered as samples sharing a common ancestral node ≤10 SNPs. D - Treemap plot showing distribution of genomes by country.

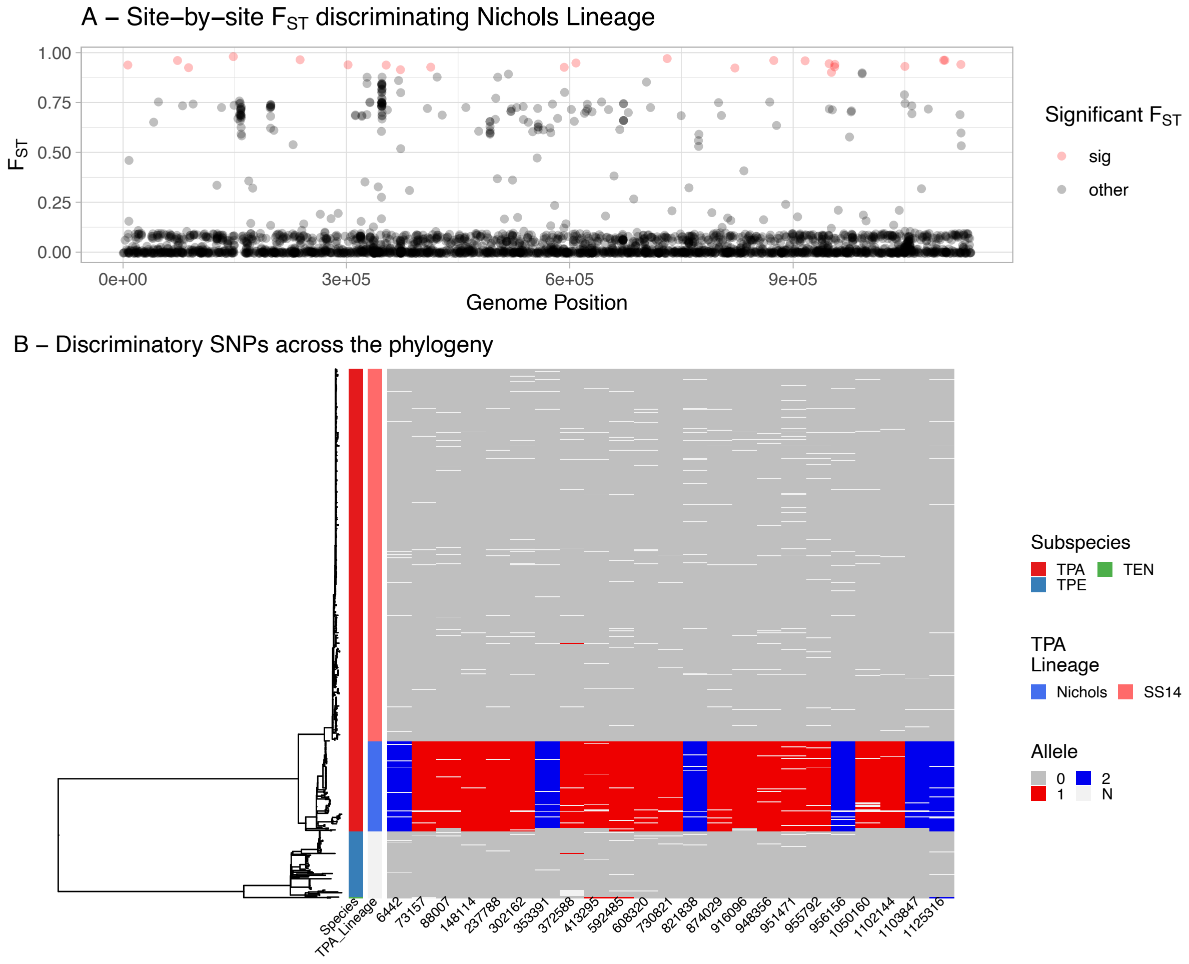

**Supplementary Figure 2. Identifying discriminatory SNPs for a single lineage.**
A – Population fixation analysis ($F_{ST}$) of variable sites. Sites in red discriminate *T. pallidum* Nichols Lineage from all other lineages at $F_{ST} \geq 0.9$. B – Maximum likelihood whole genome phylogeny showing allelic identity at each discriminatory site identified in A. Colours indicate allele detected (pale grey indicates 'N', where data was missing – common in metagenomic data).

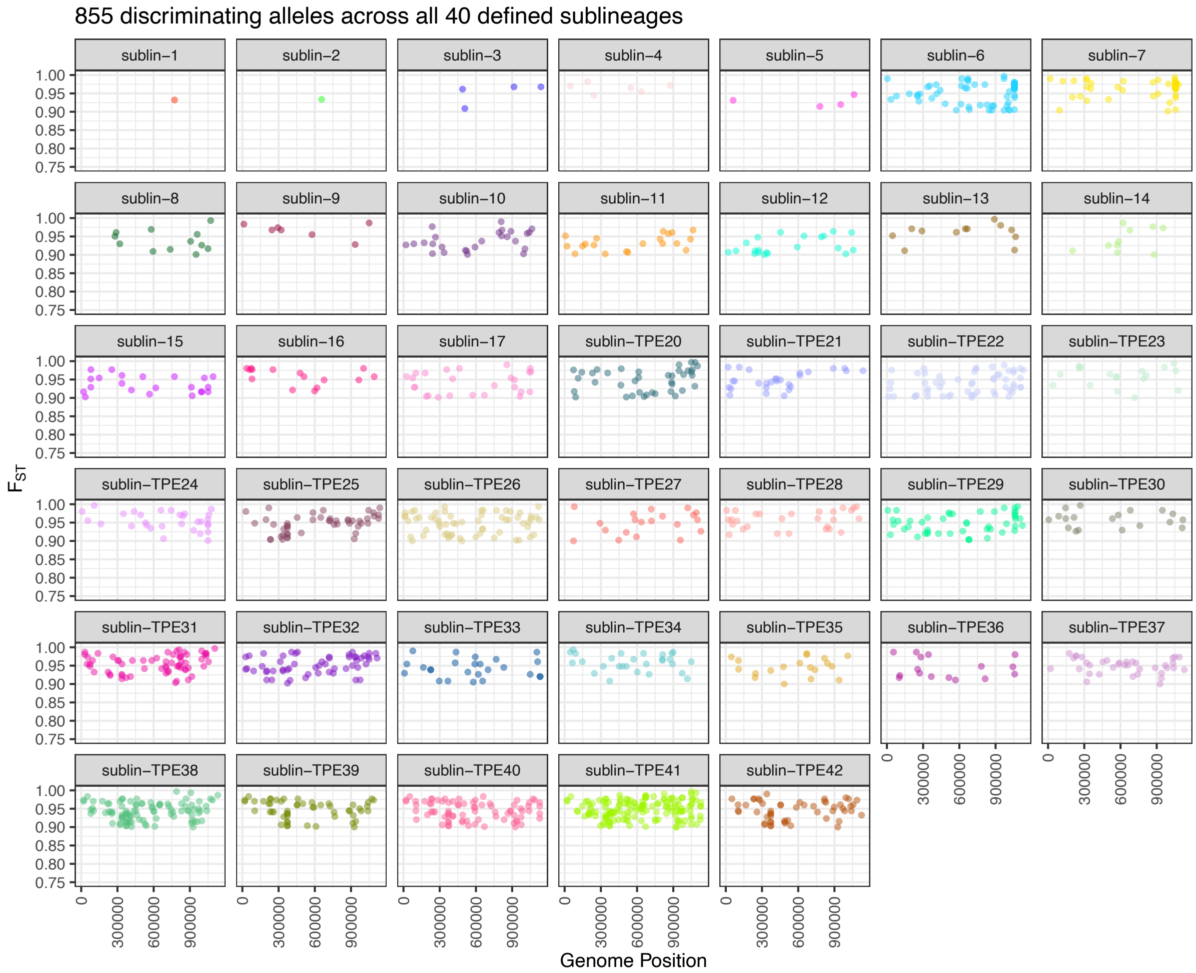

**Supplementary Figure 3. Identifying discriminatory SNPs for 40 sublineages.**
Population fixation analysis ($F_{ST}$) of variable sites for each sublineage - plots show only sites with $F_{ST}$ ≥0.9.

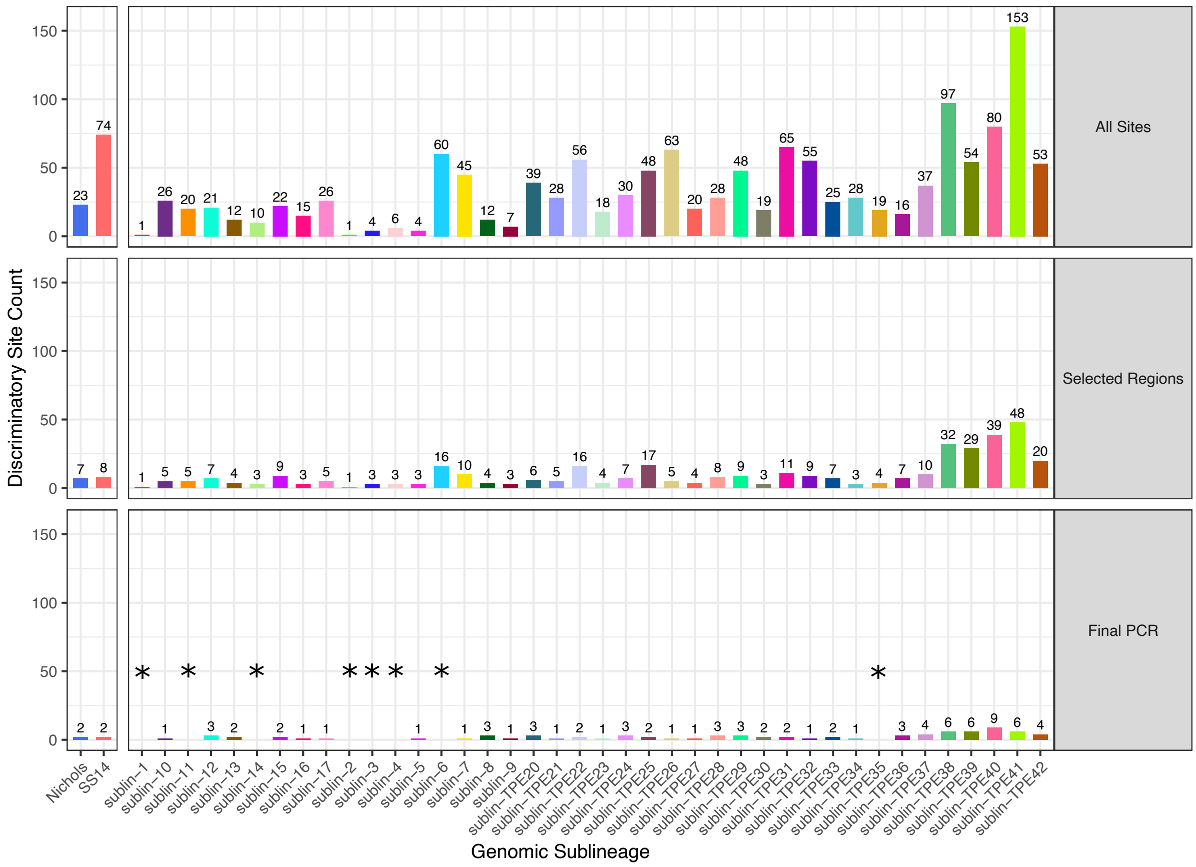

**Supplementary Figure 4. Discriminatory site support for each sublineage.**
Plot shows number of discriminatory sites specifically supporting each lineage and sublineage when considering (i) all discriminatory sites identified in the genome dataset, (ii) sites included in 74 regions selected by the Phylo-Plex selection algorithm, (iii) sites remaining after full optimization of the 59-amplicon multiplex PCR. The final PCR used for evaluation removed 15 amplicons and therefore removed direct support for 8 sublineages ('*'), but future iterations would modify the primer designs and balance to reinclude these.

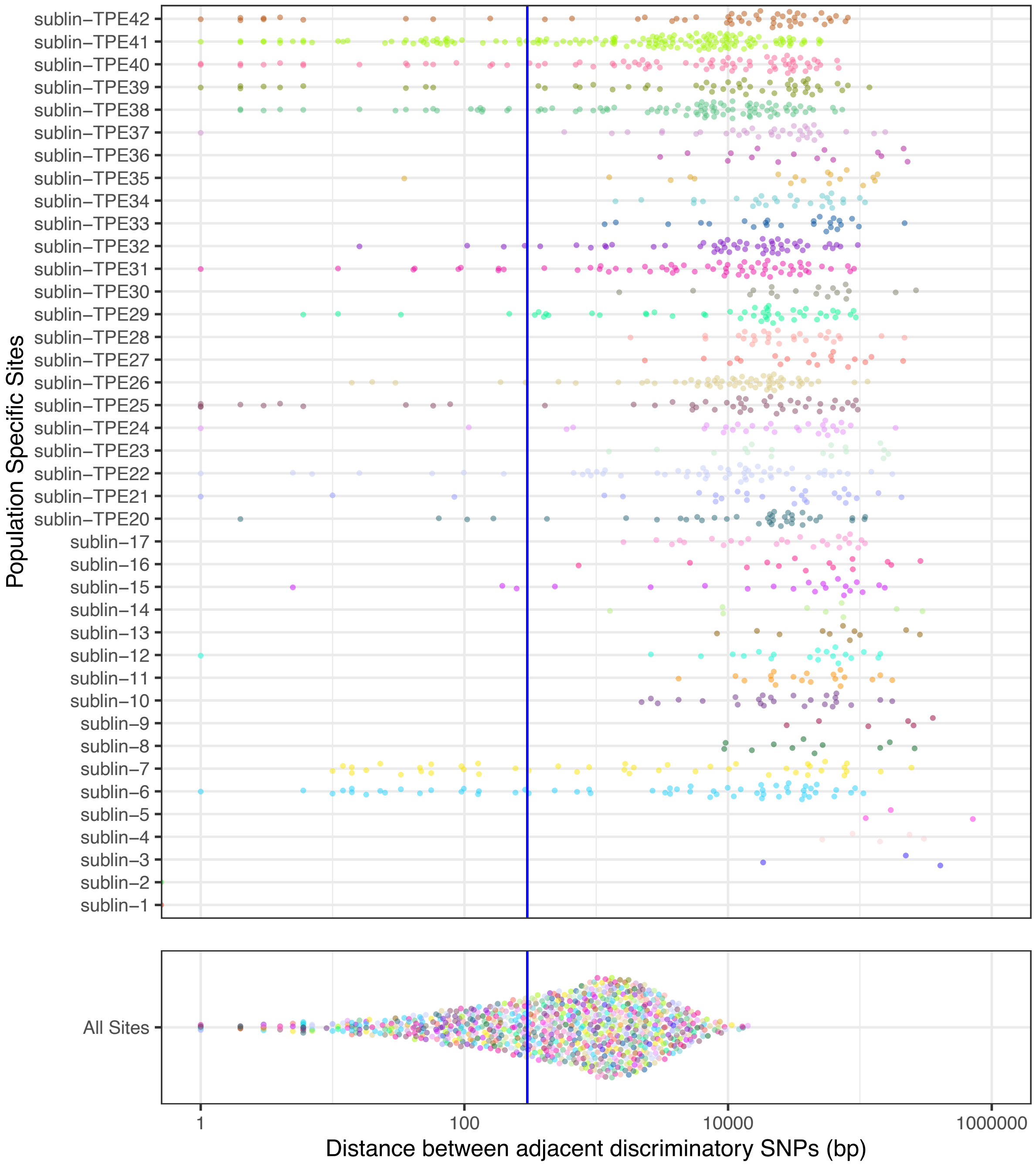

**Supplementary Figure 5. Genomic distance between individual sites is reduced when considered as a total population.**

Genetic distance between adjacent discriminatory SNPs according to sublineage and as a total population. Blue line highlights 300 bp.

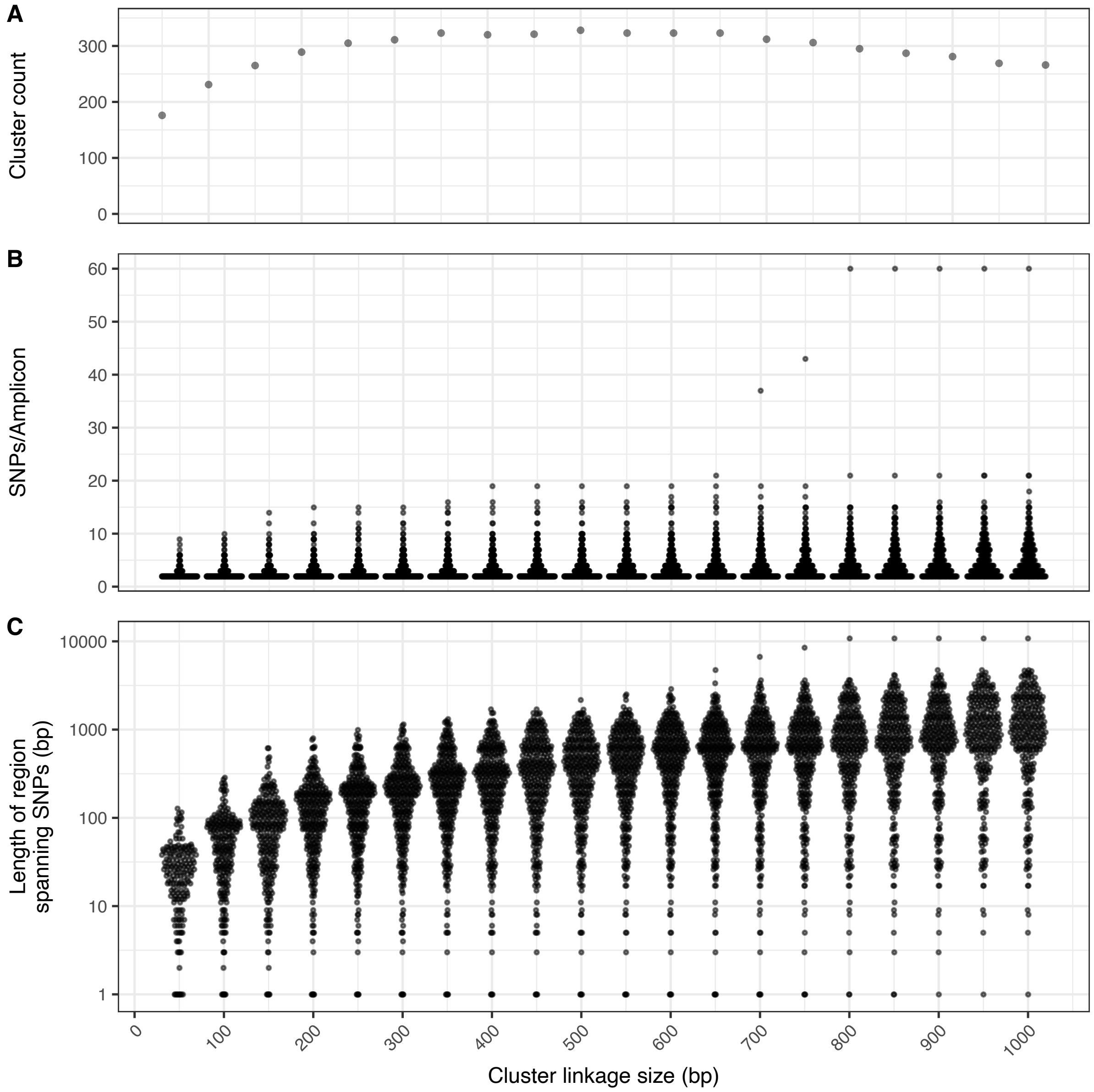

**Supplementary Figure 6. Changing distance used to link SNPs as clusters impacts cluster count, SNPs per amplicon and minimum amplicon size.**

The selection of an appropriate genomic distance for positional clustering can be tuned and impacts the number of SNPs that merge into clusters. However, increasing cluster distance between individual SNPs can also substantially impact the total length of SNP networks, resulting in candidate amplicons too long to reasonably amplify in multiplex PCR. A – Number of positional clusters produced with different genomic distance between sites. B – Number of SNPs present in each cluster (candidate amplicon) produced with different genomic distance between sites. C – Genomic distance between furthest SNPs in cluster (i.e. minimum length of candidate amplicon) produced with different genomic distance between sites.

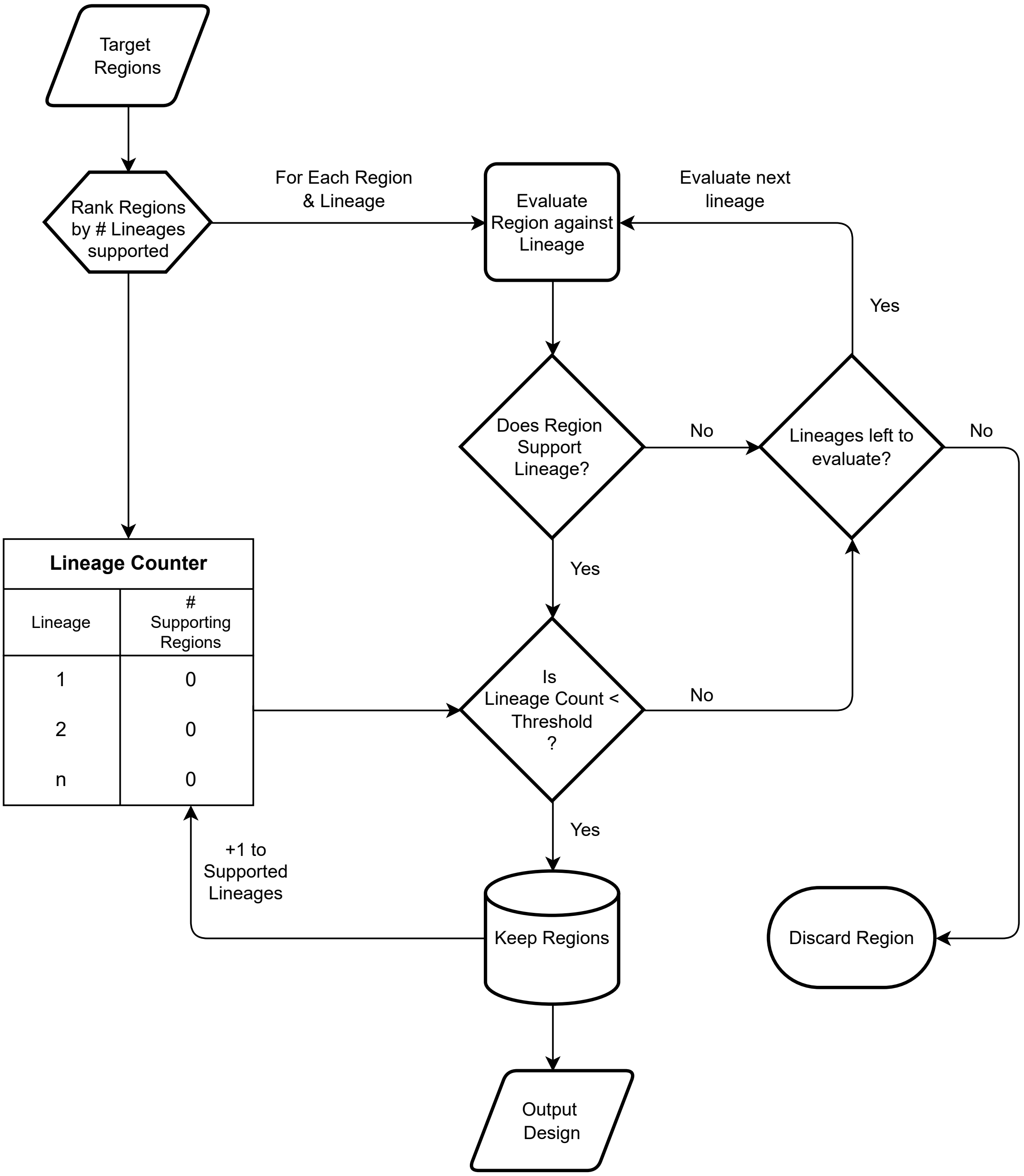

**Supplementary Figure 7. Hierarchical selection algorithm for maximising discriminatory power whilst minimising total number of amplicons.**

Each candidate region is evaluated based on the sublineages the SNPs within it support, and regions are added until support for a sublineage meets a minimum threshold (3 SNPs), after which, support for that sublineage is no longer considered.

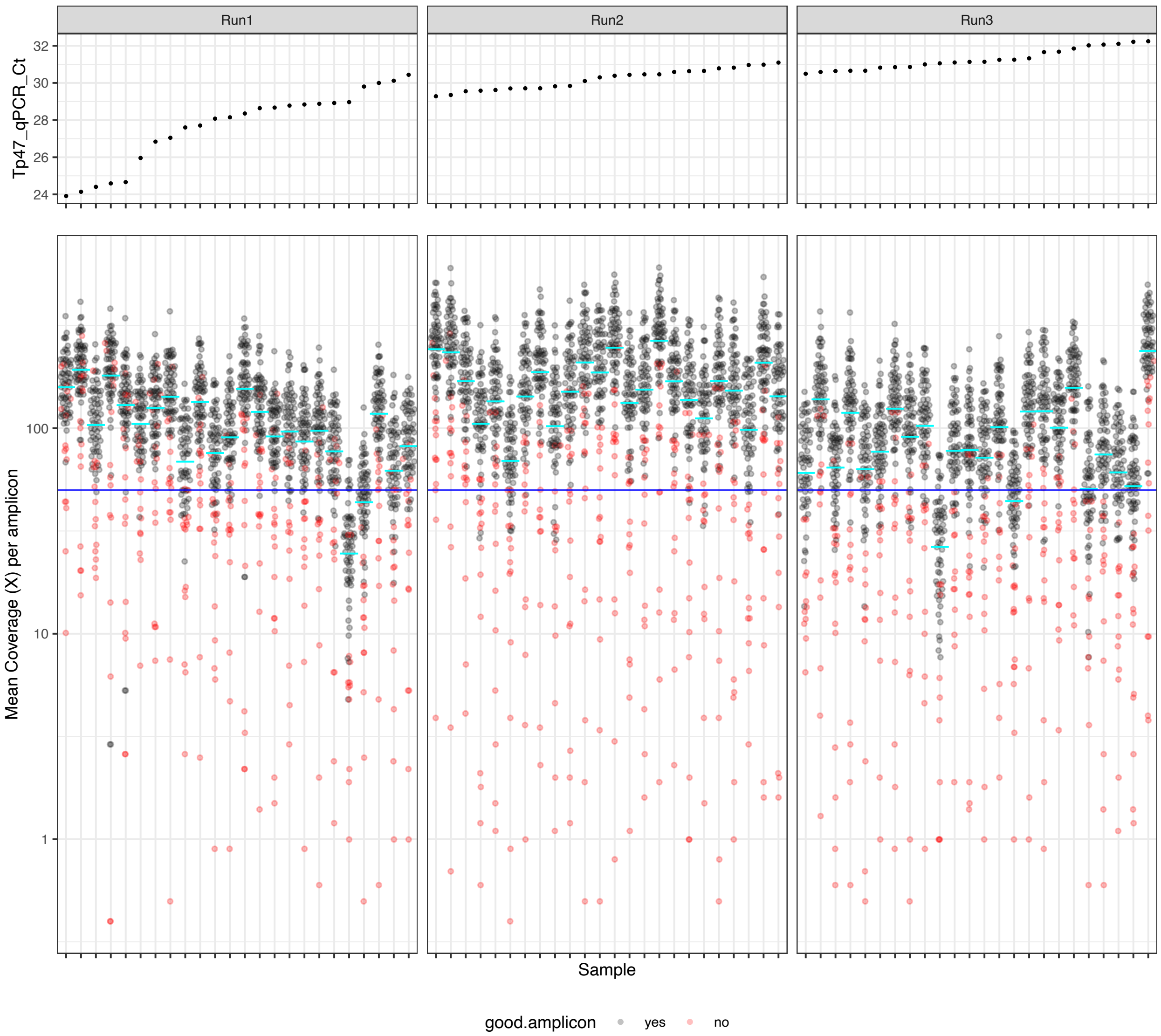

**Supplementary Figure 8. Sequencing performance of 72 South African clinical syphilis samples.** Points show mean coverage for 74 amplicons from each of 72 samples, ordered by input Treponema qPCR Ct. Samples were sequenced on three separate runs of MinION Flongle cells. Blue line indicates 50X coverage. Red points indicate 15 amplicons which consistently performed poorly (see Supplementary Figure 9). Cyan lines indicate mean coverage of all amplicons in that sample.

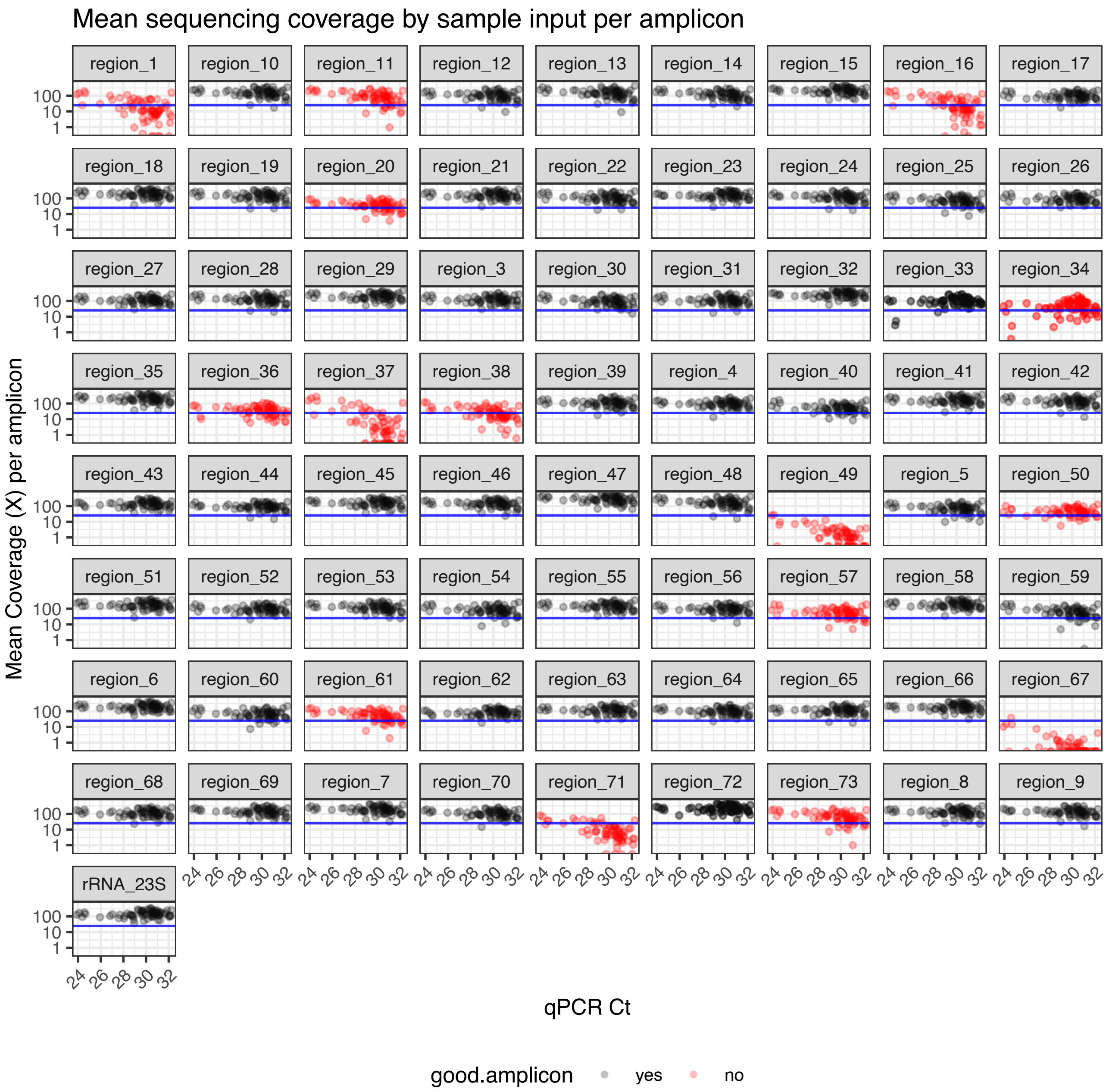

**Supplementary Figure 9. Sequencing performance of 74 multiplex amplicons in clinical syphilis samples.**

Points show mean coverage for 74 amplicons from each of 72 South African clinical syphilis samples compared to input Treponema qPCR Ct. Blue line indicates 50X coverage. Red points indicate 15 amplicons where ≥10% of samples had <25X coverage (after excluding samples which performed poorly overall).

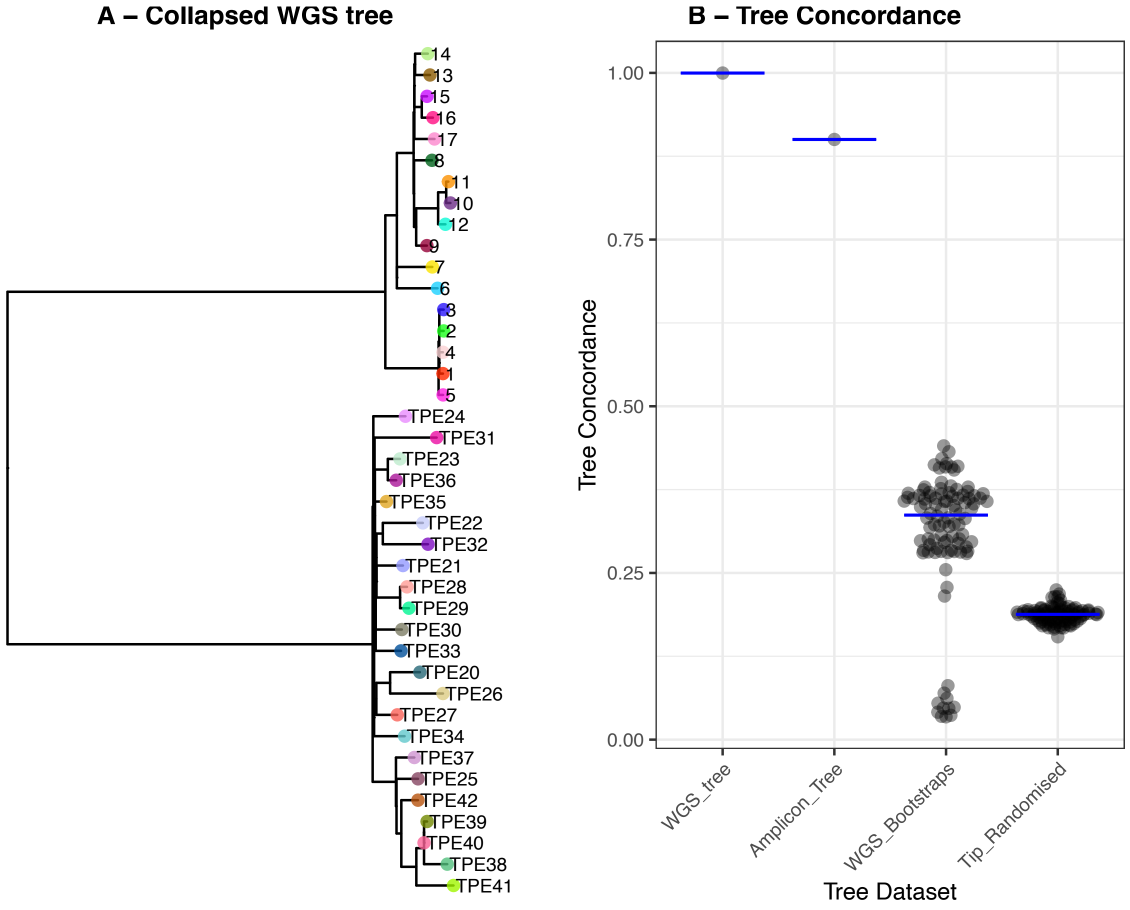

**Supplementary Figure 10. Quantitative comparison between WGS derived phylogeny and Amplicon derived phylogeny demonstrates high concordance.**

A – Collapsed WGS phylogeny (one tip per sublineage), B – Tree concordance between collapsed WGS tree and (i) full WGS tree, (ii) Amplicon derived tree (simulated), (iii) 100 bootstraps derived from full WGS tree, (iv) 100 tip-randomised full WGS trees.

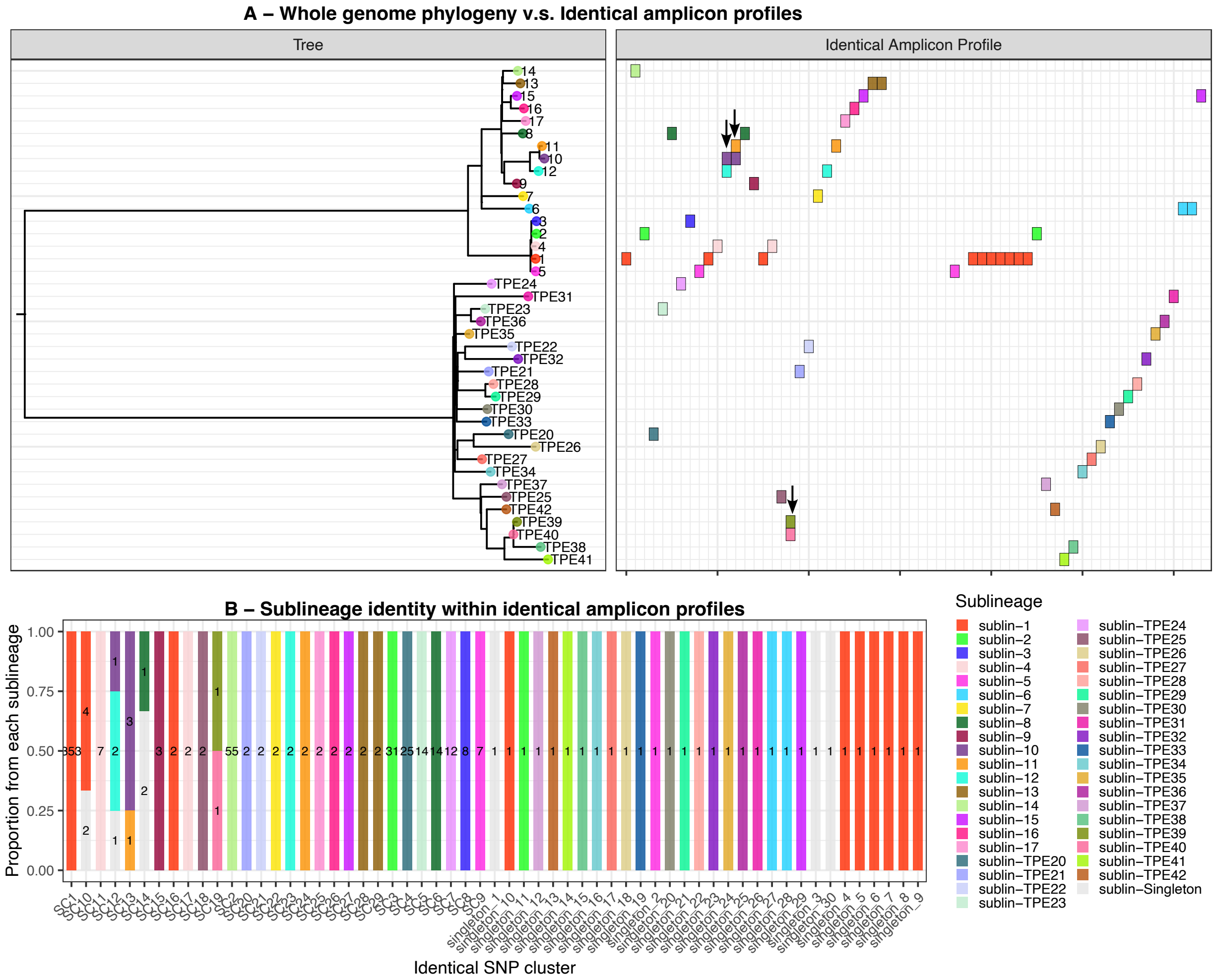

**Supplementary Figure 11.** ***In silico*** **sublineage concordance between WGS and amplicons.**

A – Collapsed WGS phylogeny (one tip per sublineage), against Amplicon profiles (identical sequences). Each column represents a cluster of identical amplicon profiles, coloured by the whole genome sublineage. Amplicon profiles which include multiple sublineages (indicated by arrows) are not fully resolved. Some sublineages consist of multiple amplicon profiles, reflecting expected SNP diversity in the amplicons. B – Distribution of samples within the amplicon profiles according to sublineage. Bars are coloured by sublineage proportion, and numbers indicate the genome count in the analysis.

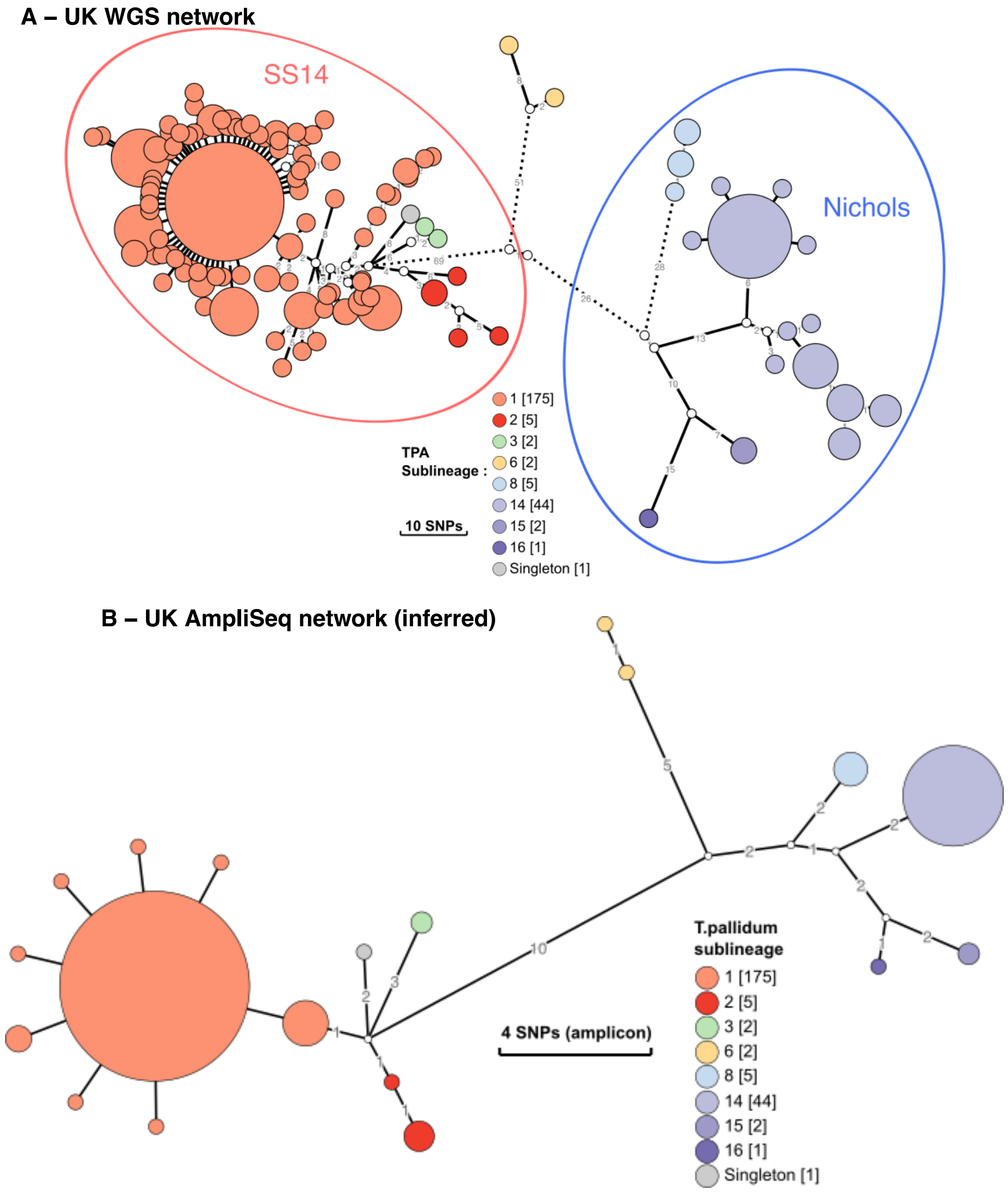

**Supplementary Figure 12. *In silico* reconstruction of UK syphilis population structure using genomes and amplicons.**
A – Minimum spanning tree of UK syphilis genomes coloured by sublineages (Beale 2023). Branch lengths indicate SNPs, and dotted lines indicate truncated branches ≥20 SNPs. B – Minimum spanning tree of UK syphilis dataset simulated using final 59-amplicon TP-Phylo-Plex scheme.

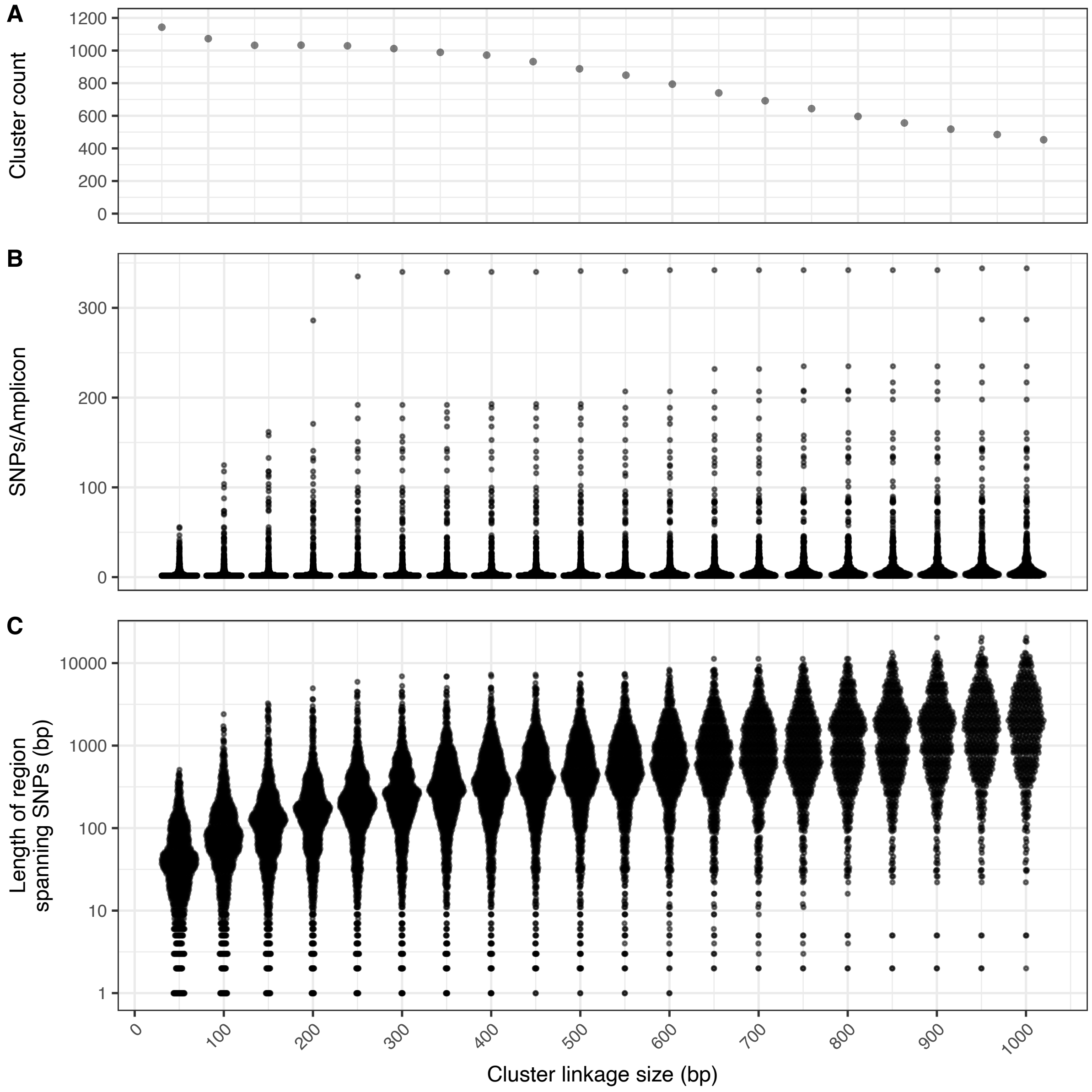

**Supplementary Figure 13. Effect of changing distance used to link SNPs for *Neisseria gonorrhoeae*.** Plots show cluster count, SNPs per amplicon and minimum amplicon size.

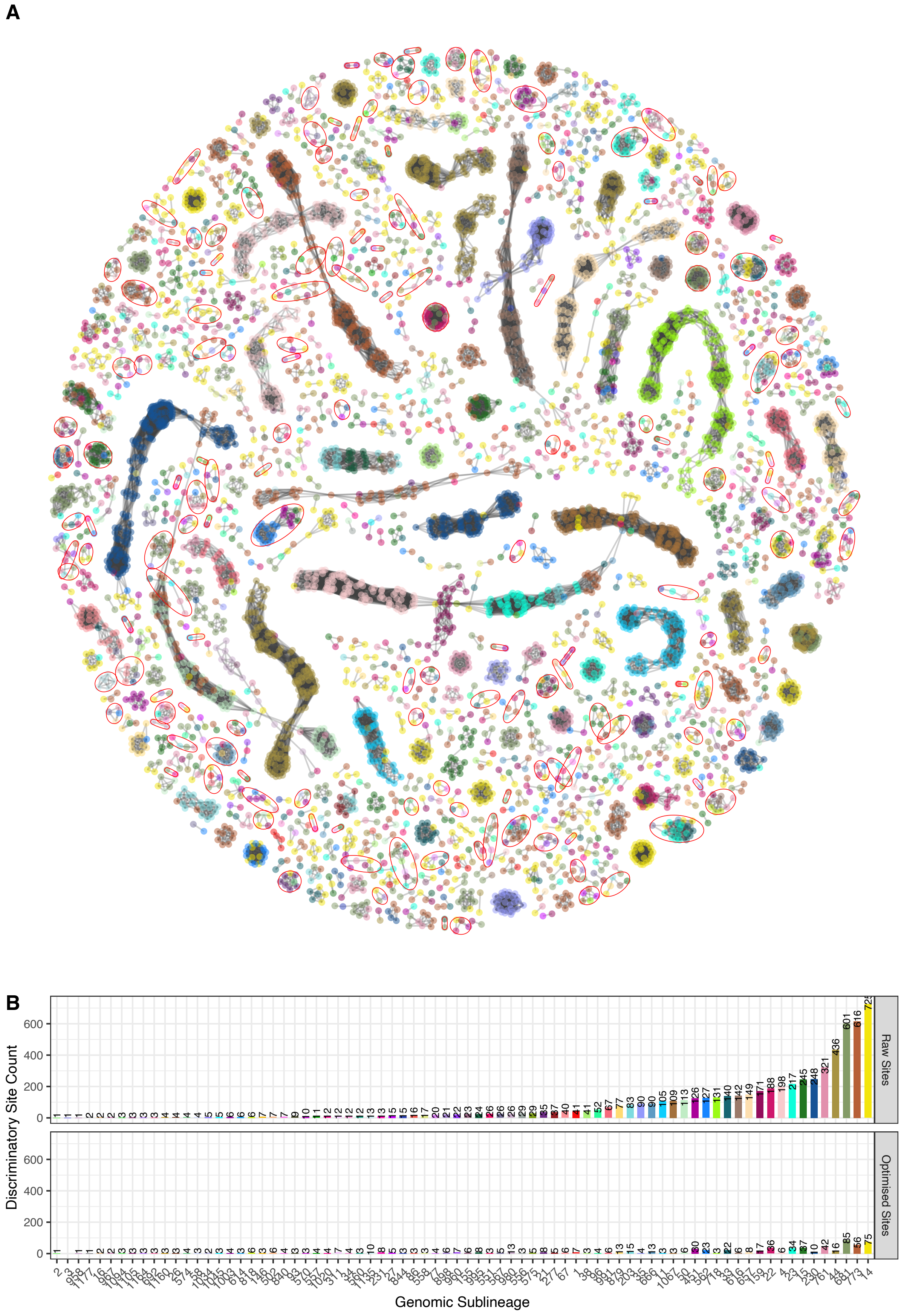
A
B
Discriminatory Site Count
Raw Sites
Optimised Sites
0
200
400
600
Genomic Sublineage

**Supplementary Figure 14. Selection of optimal amplicons (Phy-cons) enables design of an efficient scheme to recover transmission clusters in *Neisseria gonorrhoeae*.** A - Network showing the discriminatory SNPs coloured by the lineage they define and clustered by genome position. Nodes indicate individual SNPs and are coloured according to the sublineage supported. Edges indicate SNPs ≤ 300bp from each other, and form clusters of information-rich genomic regions. Red rings indicate clusters included in the final design. Large blocks of linked SNPs associated with the same sublineage likely indicate regions of recombination. B - Discriminatory site support for each sublineage, showing number of discriminatory sites specifically supporting each lineage and sublineage when considering (i) all discriminatory sites identified in the genome dataset, (ii) sites included in 169 regions selected by the Phylo-Plex selection algorithm.

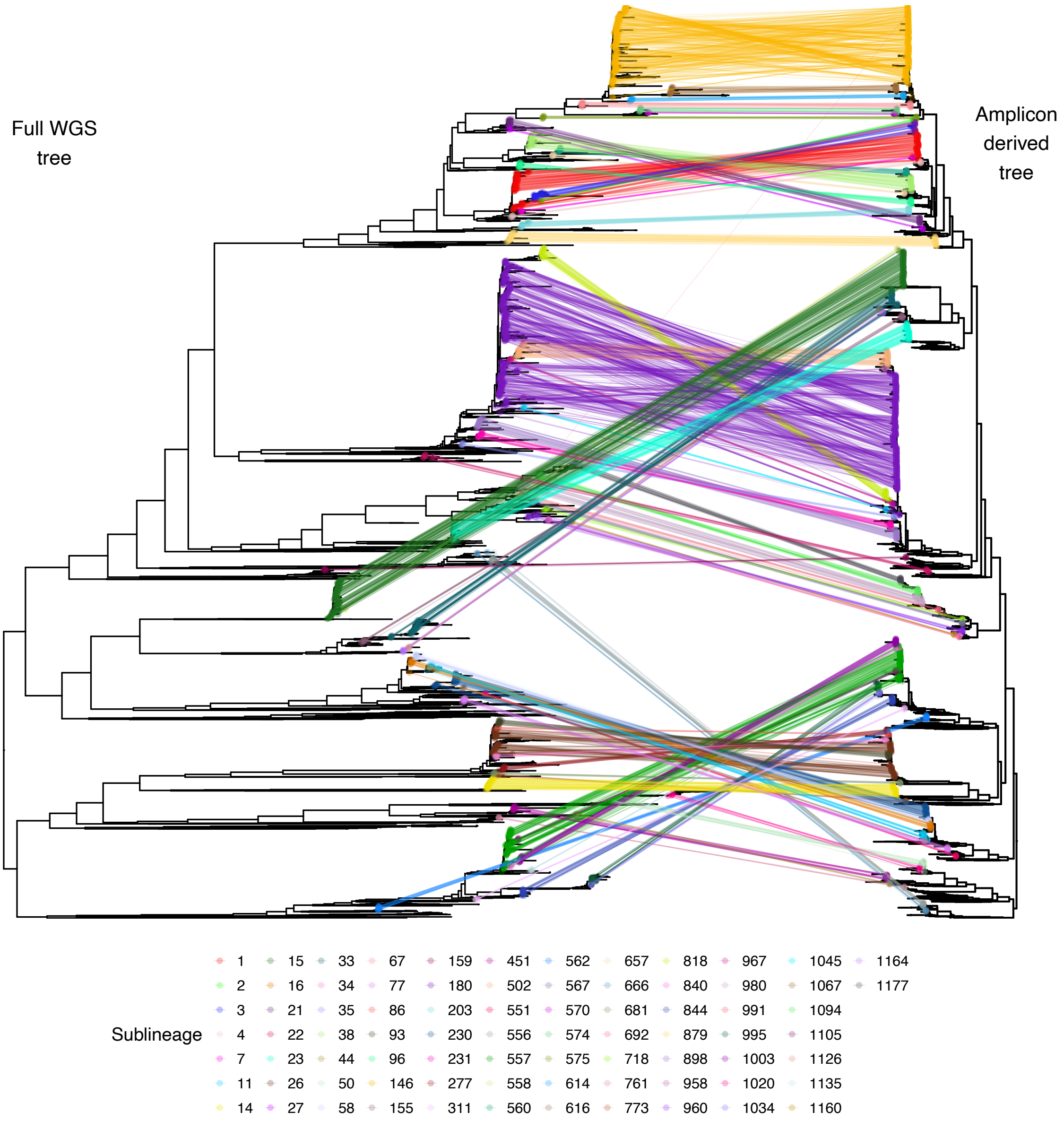

**Supplementary Figure 15. Phylogenetic recapitulation of *Neisseria gonorrhoeae* whole genome population structure and transmission clusters using Phylo-Plex.** Tanglegram comparing whole genome phylogeny with phylogeny calculated from the *in silico* predicted amplicons. Broad sublineage clustering is replicated in the vast majority of cases. Note, tree scales are not identical, since the branch lengths in the amplicon-derived tree were extended to illustrate differences; the underlying topologies were not changed.

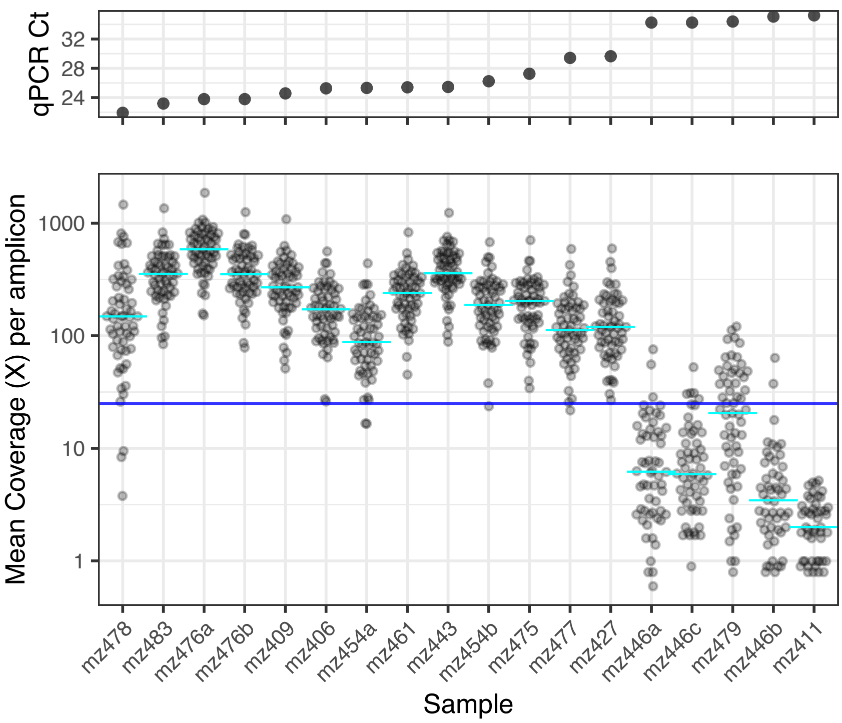

**Supplementary Figure 16. Amplicon recovery for 17 samples at Zimbabwe field site.**
Points show mean coverage for 59 amplicons from each of 14 samples collected, processed and sequenced in Zimbabwe, ordered by input Treponema qPCR Ct. Blue line indicates 25X coverage threshold. Cyan lines indicate median coverage for each sample. Includes technical replicates (mz476a/b, mz446a/c) and biological replicates (mz446a/b). Technical replicates mz476a/b had identical SNP profiles.

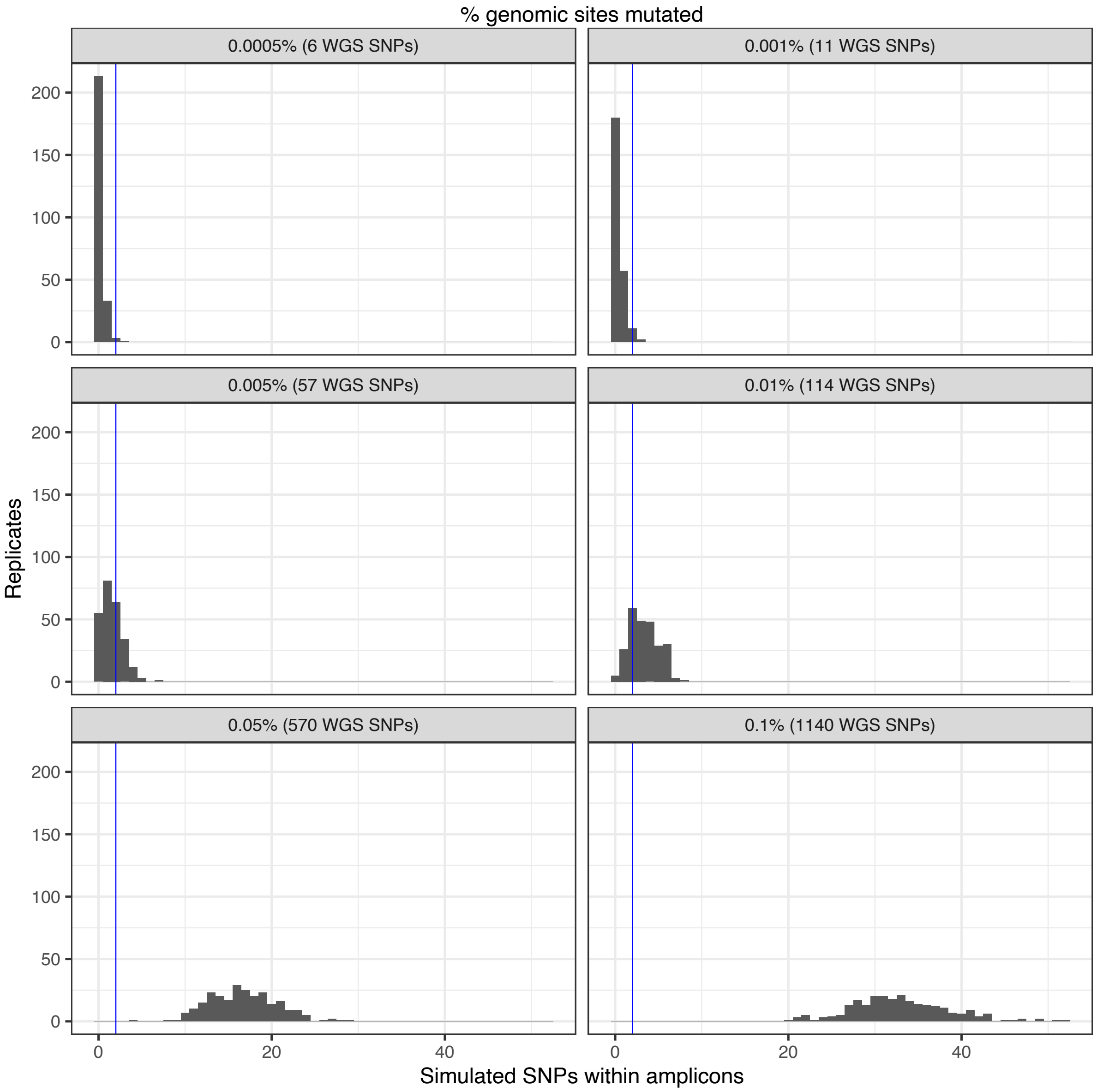

**Supplementary Figure 17. Phylo-Plex can detect *de novo* mutations and novel sublineages but is insensitive to minor changes.**

*In silico* mutations in whole *T. pallidum* genomes contained within TP-Phylo-Plex amplicons (250 simulations). With 0.001% genomic sites mutated (11 SNPs – similar to discriminatory level within most *T. pallidum* sub-lineages), only 5.2% of replicates had ≥2 SNPs occurring within Phy-cons. However, at 0.005% genome sites mutated (56 SNPs), this rose to 45.6% of replicates, and at 0.01% genomic sites (113 SNPs), it rose to 87.6% of replicates. Blue line indicates 2 SNPs (minimum SNPs needed to theoretically detect a sublineage as different in amplicon data).

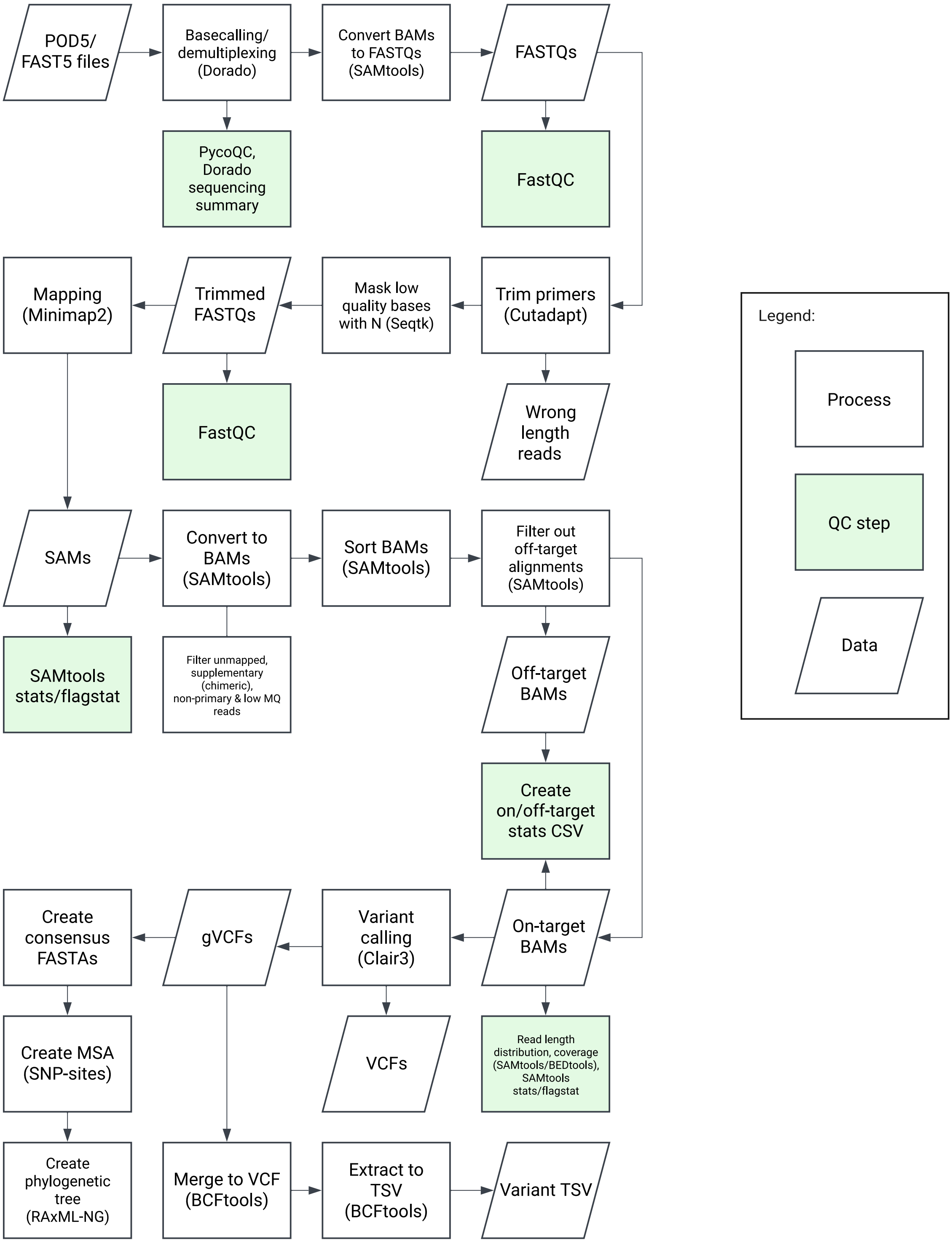

**Supplementary Figure 18. Workflow of NextFlow pipeline for automated processing of Phylo-Plex amplicon sequence data.**

Complete workflow for processing raw ONT sequence files (POD5/Fast5), including basecalling, quality assessment, trimming and filtering, mapping, coverage assessment, variant calling and pseudosequence generation.

**Supplementary Table 1. Metadata and list of genomes used for designing the TP-Phylo-Plex scheme.**

**Supplementary Table 2. Primers designed for TP-Phylo-Plex scheme.**

**Supplementary Table 3. Description of sequences and metadata for validation samples from South Africa.**

**Supplementary Table 4. Description of sequences and metadata for field work samples from Zimbabwe.**

**Supplementary Table 5. Cost breakdown of applying the Phylo-Plex method to MLST.**

**Supplementary Table 6. Genomic regions inferred for efficient reconstruction of *Neisseria gonorrhoeae* transmission clusters.**
